# Uncertainty and Inconsistency of COVID-19 Non-Pharmaceutical Intervention Effects with Multiple Competitive Statistical Models

**DOI:** 10.1101/2025.01.22.25320783

**Authors:** Bernhard Müller, Inken Padberg, Michael Lorke, Ralph Brinks, Sally Cripps, M. Gabriela M. Gomes, Daniel Haake, John P. A. Ioannidis

## Abstract

Quantifying the effect of non-pharmaceutical interventions (NPIs) is essential for formulating lessons from the COVID-19 pandemic. To enable a more reliable and rigorous evaluation of NPIs based on time series data, we reanalyse the official evaluation of NPIs in Germany. As the first part of a multi-step validation and verification project, we focus on properly analysing statistical uncertainties for time series data. Using a set of 9 competitive statistical methods for estimating the effects of NPIs and other determinants of disease spread on the effective reproduction number ℛ(*t*), we find significantly wider confidence intervals than the official evaluation. In addition to vaccination and seasonality, only few NPIs – such as restrictions in public spaces – can be confidently associated with variations in ℛ(*t*), but even then effect sizes have large uncertainties. Furthermore, due to multicollinearity in NPI activation patterns, it is difficult to distinguish potential effects of NPIs in public spaces from other interventions that came into force early, such as physical distancing. In future, NPIs should be more carefully designed and accompanied by plans for data collections to allow for a timely evaluation of benefits and harms as a basis for an effective and proportionate response.

## INTRODUCTION

The COVID-19 pandemic has arguably been the most globally disruptive event of the 21st century so far. In the aftermath of the pandemic, there is now considerable interest to revisit the handling of the crisis and derive lessons for better responses to similar events in the future.

The effectiveness of interventions to influence the spread of COVID-19 is a key piece of the puzzle in deriving such lessons. Non-pharmaceutical interventions (NPIs) of an unprecedented scale were implemented during the pandemic with substantial collateral effects and at the significant expense of civil liberties. A proper evaluation of both the benefits and harms of such interventions is required since the proportionality of the response is central to the formulation of pandemic strategy [1].

The scientific literature on NPI effects is vast, with enormous heterogeneity in methodology and quality. Official reports on the efficacy of NPIs have therefore been commissioned in countries such as the UK [2], Switzerland [3], and Germany [4] to quantify NPI effects by means of a literature review or a sufficiently comprehensive statistical analysis or meta-analysis of available data and inference models. Similar efforts have also been undertaken outside of official government reviews [5]. Such independent evaluations of NPIs are essential to establish the complete picture and some form of scientific consensus view on the magnitude and uncertainties of NPI effects.

Evidence syntheses based on the literature on NPIs face important limitations, however. Prominent studies of NPI effectiveness (e.g., 6–9) have reached substantively different conclusions both about the effectiveness of individual NPIs and the overall impact of NPIs. The vast majority of NPI studies are observational (sometimes based on data of questionable quality) or merely simulation-based. Consequently, only four studies [10–13] in the extensive review of [2] have been classified as evidence of “moderate” quality in the GRADE [14] assessment framework. Disparate explanatory variables, outcome variables (“apples and oranges”) and methodologies also complicate the comparison of study results, as already pointed out for early studies on the first year of the pan-demic [15]. Most importantly for this study, critical sources of uncertainty in the statistical analysis and the epidemiological assumptions are often not dealt with adequately in the existing NPI literature.

Under these conditions, it is imperative to also improve the reliability of NPI studies individually through increased methodological rigour by better uncertainty quantification and by reducing biases. One key instrument for this purpose is *independent verification and validation* (IV&V) of every step of the analysis pipeline. Furthermore employing multiple models rather than just one may help achieve greater objectivity by fairly representing the range of defensible model assumptions and, under appropriate circumstances, also better predictive performance than any single model by model averaging [16].

Such an approach can help to better assess the robustness of NPI effect estimations and systematically identify key sources of uncertainty that need to be addressed by follow-up studies.

Some IV&V exercises have been conducted during the pandemic on epidemiological models for prediction and inference, and have provided important insights on their uncertainties and sensitivities [17, 18].

We here present the first part of an IV&V project for the evaluation of government interventions on disease spread in Germany by the *StopptCOVID* study. The purpose of our IV&V project is twofold: It seeks to improve NPI effect estimation and uncertainty quantification in the concrete case of Germany, but is also envisaged as a template for similar evaluations of government interventions in other countries in the future.

The original *StopptCOVID* study was commissioned by the German Federal Ministry of Health in 2020 [4] to be carried out by the Robert-Koch Institute (RKI, German Centre for Disease Control) and external collaborators. Importantly, the commissioning of the study was viewed by the constitutional court as partial fulfilment of the legislators’ duty of investigation to ensure proportionality of public health interventions in a key decision from late 2021 [19]. Nevertheless, results were only published as a non-refereed report and released to the press in mid-2023 [20]. Despite critiques of the study methodology [21, 22] the underlying data and the analysis code were not initially made public. They were finally made available to the community early April 2024 [23] after significant political pressure on the German Health Ministry for transparency in a matter of major societal relevance [24].

After the release of the data, we therefore set up an IV&V project to re-examine key findings of the study, both in recognition of the added benefit of a completely independent reanalysis, and of RKI’s limited resources to perform more extensive verification and validation. Because of the complexity of NPI effect inference, the IV&V project is broken up into three work packages. In this paper, we report on Work Package 1, which thoroughly investigates the statistical analysis and uncertainty quantification as a fundamental ingredient for the evaluation of interventions. Work Packages 2 and 3 will address the epidemiological model assumptions and data quality. This multi-step approach means that we initially accept a number of possibly questionable assumptions made in *StopptCOVID* and only obtain provisional results after Work Package 1. But in return such a modular approach makes it easier to ensure that competing models and techniques are objectively assessed at each step, and that competitive solutions for each *component* of the problem (rather than quick fixes for doing everything at once) are effectively identified and implemented. This means building the final product in an “assembly line” approach.

In this spirit, the current study focuses on the proper treatment of *statistical errors* peculiar to the problem of NPI evaluation, in order to obtain estimates and error bars for effects of NPIs and other selected determinants of disease spread on ℛ(*t*). It also seeks to elucidate and compare capabilities and limits of commonly used inference techniques for NPI evaluation, and to identify gaps in data for assessing NPI effectiveness and potential remedies (e.g., the need for certain experimental studies). In keeping with the *Guidelines for Accurate and Transparent Health Estimates Reporting* (GATHER) [25], we provide exhaustive supporting materials containing a literature review, a conceptual overview of the analysis methods and a description of methods to calculate uncertainties (Supplementary SectionsS3–S5). Despite the focus on statistical errors, it is, of course, important to be aware of the future steps of the “assembly line”. The Supplementary Materials therefore also outline some of the issues relevant to the future work packages.

## METHODS

### *StopptCOVID* baseline model

For estimating the effects of NPIs in Germany, *StopptCovid* uses a linear regression model for the logarithm of the time-dependent reproduction number R *_j_*(*t*) for each federal state (“Bundesland”) *j*. NPIs are included as a set of *N*_NPI_ = 51 explanatory variables *X_i_* on a scale from 0 to 1 (see below for details). In addition, the model includes two trigonometric terms for a harmonic seasonal modulation of R *_j_*(*t*) with arbitrary phase, a dependence on the fraction of vaccinated individuals (at least one dose), and fixed effects α *_j_* for state *j*. The model assumes that ln ℛ(*t*) increases by 0.3 and 0.6 times the share ν_α_ and ν_δ_ of the Alpha and Delta variant, respectively. Effects of variants are not estimated but imposed manually as fixed parameters. The effects of vaccination and of NPIs are assumed to occur with lags τ_vac_ and τ_NPI_ with respect to the corresponding explanatory variables. Depletion of susceptibles by infections is neglected.

In terms of these explanatory variables and their regression coefficients β*_i_*, the model for ln R *_j_*(*t*) in state *j* reads,

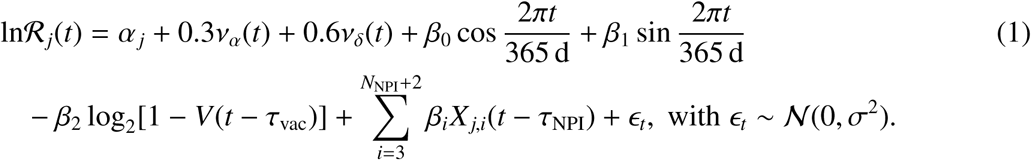

Errors are modelled as normally distributed and uncorrelated across time and states, so that for the observed reproduction number ln R^obs^ at discrete time indices *t* as response variable,

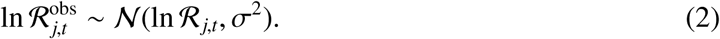

Weighted least-squares (WLS) regression is used for the baseline model, with weights given by 7-day averages of case numbers. This choice of weights can be justified as reducing heteroskedasticity in the observational errors (see below).

Note that the dependence on the vaccination fraction *V* in this model is taken to be non-linear. This particular form of the vaccine effect is problematic, as explained in Supplementary Section S1 C.

*StopptCOVID* determines the delay between interventions and their effect by optimising the model fit based on the Akaike information criterion (AIC; 26). The optimum delay is found to be *negative* (τ_NPI_ = −1 d) for NPIs; and τ_vac_ is found to be 5 d relative to the time of the first dose. The negative delay is problematic (Supplementary Section S1 D), but we accept the delays inferred by *StopptCOVID* as fixed parameters *not subject to errors* for the purpose of our statistical analysis, which is an assumption favourable to the original *StopptCOVID* model.

The study considers the time period from 1 March 2020 until 31 August 2021.

### Data Sources

The effective reproduction number R *_j_*_,*t*_ is calculated from smoothed, 7-day average case data. R *_j_*_,*t*_ is expressed in terms of the incident daily cases I,

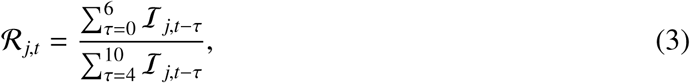

assuming a generation time of τ_gen_ = 4 d. Compared to, e.g., the case data from the Johns Hopkins Coronavirus Resource Center, the *StopptCOVID* dataset uses superior timing information. Instead of using the test or notification date, incident daily case data are based on the date of symptom onset, which is available for almost 60% of cases during the study period and 80% during the first few weeks. Where the time of symptom onset is unknown, RKI uses a careful imputation procedure for symptom onset based on the distribution of notification delay times. Note that for Poisson-like event statistics with var I ∝ I (accounting for the possibility of overdispersion), linearisation of Equation (3) and power-counting yield var 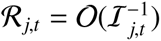 for stochastic noise in R, which justifies the choice of WLS regression.

The explanatory NPI variables are constructed from a detailed, county-level data set of NPIs compiled by *infas* (Institut für angewandte Sozialwissenschaft, Institute for Applied Social Science), which are available online from www.healthcare-datenplattform.de/. The *infas* dataset codes 537 subcategories for 23 main categories of NPIs (e.g., for contacts in private settings, primary and secondary schools, masking). Compared to, e.g., the Oxford Stringency Index [27], the *infas* dataset provides more detailed information on the nature of NPIs with finer spatial resolution, fully based on official data. *StopptCOVID* uses a subset of these subcategories to assign a level to each main category of NPIs. Up to 6 levels for NPI settings are distinguished, with Level 2 (L2) representing the least stringent form of restrictions. The NPI level is determined by the most stringent active restriction. For a detailed breakdown of restrictions at each level, see Supplementary Table II. Due to strong correlations, NPIs for the cultural sector, the hotel and restaurant industry and sports (CHRS) are included in combined categories, depending on how many of these sectors were subject to the highest level of NPI stringency, or failing that, on the second-highest level of stringency.

The different levels are treated as binary variables at the county level. Gaps in the NPI dataset were filled by imputation (last observation carried forward). State-level NPI variables on a continuous scale from 0 to 1 are then constructed as population-weighted averages of the county-level NPI variables, and a lag by τ_NPI_ is applied before these are fed into the linear regression model.

For maximum consistency with *StopptCOVID*, the input data for R, case numbers NPIs are read out from their publicly available R scripts [23]. In line with our strict focus on the statistical analysis, this eliminates the danger of divergent results due to potential misunderstandings about the coding and imputation of the explanatory variables. We highlight, however, that a superficial examination of the data revealed some anomalies. For example, the coded NPI variables do not show any health restrictions in child care facilities in the state of Mecklenburg-Vorpommern in 2020. This contradicts information by the state government [28] and is evidently wrong.

However, the *StopptCOVID* dataset does not include the response variable and the explanatory variables for every day of the period of interest. Data are not provided for short periods without cases in individual states. During these phases, we impute data for all variables by linear interpolation to permit the application of certain analysis methods for time series that cannot easily deal with data gaps.

### Statistical Concerns with the Baseline Model

The results of this baseline model were replicated to very high accuracy. However, based on the model fits and the NPI activation patterns, we diagnosed two important concerns with the statistical analysis. The fitted time series for individual states are shown in Figure 1. For comparison, an overall NPI score (with a maximum value of 47) for each state is also shown. This is defined as

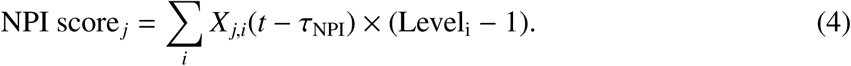

**FIG. 1.**
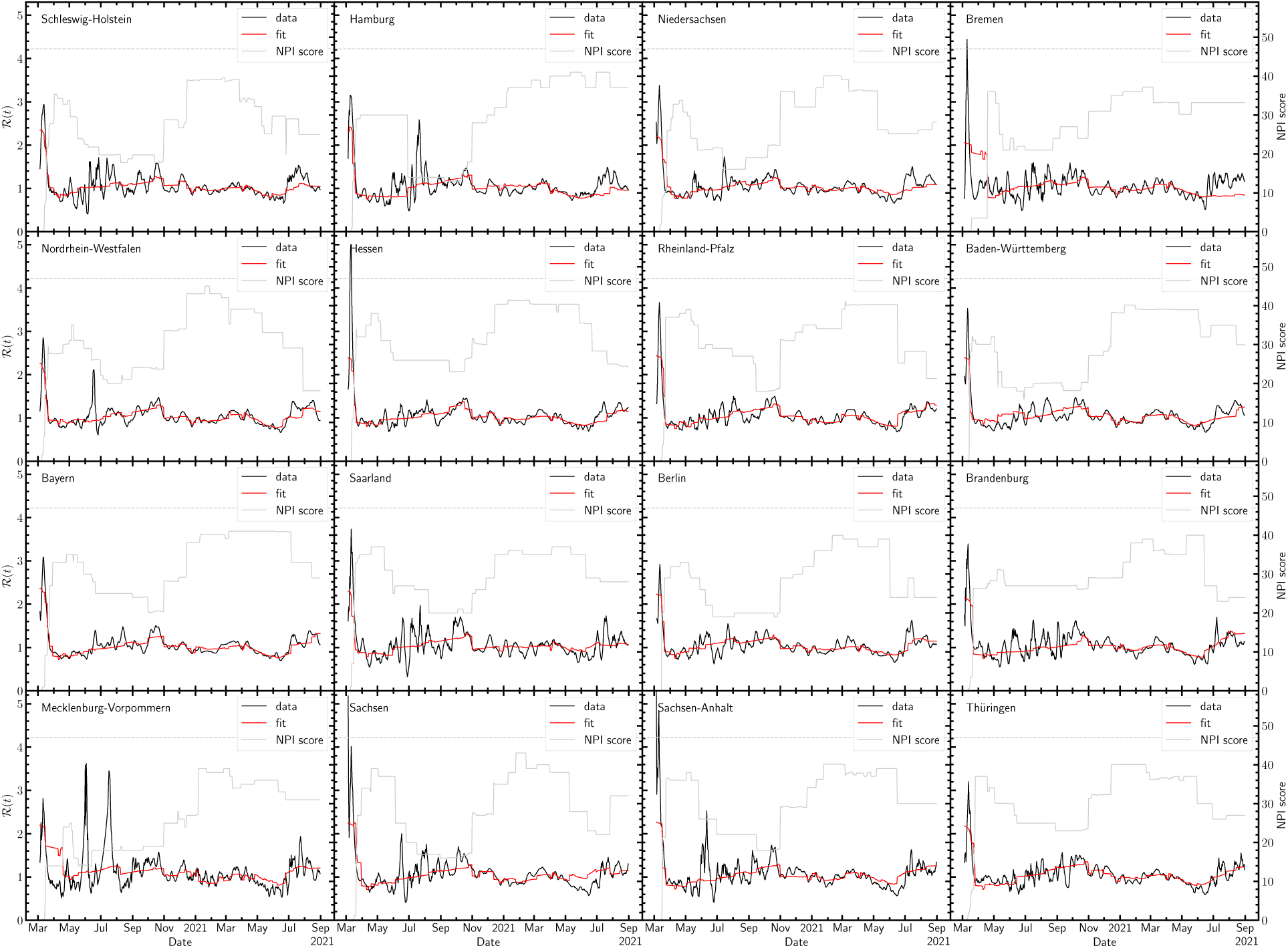
ℛ(*t*) for individual states (black) compared to the fit of the baseline model (red), along with the total NPI score (grey) and its theoretical maximum (grey, dashed).

The level of the combined CHRS category is set to range from 2 to 7 with increasing stringency. We note that despite a coefficient of determination of *R*^2^ = 0.831, the data show substantial dynamics that are not reproduced by the fit. Visual inspection already shows that the errors dis-play *autocorrelation* (i.e., non-independent residuals at adjacent data points), which violates the assumption made by *StopptCOVID* (see also Supplementary Figure 1). Formally, strong autocorrelation is indicated by very low values of the Durbin-Watson statistic [29] of the residuals around 0.2 or less for all federal states. The presence of autocorrelation in the errors terms implies that standard regression errors for effect sizes do not apply and may substantially underestimate the actual errors.

Autocorrelation in the residuals also implies that the evolution of ℛ(*t*) is either affected markedly by *unmodelled processes* (e.g., cluster effects in networks) or by *measurement artefacts* (e.g., ramp-up of testing) that can produce the observed autocorrelation structure in ℛ(*t*), or by both. The fact that the residuals form highly stochastic time series may point to unmodelled processes rather than observational artefacts. There are, e.g., no evident structural breaks in the residuals that would be expected from sudden changes in testing policies. Unmodelled processes or measurement artefacts may have a bigger impact on epidemic dynamics than suggested by the residuals if the baseline model is misspecified; the residuals merely define a minimum level for the magnitude of unmodelled processes or observational noise.

A second issue is *multicollinearity*, i.e., the presence of (strong) correlations among the explanatory variables (i.e., NPI activation variables). Strong multicollinearity leads to a highly ill-conditioned regression problem, and can result in a spurious increase, decrease, or even reversal of effect sizes and inflate the estimated confidence intervals [30, 31]. The degree to which estimates for the regression coefficients for explanatory variables are affected by multicollinearity can be quantified by the variance inflation factor (VIF; see Supplementary Section S1 A for details). Empirical rules-of-thumbs are typically used to identify “serious” multicollinearity, e.g., threshold values of VIF > 10. In the NPI data set, many of the included NPI variables are subject to severe multicollinearity, and some of the VIFs exceed 100.

Given these two problems, there are concerns that the confidence intervals and point estimates from *StopptCOVID* are not valid simply from a statistical perspective. Further limitations due to the epidemiological assumptions are reviewed in Supplementary Section S1. One important concern are uncertainties in the calculation of R, which are not propagated downstream in the current two-step approach from case data to R and then to NPI estimates. The smoothing procedure employed by *StopptCOVID* does not follow established guidelines [32] and introduces a spurious lag (Supplementary Section S1 B). Moreover, the underlying case data are neither drawn from a representative sample, nor at least based on uniform testing criteria. In the current two-step approach of computing R from case data and then estimating NPI effects, the model also neglects any slowdown of epidemic growth due to past infections, which may be aided by heterogeneity effects [33, 34].

### Procedures for IV&V exercise

To select methods for a multi-model IV&V exercise, we adapted the procedures outlined by den Boon et al. [35]. To identify suitable methods for the reevaluation, we applied predefined general selection criteria, namely,

- use in prior NPI studies and widespread use for inference and regression problems in other fields,
- rigorous derivation from first principles,
- the extent by which key problems (e.g., autocorrelation and multicollinearity) were ad-dressed,
- sufficient differentiation from other approaches included in the comparison (to avoid accidentally obtaining similar results by construction),
- and for final inclusion the demonstration of superior/competitive sensitivity and precision when compared to the RKI approach.

To survey methods commonly used in the evaluation of NPI effectiveness, we screened all studies cited in the Royal Society’s recent NPI review [2] examining the effectiveness of NPIs for SARS-CoV-2, unless the study type was deemed not relevant in the context of time series analyses. The following study types were excluded: case report (study) or series, contact survey, randomised control trial. All others were included, even if the use of time series was not made explicit (e.g., ecological studies). In an initial round of screening, we reviewed the subset of studies that considered ℛ(*t*) as outcome, and whose quality of evidence was not rated as *very low* by [2] according to their GRADE assessment [14, 36]. Based on this initial review, we defined various relevant dimensions to broadly categorise all studies according to their different methodological approaches. A description of those categories and dimensions can be found in Supplementary Section S3 (taxonomy of models for NPI effect estimation), S4 (methods for error analysis), and S5 (methods for addressing multicollinearity). The literature review was also used to assess the methodological quality of NPI studies based on time-series or panel data. Screening 338 papers considered by Murphy et al. [2] as well as some other prominent studies revealed that many studies of NPI effects based on time series or panel data suffer from similar limitations in their statistical analysis. For example, temporal autocorrelation is often ignored completely [6]; and even some of the more carefully designed studies [7, 8] still rely on non-rigorous approaches like coarsening of the data or the addition of random-walk component with a pre-specified spectrum.

The results were presented to the project working group. Based mainly on the above selection criteria, a specific subset of methods representing standard approaches for the different categories were chosen (Table I). The selection criteria were interpreted in line with the modular design of the validation project as described in the Introduction. Emphasis was placed on selecting diverse and competitive models for treating the statistical errors peculiar to problems involving panel data and many covariates, and not specficially on the realism and diversity of the epidemiological modelling assumptions. To ensure comparability, the ability to work with the *StopptCOVID* dataset with its specific covariates and R or case data as response variable, and the same epidemiological model assumptions as *StopptCOVID* was a key criterion. To maximise comparability and pin down differences between models, it was also important that they be readily testable and “debuggable” and – related to that – implementable at reasonable effort and computationally efficient. For these reasons, more complex approaches with potentially higher epidemiological fidelity, e.g., data-driven agent-based modelling [37] were not considered at this stage of the project despite their broader merits. The decisions for inclusion or rejection of different approaches are outlined and further discussed in Supplementary Section S3. The chosen methods were implemented using the dataset employed by *StopptCOVID* without altering the epidemiological model assumptions. Finally, a framework for determining effect sizes and uncertainties of summary measures values from all models was agreed upon and documented. For more details on the literature review and decision process, see Supplementary Section S2.

**TABLE I.** Overview of implemented models.

| Model | Model type | Treatment of<br>autocorrelated errors | Shrinkage method for<br>mitigating multicollinearity |
| --- | --- | --- | --- |
| Baseline | Linear regression | None (standard errors) | — |
| DK | Linear regression | Driscoll-Kraay | — |
| Ebisuzaki | Linear regression | Ebisuzaki | — |
| BT | Linear regression | Stationary bootstrap | — |
| 2WFE | Linear regression, two-way fixed effects | Stationary bootstrap | — |
| ARMA($p, q$) | Linear regression | ARMA($p, q$) errors | — |
| DYN | Renewal equation | Stationary bootstrap | — |
| RF | Random forest regression | Stationary bootstrap (cases) | — |
| Elastic net | Linear regression | Stationary bootstrap | Elastic net |
| PCR | Linear regression | Stationary bootstrap | Principal component regression |

An open call for participation was sent to a number of scientific societies at the beginning of the project: Association of the Scientific Medical Societies in Germany (AWMF), German Society for Epidemiology, German Association for Medical Informatics, Biometry and Epidemiology (GMDS), Deutsche ArbeitsGemeinschaft Statistik (DAGStat), German Statistical Society (DStatG), Verein für Socialpolitik e.V., Deutsche Mathematiker-Vereinigung (DMV), Deutsche Physikalische Gesellschaft, Deutsches Klima-Konsortium (DKK), German Reproducibility Network. The German Network for Evidence-Based Medicine and the German Society for Epidemiology kindly disseminated the call, and the German Reproducibility Network provided contact details of member institutions for further distribution.

### Competitive Model Set

Among the methods selected for the multi-model comparison (Table I), models DK, Ebisuzaki, and BT merely use different methods for calculating confidence intervals from the residuals in the case of autocorrelated errors, and are implemented on top of the baseline WLS model. **Model DK** employs the Driscoll-Kraay estimator [38, 39], which uses an estimate of the error covariance matrix up to a specified temporal lag and across entities to compute the variances of the regression coefficients. **Model Ebisuzaki** adapts a frequency-domain method [40] that takes autocorrelation into account by decomposing the residuals into Fourier components and computes confidence intervals based on the power spectrum of the residuals. **Model BT** computes errors using a time series bootstrap [41] that randomly resamples chunks of the time series of residuals such as to preserve their autocorrelation structure. Such a bootstrap is also used for models 2WFE, DYN, RF, Elastic Net and PCR. **Model 2WFE** uses fixed effects both for entities and time [42] to subtract unmodelled temporal dynamics common to all Federal states. Effect estimates for seasonality are obtained in a hierarchical approached by regressing the fixed effects in terms of the seasonal variables. **Model ARMA**(*p*, *q*) models regression errors *n_j_*_,*t*_ as an autoregressive moving-average process of order (*p*, *q*),

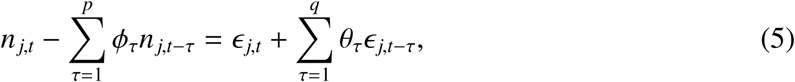

and estimates the regression coefficients and the autoregression coefficients φ_τ_ and θ_τ_ by maximum likelihood estimation using a state-space formulation [43]. The optimal choice (*p*, *q*) = (1, 11) is obtained by minimising the Bayesian information criterion [44]. **Model DYN** combines Equations (1) and (3) into a renewal equation for the 7-day average case data, and is fitted directly to the case data instead of ℛ(*t*). **Model RF** is an implementation of random forest regression [45], which constructs an ensemble of decision trees in the explanatory variables from random samples of the data and then averages the results. The trees are fitted to minimise the squared error. The number and depth of trees and the features considered for the tree splits are optimised by cross validation with a time series split. **Models Elastic Net** and **PCR** use linear regression with regularisation as a possible remedy for multicollinearity. Elastic net regression [46] adds penalty terms to the likelihood for more stable estimates of regression coefficients in exchange for some bias.

Model PCR uses truncated singular value decomposition [47] (non-centred principal component analysis) to filter out patterns in the explanatory variables that contribute to unstable estimates. The regularisation parameters are again determined by cross validation.

Except for model RF, the fitted models immediately yield estimates of linear effects on ln ℛ(*t*) that have exactly the same interpretation as in the baseline model. For model RF, linear effect sizes are extracted as the weighted average difference in ln ℛ(*t*) between two counterfactual scenarios when an intervention is switched on or off completely, while the other NPIs have their actual activation patterns (case RF1). In addition, we also consider this average difference for the case when only seasonal effects and holidays are switched on in the model (case RF0). For the formal definition, see Equations (78,79) in S6. Crudely speaking, cases RF1 and RF0 give estimates for the average marginal effect of an NPI in conjunction with all others, or for the marginal effect it would have as a single, isolated intervention. The partitioning of effects in such machine learning models is non-trivial, and standard approaches, e.g., Shapley values [48, 49] can also face subtle challenges (Supplementary Section S9).

The model set was implemented in Python using statsmodels [50] and sklearn [51]. For a detailed technical description, we refer to Supplementary Section S6.

## RESULTS

### Injection-Recovery Test

To gauge the sensitivity of the models and to test whether disagreement with the baseline model may be the result of bias, we run all the models on synthetic panel data assuming that the baseline model for ℛ(*t*) is correct (for want of experimental data with known effect sizes). This approach (injection-recovery test) is a standard method for determining the sensitivity of analysis methods for time series and other complex data [e.g., 52, 53]. We sort the models into two groups based on whether they can recover the hypothetical effect sizes from *StopptCOVID* without bias with respect to the baseline model (Group A) or not (Group B). Note that since we have no test data with known NPI effects from observations or a “true” model, we can only test the sensitivity of the various methods to an *assumed* signal.

Results of the injection-recovery test are shown in Figures 2 and 3. Models DK (Driscoll-Kraay errors), BT (bootstrap errors), Ebisuzaki, 2WFE (two-way fixed effects) and ARMA(1,11) by construction recover the effects of the baseline model exactly, and hence this test is only a sanity check for their implementation. As it is formulated for consistency with the baseline model, the dynamical model DYN also recovers these effects without bias. With the best-fit hyperparameters from cross-validation, models RF, Elastic Net and PCR all exhibit bias with respect to the baseline model in the injection-recovery test and are therefore assigned to Group B. For the largest effects in the baseline model (such as public spaces L2-L5 and vaccination), the bias is consistently towards the null. In some cases (e.g., COVID tests L3, the sign of the effect in the baseline model is inverted). One notable exception concerns physical distancing L2, to which Group B models consistently ascribe a relatively large effect. Random forest regression ascribes a reduction of about 25% in ℛ(*t*) to physical distancing relative to the state without interventions (case RF0). The reduction of the effects for public spaces and the increased effect of physical distancing L2 is essentially a reassignment of effects within groups of NPIs that were in place (at some level) quite consistently from the early phase of the pandemic.

**FIG. 2.**
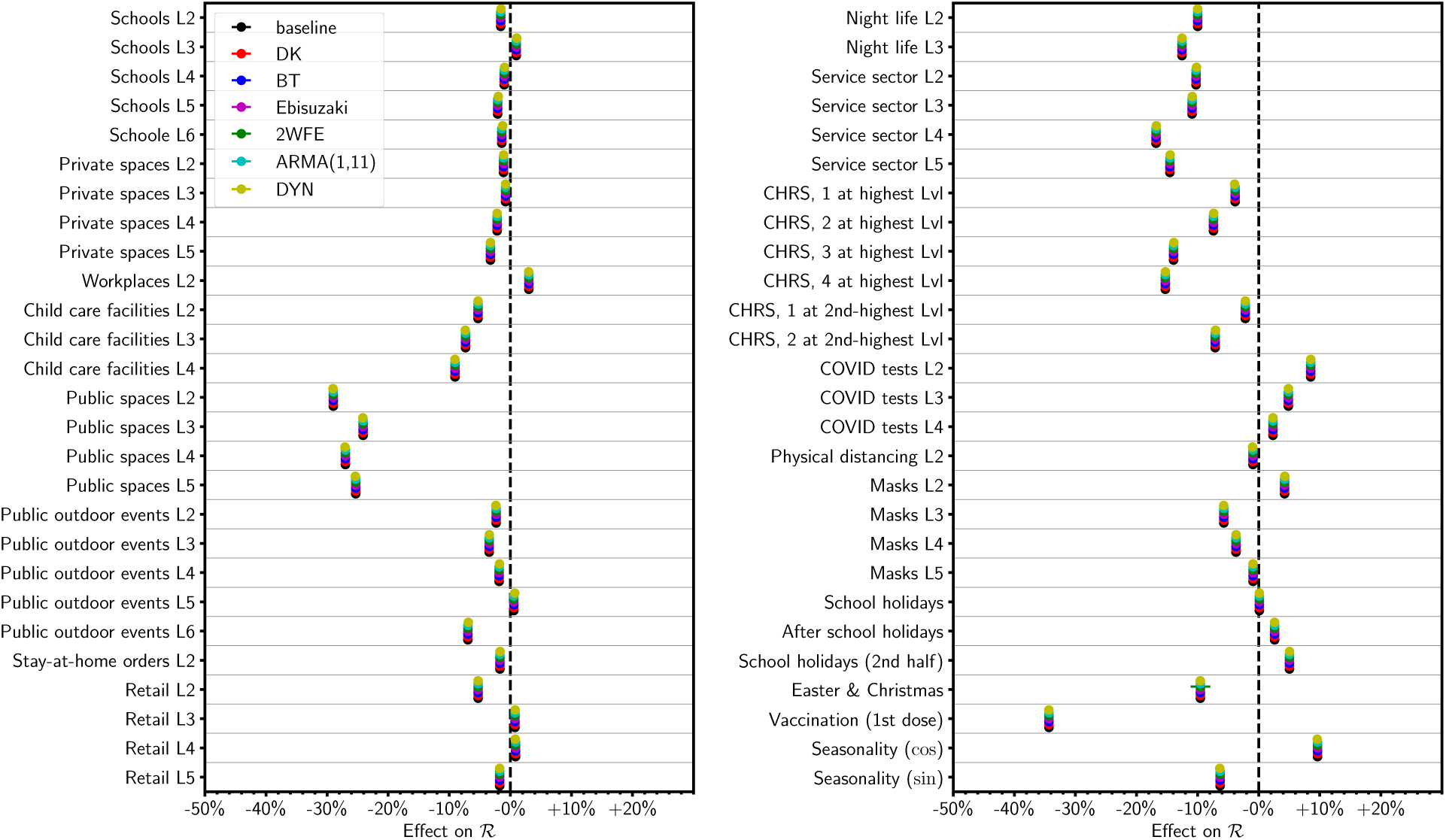
Injection-recovery test for model Group A. For the models in this group, this is merely a sanity check for correct implementation. Numbers L*x* indicate stringency levels of NPIs. Note that CHRS is a combined category for the cultural sector, the hotel and restaurant industry and sports.

**FIG. 3.**
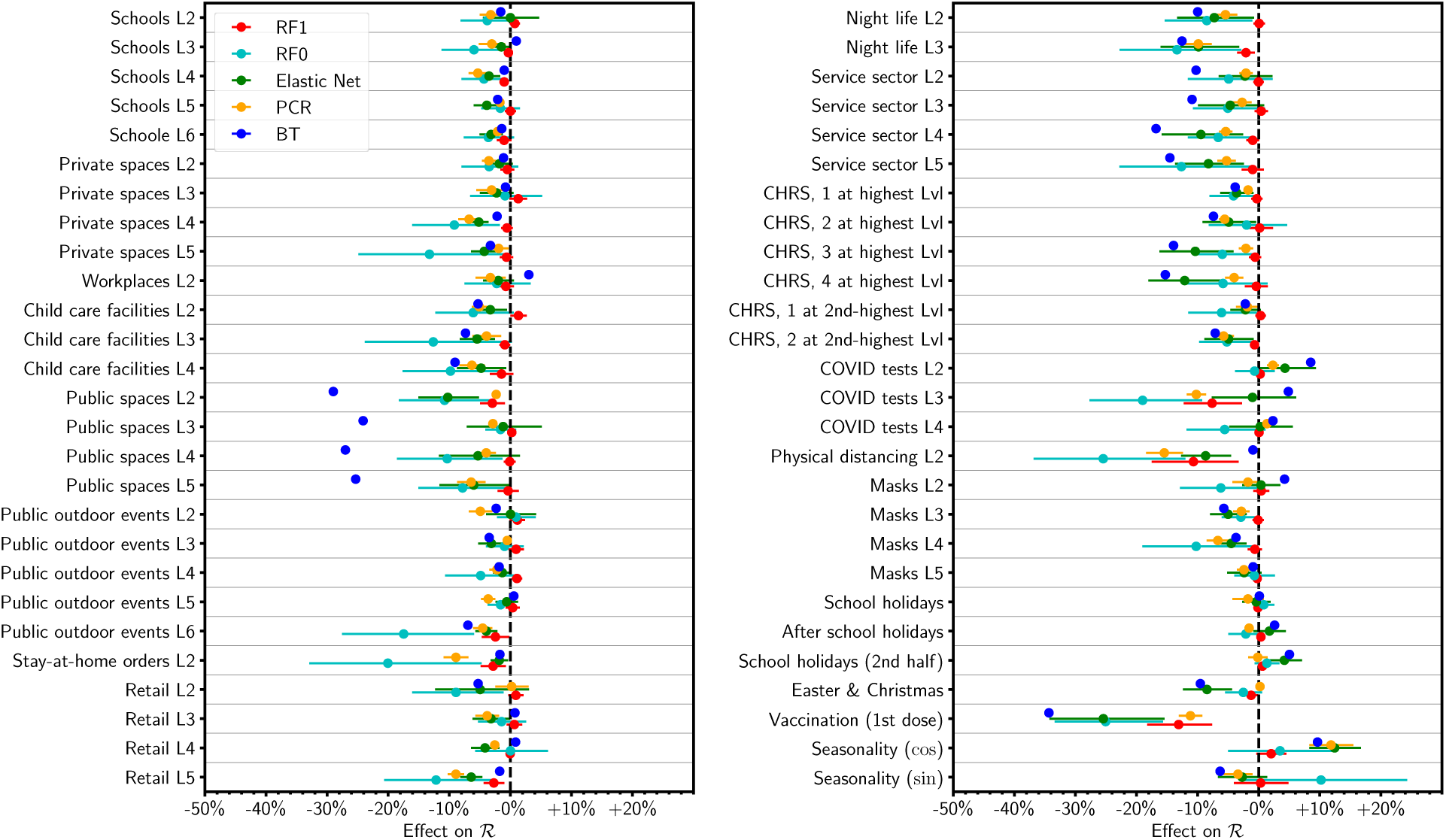
Injection-recovery test for model Group B. The injection-recovery test shows that these models cannot exactly recover the point estimates for the baseline model (identical to model BT in), but are subject to bias.

Random forest regression yields different linear effect estimates depending on the reference state for the linearisation of the model. The average marginal effects of NPIs on top of the other NPIs (case RF1) is smaller than the effect in the baseline model, whereas the predicted marginal effects of NPIs as isolated interventions (case RF0) is larger. For example, case RF0 yields large effects – though with big error bars – for public outdoor events L6, stay-at-home-orders and physical distancing. Clearly, neither the RF0 nor the RF1 estimates from model RF are satisfactory in the injection-recovery test. Further analysis of the behaviour of model RF is provided in Supplementary Section S9. This analysis also reveals that the effect sizes for the sine and cosine component in random forest regression *cannot* be interpreted as amplitudes of the seasonal variation, and will therefore be discarded in the subsequent discussion.

The bias in the Group B models does not render these models incorrect for effect estimation. By design, the Elastic Net and PCR models produce more *parsimonious representations* to the actual data for ℛ(*t*) and *may* filter out noise in the effect estimates. However, they cannot provide direct evidence to *rule out* the baseline model because they would not recover its estimates even if this model were true. However, because these models would not reproduce the baseline model’s estimates even if the baseline were correct, they cannot be used to directly evaluate or exclude it. Group A therefore forms the primary basis for our conclusions, while Group B serves to aid and temper the interpretation of the inferred statistical associations.

Comparing the results of random forest regression to linear models requires particular care. The model suggests that non-linear interactions of NPIs are important and *may* correctly perceive saturation effects, i.e., little additional effect by single NPIs when many others are switched on already, and its effect estimates in some sense correctly reflect the difficulty of distinguishing the effects of NPIs with similar activation patterns. Random forest regression may be useful for generating hypotheses for non-trivial interactions between NPIs. More work and additional, independent data are required to compare the relative merit of random forest regression and simple additive or multiplicative models for NPI effects.

### Effect estimates and confidence intervals

Effect estimates and confidence intervals for model groups A and B are shown in Figure 4 and 5, respectively, and are also listed in Supplementary Table III. To facilitate comparison between the two groups, the results for model BT (linear regression with bootstrap errors) are also included in Figure 5.

**FIG. 4.**
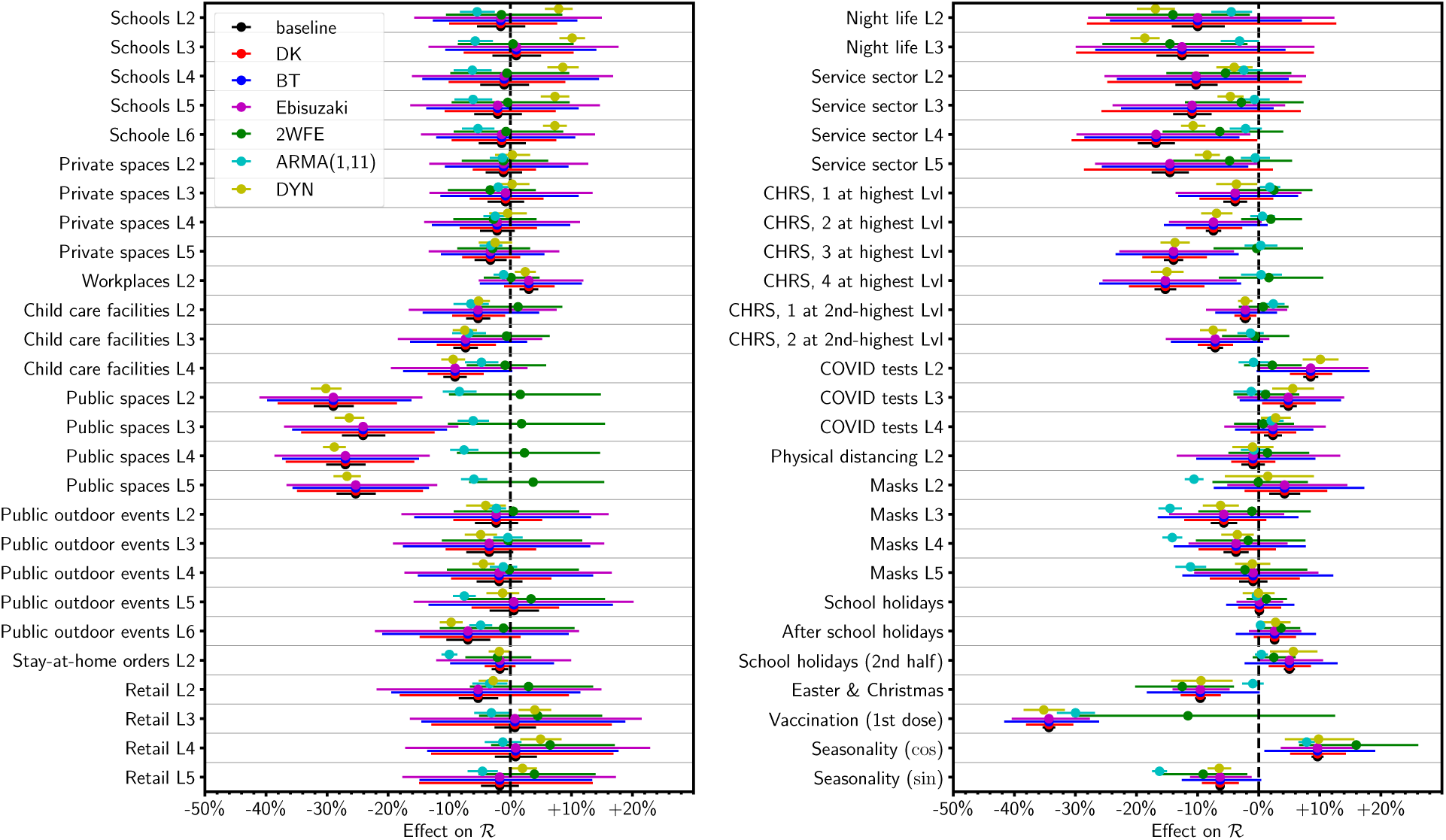
Point estimates and confidence intervals for model Group A.

**FIG. 5.**
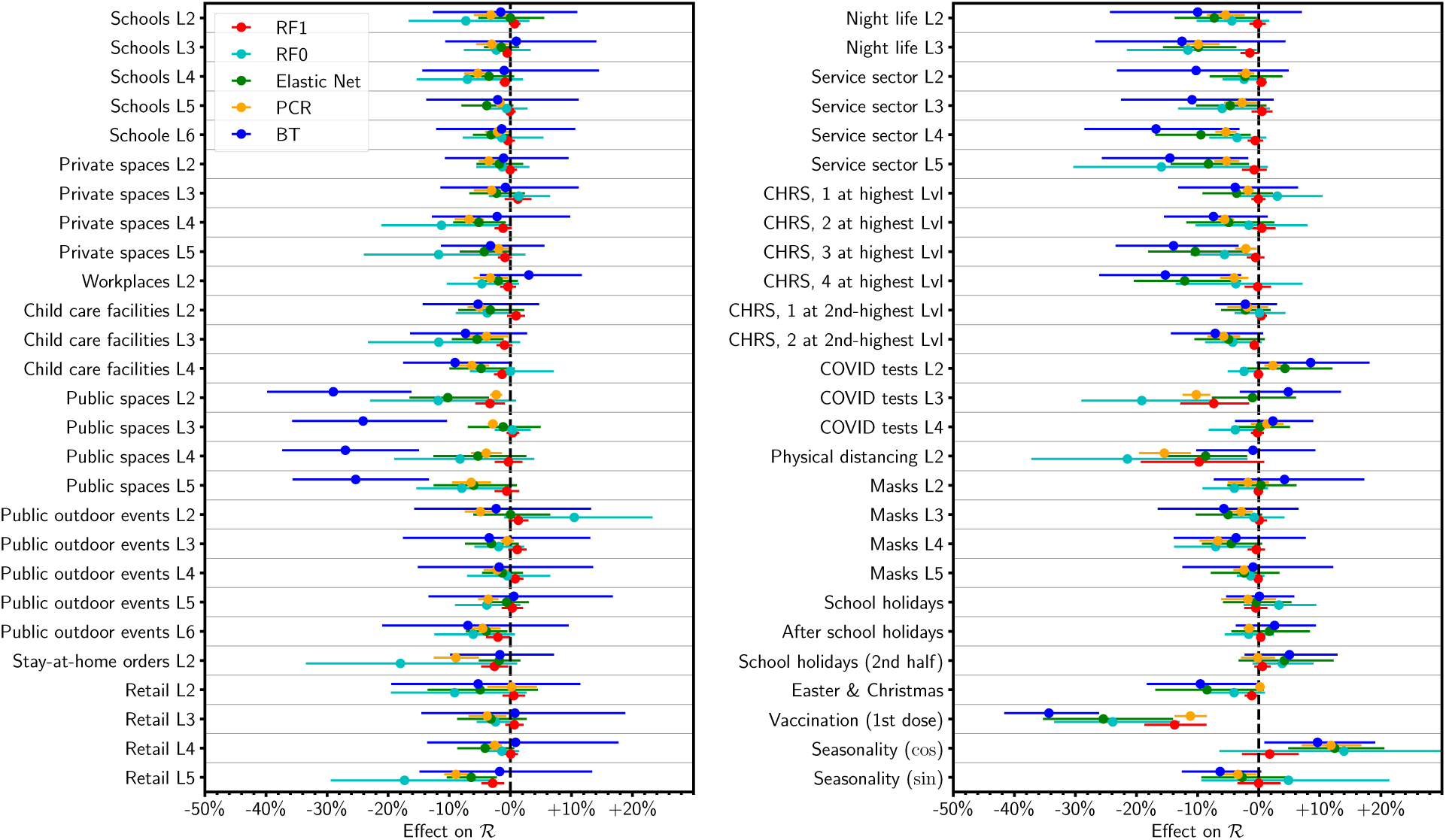
Point estimates and confidence intervals for the (biased) models in group B, compared to linear regression with bootstrap errors.

#### Model Group A

Within Group A, the effect sizes in the linear regression models DK, BT, and Ebisuzaki trivially agree with the baseline model. The confidence intervals for these models are much wider than for the baseline model from the original *StopptCOVID* study. Confidence intervals calculated used Driscoll-Kraay errors, Ebisuzaki’s method, or the stationary bootstrap are generally very similar.

Model 2WFE generally yields similar effect estimates and confidence intervals, with a few notable exceptions. First, the estimated effects of NPIs for public spaces and several NPI levels for the cultural sector, the hotel and restaurant industry and sports (CHRS) are close to the null, and in some cases, the confidence intervals do not overlap with the DK, BT, and Ebisuzaki models. The only NPIs for which the model yields a significant beneficial effect are those for night life (L2–3). Second, the estimated vaccine effect is considerably smaller, but with a very large confidence interval that overlaps with those of models DK, BT, and Ebisuzaki. The reason for the larger error bar lies in the close synchronisation of vaccination across the federal states. The two-way fixed effects model is, in a sense, optimised to detect effect based on difference between the response variable and explanatory variables across entities and therefore struggles to deliver a precise estimate. We take this into account by constructing assessment criteria that are robust to such an outlier result.

Regression with ARMA(1,11) errors gives similarly narrow confidence intervals, and in a few cases even narrower confidence intervals, than the baseline model, but the interval for vaccination remains substantially wider. The effect estimates, however, often differ markedly from the baseline model. The effect estimates for public spaces are considerably smaller. In turn, the model ascribes more than a 10% reduction in R to masks and stay-at-home orders and small, but statistically significant effects to a few others NPIs, e.g., in schools. Generally, the point estimates and error bars for masks are compatible with findings from randomised and cluster-randomised trials [54–56] that also indicate no or modest effects at the real-world population level.

The DYN model (renewal equation) tends to yield effect estimates within the error bars of the DK, BT, and Ebisuzaki models. Thus, fitting the case data instead of ℛ(*t*) yields relatively consistent results within these “safe” error bars. However, the confidence intervals for model DYN are often as narrow as for the baseline model, sometimes even narrower (e.g., for public outdoor events), and sometimes wider (e.g., for masks). The confidence intervals often do *not* overlap with the baseline model. This suggests that the bootstrapping procedure used for model DYN does not yet fully account for autocorrelation.

Overall, however, the Group A confidence intervals for most explanatory variables overlap well and the scatter between the point estimates of different models tends to be bounded by the DK, BT, and Ebisuzaki error bars. Notable exceptions include the NPIs for public spaces, some NPIs for the cultural sector, the hotel and restaurant industry and sports (CHRS), vaccination (with the DYN and 2WFE models as outliers) and the sine component of the seasonal modulation. In the case of the vaccine effect, the relative uncertainty due to the between-model scatter is modest compared to the large effect size, however.

Following *StopptCOVID*, the vaccine effect intends to represent the result of halving the fraction of unvaccinated individuals. As the original implementation leads to pathological behaviour in the limit of a high vaccination fraction, we also considered a modification of model DYN that correctly implements the vaccine effect (Supplementary Section S1 C). The corrected model yields a nominal vaccine efficacy of about 90% against infection, and the other effect estimates remain within the “safe” BT, DK and Ebisuzaki error bars. The bulk of first-dose vaccination occurred about three months before the end of the study period, i.e., the waning of vaccine efficacy played a lesser role. The estimated vaccine efficacy is therefore roughly consistent with the high short-term efficacy against infection inferred by the clinical trials and cohort studies [57–60], especially bearing in mind that some conflation of the effects of the first- and second-dose may be implicit in the *StopptCOVID* model.

### Model Group B

The models with explicit shrinkage and the RF1 estimates from random forest regression all yield significantly narrower confidence intervals than non-regularised regression with bootstrap errors (which is the purpose of shrinkage in the first place). The RF1 estimates from random forest regression and principal component regression tend to shrink the confidence intervals even more strongly. The RF0 estimates do not show much shrinkage and sometimes result in wider confidence intervals than linear regression with bootstrap errors.

With regard to point estimates, there is a rough tendency of elastic net and principal component regression to shrink strong effects in the baseline model to modest or small effects, and to magnify a few small effect estimates. As in the recovery-injection test, random forest regression tends to yield very small effects for many NPIs if the actual NPI activation is used as reference state (case RF1), but some very large effects for single NPIs without any other concurrent interventions (case RF0).

The prominent cases where the models with significant shrinkage of the confidence intervals yield *smaller* effects outside the BT error bars are the NPIs for public spaces – as the ARMA(1,11) and 2WFE models in the previous subsection – and vaccination. Exactly as in the injection-recovery tests, the models with shrinkage prefer to attribute a greater effect to physical distancing, and to some extent to policy COVID tests L3.

### Ranking of Effects

Despite considerably wider error bars than in *StopptCOVID*, some statistically significant associations of variations in R with interventions or environmental factors can be detected. Visually, the effects of vaccination and seasonality emerge most clearly. For vaccination, only model 2WFE has a confidence interval that overlaps with zero, which we consider an outlier for reasons described above. For the sine component of seasonality, only one confidence interval marginally overlaps with the null, and all Group A models shows a significant cosine modulation. This is reinforced by the regularised regression models (elastic net and PCR), which also show a significant cosine component.

For a more quantitative identification of the NPIs that may be associated with lower ℛ(*t*), we define two different scores (Tables II and III) to quantify how confidently a null effect can be excluded based on within-model error bars and between-model consistency. The first score is the number of models that disagree with the sign of the median effect estimate across models plus the number of models with confidence intervals that overlap with the null.

**TABLE II.** Integer score for ranking the likelihood of a real association of covariates with ℛ(*t*) based on model Group A. Lower scores are better. Only the top-14 NPIs are shown. Scores of up to 2 are rated as potentially indicative of a real association with variations in ℛ(*t*) (see text).

| Explanatory<br>variable | Disagreement<br>on sign | Overlap<br>with null | Sum<br>(integer score) |
| --- | --- | --- | --- |
| Seasonality (cos) | 0 | 0 | 0 |
| Vaccination (1st dose) | 0 | 1 | 1 |
| Easter & Christmas | 0 | 1 | 1 |
| Seasonality (sin) | 0 | 1 | 1 |
| Service sector L4 | 0 | 2 | 2 |
| Child care facilities L4 | 0 | 2 | 2 |
| Public spaces L2 | 1 | 1 | 2 |
| Public spaces L4 | 1 | 1 | 2 |
| Public spaces L5 | 1 | 1 | 2 |
| Public spaces L3 | 1 | 1 | 2 |
| Service sector L5 | 0 | 3 | 3 |
| Night life L2 | 0 | 3 | 3 |
| Child care facilities L3 | 0 | 3 | 3 |
| CHRS, 3 at highest Lvl | 1 | 2 | 3 |

**TABLE III.** False positive risk score for the 14 most highly-ranked explanatory variables based on model Group A. Scores lower than 0.1 are rated as indicative of a real association with variations in ℛ(*t*) (see text).

| Explanatory<br>variable | False positive<br>risk score |
| --- | --- |
| Seasonality (cos) | 0.002 |
| Seasonality (sin) | 0.009 |
| Vaccination (1st dose) | 0.026 |
| Easter & Christmas | 0.028 |
| Service sector L4 | 0.037 |
| Night life L3 | 0.058 |
| Child care facilities L4 | 0.081 |
| Night life L2 | 0.082 |
| Service sector L2 | 0.092 |
| Service sector L5 | 0.097 |
| Public spaces L2 | 0.101 |
| CHRS, 2 at 2nd-highest Lvl | 0.101 |
| Child care facilities L3 | 0.101 |
| Public spaces L3 | 0.102 |

The second score (“false positive risk score”) is the average of the false-positive probabilities *Q* (for estimating an effect ≤ β*_i_* when the true effect is null) in the models. *Q* is computed from the cumulative *t*-distribution as appropriate for errors of linear regression coefficients,

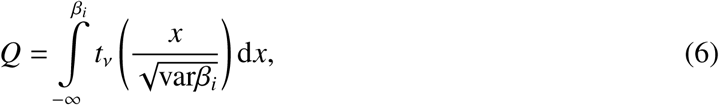

where the number of degrees of freedom ν is the number of observations minus the number of federal states and explanatory variables. We use the exact *t*-distribution, although ν is so large that the normal approximation applies. In line with guidelines for multi-model comparisons [35], one should avoid interpreting the resulting metric as a probability. Both scores are merely heuristic scales that penalise lack of significance and between-model variation. Recognising the bias in Group B models, these scores are computed only for Group A, and the baseline model is also excluded because of its unrealistic error model.

The explanatory variables with the top-14 scores are shown in Tables II and III, respectively. As a cut-off for a potentially indicative association of an NPI with lower ℛ(*t*), we tentatively suggest that there should *either* be no more than two vetos (integer score of two or less), *or* that the average false-positive risk score should not exceed 0.1. This allows us to accept effects as likely even when a model is a clear outlier, or when a few models do not find a significant effect, but all point estimates clearly cluster on one side. Along with vaccination, seasonality and Easter & Christmas, this leaves NPIs for public spaces, the service sector (L4), night life and child care facilities (L4) as the best candidates for associations with lower ℛ(*t*). Among these, we rate the effect of restrictions in public spaces as most statistically robust because only model 2WFE fails to find a significant association with lower ℛ(*t*) and is responsible for the lower ranking of these NPIs on the second score.

The models with shrinkage serve to temper the scores in Tables II and III, however. They indicate that the effects of NPIs for public spaces and night life may be weaker and hard to distinguish from the effects of physical distancing. Rather than blindly accepting the scores in Tables II and III, further research is required to better distinguish the effects of classes of NPIs that came into force early during the pandemic and therefore tend to be assigned large effects by the models.

## DISCUSSION

Our set of competitive models demonstrates that the confidence intervals from the original evaluation of German NPIs by *StopptCOVID* are substantially too narrow. They are neither consistent with a more rigorous error analysis for linear regression, nor with the between-model variation. *The data for* ℛ(*t*) *and NPIs are insufficient for confidently assigning effects to most NPIs.* More-over, the confidence intervals are generally so wide that it is impossible to confidently detect trends with increased stringency, contrary to claims by *StopptCOVID*. Especially for NPIs in public spaces, even the point estimates do not suggest additional benefits from higher stringency levels, similar to other recent claims in the literature [61].

It is important to point out that any inferred effects on ℛ(*t*) still need to be translated into relevant public health outcomes (e.g., total or peak hospitalizations, years or quality-adjusted years of lives saved) for a satisfactory assessment of interventions. This complex task requires additional information *beyond* the effects of NPIs on ℛ(*t*). Moreover, estimation of NPI effects may have even more error and uncertainty than what we estimated here, if data are unreliable, a common feature in the chaotic circumstances of the COVID-19 pandemic.

Furthermore, the epidemic model considered in this study cannot determine feedback effects and non-linear effects can impact both disease spread and the risk factors; hence constructing counterfactual trajectories of ℛ(*t*) can be misleading. For example, self-regulating behaviour in the population [62, 63] or population heterogeneity [33, 34] may lead to a slow-down of disease spread independent of NPIs. Time dependence, in particular waning of immunity after vaccination or infection is another relevant complication that can lead to counterinuitive results such as “immunity debt” [64]. In the context of vaccination against seasonal respiratory diseases, higher vaccine coverage can under certain conditions increase infection peaks [65–70]. Similarly unintuitive disease dynamics – which obviously cannot be translated directly to COVID-19 – has also long been studied for rubella [71, 72] and measles [73]. The implications of temporary vaccine-induced immunity should only be assessed based on a sufficient understanding of the principles of time-dependent disease dynamics.

We are forced to conclude that the approach taken by the German Federal Ministry of Health is insufficient for ascertaining public health outcomes of NPIs given the revealed statistical limitations. In fact, much of the prior literature on time-series based NPI studies does not adequately address critical statistical problems like autocorrelation and multicollinearity either. As under-estimation of NPI effect uncertainties is likely a wider problem, we recommend that key results be subjected to a similar reanalysis to provide reliable information for pandemic planning to policymakers. Without rigorous uncertainty quantification, pandemic management suffers from an impaired feedback loop between research and decision-making: Decision-makers cannot reliably determine whether their interventions achieve the desired outcomes, and may overlook the need to consider alternatives when they do not. Failure to quantify and report uncertainty may also erode public trust in policy decisions.

To enable robust evaluation and adjustments of NPIs to ensure proportionality and balance benefits and harms, future public health interventions should be designed with sufficient pre- and post-intervention observation times to permit a meaningful determination of effects. This should be coupled with careful epidemiological considerations and plans for relevant data collection [74]. Where a preliminary cost-benefit analysis suggests rough equipoise, interventions should include a control group. Pandemic research programs need to be coordinated to systematically identify knowledge gaps and ensure that data for interventions effectiveness in relevant settings (hospitals, nursing homes, etc.) are obtained to complement studies of population-wide disease spread.

State-of-the art time series analysis should be used to inform the required study design or observation periods for non-controlled designs, similar to the use of sample size calculations to ensure sufficient power for other epidemiological studies. When complex models are used for analysing epidemiological data, forecasting or scenario development, independent validation and verification (as outlined in this paper) should be sought by decision-makers for key modelling studies. Employed methods should be (or have been) tested for sensitivity and specificity using tools like injection-recovery tests. In future, policymakers and funders should more broadly support validation research, and actively seek validation for critical policy-relevant research.

## Supporting information

Supplementary materials (single file)

## DATA AVAILABILITY

Our Python code is freely available on Zenodo (https://doi.org/10.5281/zenodo.14761068). The code utilises processed data from the original *StopptCOVID* project [23], which in turn uses NPI data from *infas* (www.healthcare-datenplattform.de/), which are freely available after registration. Our code repository contains instructions to download the required third-party data and code to generate the requisite input data for our analysis.

## ACKNOWLEDGEMENTS

We thank M. an der Heiden and V. Bremer for answering technical questions on the original *StopptCOVID* project. We acknowledge helpful discussions with W. Baumgarten, O. Beige, G. Meyer, I. Mühlhauser, D. Schuricht, and T. Wieland. We are grateful to the German Network for Evidence-Based Medicine, the German Society for Epidemiology and the German Reproducibility Network for help in distributing the project call.

## AUTHOR CONTRIBUTIONS

B.M.: Conceptualisation, Project Management, Software, Formal analysis, Visualisation, Writing – Original draft, Review and Editing. I.P.: Conceptualisation, Procedures, Literature Review, Writing – Original draft, Review and Editing. ML: Conceptualisation, Code checks (models DK and Ebisuzaki), Writing – Review. R.B.: Conceptualisation, Writing – Review and Editing. S.C.: Conceptualisation, Writing – Review and Editing. M.G.M.M.: Conceptualisation, Writing – Re-view. D.H.: Conceptualisation, Writing – Review and Editing. J.P.A.I.: Conceptualisation, Writing – Review and Editing.

## FUNDING

There is no funding to report.

## ETHICS DECLARATION

This study did not involve research on humans or animals, and only used publicly available, non-personal data sets.

## COMPETING INTERESTS

B.M. I.P. M.L., and R.B. are signatories of a call for a non-partisan pandemic review in Ger-many (https://pandemieaufarbeitung.net). B.M. has been engaged in discussion and consultation of pandemic policy and science policy with members of several German parties (CDU, CSU, FDP, SPD, BSW, Greens), but is not receiving remuneration. S.C., M.G.M.G., D.H., and J.P.A.I. have no competing interests to declare.

## Notes

### Competing Interest Statement

B.M., I.P., M.L., and R.B. are signatories of a call for a non-partisan pandemic review in Germany (https://pandemieaufarbeitung.net).
B.M. has been engaged in discussion and consultation of pandemic policy and science policy with members of several German parties (CDU, CSU, FDP, SPD, BSW, Greens), but is not receiving remuneration.
S.C., M.G.M.G., D.H. and J.P.A.I. have no competing interests to declare.

### Summary of Updates

Revised version, accepted for publication in Scientific Reports. Restructured manuscript with extended introduction and discussion section and additional supplementary materials. Minor changes in figures.

## References

[1] UK Covid-19 Inquiry. Module 1 report: The resilience and preparedness of the United Kingdom (2024). URL https://covid19.public-inquiry.uk/reports/module-1-report-the-resilience-and-preparedness-of-the-united-kingdom/.

[2] Murphy, C. et al. Effectiveness of social distancing measures and lockdowns for reducing transmission of COVID-19 in non-healthcare, community-based settings. Philos. Trans. A Math. Phys. Eng. Sci. 381, 20230132 (2023).

[3] Funk, M., et al. Wirksamkeit von Corona-Massnahmen in der Schweiz (2022). URL https://www.seco.admin.ch/seco/de/home/Publikationen_Dienstleistungen/Publikationen_und_Formulare/Strukturwandel_Wachstum/Wachstum/wirksamkeit-corona-massnahmen-schweiz.html.

[4] Bundesministerium für Gesundheit. Evaluation der Rechtsgrundlagen und Maßnahmen der Pan-demiepolitik. Bericht des Sachverständigenausschusses nach §5 Abs. 9 IfSG (2022). URL https://www.bundesgesundheitsministerium.de/fileadmin/Dateien/3_Downloads/S/Sachverstaendigenausschuss/BER_lfSG-BMG.pdf.

[5] Iezadi, S. et al. Effectiveness of non-pharmaceutical public health interventions against COVID-19: A systematic review and meta-analysis. PLoS One 16, e0260371 (2021).

[6] Flaxman, S. et al. Estimating the effects of non-pharmaceutical interventions on COVID-19 in europe. Nature 584, 257–261 (2020).

[7] Sharma, M. et al. Understanding the effectiveness of government interventions against the resurgence of COVID-19 in europe. Nat. Commun. 12, 5820 (2021).

[8] Bendavid, E., Oh, C., Bhattacharya, J. & Ioannidis, J. P. A. Assessing mandatory stay-at-home and business closure effects on the spread of COVID-19. Eur. J. Clin. Invest. 51, e13484 (2021).

[9] Bendavid, E. & Patel, C. J. Epidemic outcomes following government responses to covid-19: Insights from nearly 100,000 models. Science Advances 10, eadn0671 (2024). URL https://www.science.org/doi/abs/10.1126/sciadv.adn0671. https://www.science.org/doi/pdf/10.1126/sciadv.adn0671.

[10] Ramirez, D. W. E., Klinkhammer, M. D. & Rowland, L. C. COVID-19 transmission during transportation of 1st to 12th grade students: Experience of an independent school in virginia. J. Sch. Health 91, 678–682 (2021).

[11] Helsingen, L. M. et al. Covid-19 transmission in fitness centers in norway - a randomized trial. BMC Public Health 21, 2103 (2021).

[12] Hunter, P. R., Colón-González, F. J., Brainard, J. & Rushton, S. Impact of non-pharmaceutical interventions against COVID-19 in europe in 2020: a quasi-experimental non-equivalent group and time series design study. Euro Surveill. 26 (2021).

[13] Wang, G. Stay at home to stay safe: Effectiveness of stay-at-home orders in containing the COVID-19 pandemic. Production and Operations Management 31, 2289–2305 (2022). URL https://ideas.repec.org/a/bla/popmgt/v31y2022i5p2289-2305.html.

[14] Guyatt, G. et al. GRADE guidelines: 1. Introduction-GRADE evidence profiles and summary of findings tables. J. Clin. Epidemiol. 64, 383–394 (2011).

[15] Banholzer, N. et al. The methodologies to assess the effectiveness of non-pharmaceutical interventions during covid-19: a systematic review. European Journal of Epidemiology 37, 1003–1024 (2022). URL 10.1007/s10654-022-00908-y.

[16] Raftery, A. E., Madigan, D. & Hoeting, J. A. Bayesian Model Averaging for Linear Regression Models. Journal of the American Statistical Association 92, 179–191 (1997). URL 10.1080/01621459.1997.10473615.

[17] Edeling, W. et al. The impact of uncertainty on predictions of the CovidSim epidemiological code. Nat. Comput. Sci. 1, 128–135 (2021).

[18] Chin, V., Ioannidis, J. P. A., Tanner, M. A. & Cripps, S. Effect estimates of COVID-19 non-pharmaceutical interventions are non-robust and highly model-dependent. J. Clin. Epidemiol. 136, 96–132 (2021).

[19] Bundesverfassungsgericht. decisions 1 BvR 971/21 and 1 BvR 1069/21. https://www.bundesverfassungsgericht.de/SharedDocs/Entscheidungen/DE/2021/11/rs20211119_1bvr097121.html (2021).

[20] an der Heiden, M., Hicketier, A. & Bremer, V. Wirksamkeit und Wirkung von anti-epidemischen Maßnahmen auf die COVID-19-Pandemie in Deutschland (StopptCOVID-Studie) (2023). URL https://www.rki.de/DE/Content/InfAZ/N/Neuartiges_Coronavirus/Projekte_RKI/StopptCOVID_studie.html.

[21] Meyer, G., Mühlhauser, I., Brinks, R. & Müller, B. Wirksame Kontrollmaßnahmen in der SARS-CoV-2-Pandemie? KVH Journal 10/2023 (2023).

[22] Baumgarten, W., Beige, O., Haake, D., Merkl, J. & Wieland, T. Was die ‘StopptCOVID’-Studie des RKI sagt - und was nicht. Cicero Online (2023). URL https://www.cicero.de/innenpolitik/corona-pandemie-robertkochinstitut-studie.

[23] Hicketier, A. & an der Heiden, M. StopptCOVID-Studie - Daten, Analyse und Ergebnisse (2024). URL https://zenodo.org/records/10888033.

[24] Bodderas, E. Kanzleramt drängt Lauterbach zur Offenlegung von Corona-Studie. WELT, 8.2.2024 (2024). URL https://www.welt.de/politik/deutschland/plus250799234/Pandemie-Aufarbeitung-Kanzleramt-draengt-Lauterbach-zur-Offenlegung-von-Corona-Studie.html.

[25] Stevens, G. A. et al. Guidelines for accurate and transparent health estimates reporting: The GATHER statement. Lancet 388, e19–e23 (2016).

[26] Akaike, H. Information Theory and an Extension of the Maximum Likelihood Principle, 199–213 (Springer New York, New York, NY, 1998). URL 10.1007/978-1-4612-1694-0_15.

[27] Roser, M. What is the covid-19 stringency index? Our World in Data (2021). Https://ourworldindata.org/metrics-explained-covid19-stringency-index.

[28] State Government of Mecklenburg-Vorpommern. Kitas und Kindertagespflege ab Montag flächen-deckend geschlossen (2020). URL https://www.regierung-mv.de/Landesregierung/sm/Aktuell/?id=158498&processor=processor.sa.pressemitteilung.

[29] Durbin, J. & Watson, G. S. Testing for serial correlation in least squares regression. i. Biometrika 37, 409–428 (1950). URL 10.1093/biomet/37.3-4.409.

[30] Greene, W. H. Econometric Analysis (Pearson Education, 2003), fifth edn. URL http://pages.stern.nyu.edu/∼wgreene/Text/econometricanalysis.htm.

[31] Johnston, R., Jones, K. & Manley, D. Confounding and collinearity in regression analysis: a cautionary tale and an alternative procedure, illustrated by studies of British voting behaviour. Quality & Quantity 52, 1957–1976 (2018). URL 10.1007/s11135-017-0584-6.

[32] Gostic, K. M. et al. Practical considerations for measuring the effective reproductive number, rt. PLOS Computational Biology 16, 1–21 (2020). URL 10.1371/journal.pcbi.1008409.

[33] Neipel, J., Bauermann, J., Bo, S., Harmon, T. & Jülicher, F. Power-law population heterogeneity governs epidemic waves. PLoS ONE 15, e0239678 (2020). 2008.00471.

[34] Gomes, M. G. M. et al. Individual variation in susceptibility or exposure to SARS-CoV-2 lowers the herd immunity threshold. Journal of Theoretical Biology 540, 111063 (2022). URL https://www.sciencedirect.com/science/article/pii/S0022519322000613.

[35] den Boon, S., et al. Guidelines for multi-model comparisons of the impact of infectious disease interventions. BMC Medicine 17, 163 (2019).

[36] Balshem, H. et al. GRADE guidelines: 3. rating the quality of evidence. J. Clin. Epidemiol. 64, 401–406 (2011).

[37] Dekker, M. M., Coffeng, L. E., Pijpers, F. P., Panja, D. & de Vlas, S. J. Reducing societal impacts of sars-cov-2 interventions through subnational implementation. eLife 12, e80819 (2023). URL 10.7554/eLife.80819.

[38] Driscoll, J. C. & Kraay, A. C. Consistent Covariance Matrix Estimation With Spatially Dependent Panel Data. The Review of Economics and Statistics 80, 549–560 (1998). URL https://ideas.repec.org/a/tpr/restat/v80y1998i4p549-560.html.

[39] Hoechle, D. Robust standard errors for panel regressions with cross-sectional dependence. Stata Journal 7, 281–312 (2007). URL https://ideas.repec.org/a/tsj/stataj/v7y2007i3p281-312.html.

[40] Ebisuzaki, W. A Method to Estimate the Statistical Significance of a Correlation When the Data Are Serially Correlated. Journal of Climate 10, 2147–2153 (1997).

[41] Politis, D. N. & Romano, J. P. The stationary bootstrap. Journal of the American Statistical Association 89, 1303–1313 (1994). URL 10.1080/01621459.1994.10476870.

[42] Wooldridge, J. M. Two-way fixed effects, the two-way mundlak regression, and difference-in-differences estimators (2021). URL https://ssrn.com/abstract=3906345.

[43] Harvey, A. State space models, 269–275 (Palgrave Macmillan UK, London, 2010). URL 10.1057/9780230280830_30.

[44] Schwarz, G. Estimating the Dimension of a Model. The Annals of Statistics 6, 461–464 (1978). URL http://www.jstor.org/stable/2958889.

[45] Breiman, L. Random forests. Machine Learning 45, 5–32 (2001).

[46] Zou, H. & Hastie, T. Regularization and variable selection via the elastic net. Journal of the Royal Statistical Society Series B: Statistical Methodology 67, 301–320 (2005). URL 10.1111/j.1467-9868.2005.00503.x.

[47] Hansen, P. C. Truncated Singular Value Decomposition Solutions to Discrete Ill-Posed Problems with Ill-Determined Numerical Rank. SIAM Journal on Scientific and Statistical Computing 11, 503–518 (1990). URL 10.1137/0911028. 10.1137/0911028.

[48] Shapley, L. S. 17. A Value for n-Person Games, 307–318 (Princeton University Press, Princeton, 1953). URL 10.1515/9781400881970-018.

[49] Lundberg, S. M. et al. From local explanations to global understanding with explainable AI for trees. Nature Machine Intelligence 2, 56–67 (2020).

[50] Seabold, S. & Perktold, J. statsmodels: Econometric and statistical modeling with python. In 9th Python in Science Conference (2010).

[51] Pedregosa, F. et al. Scikit-learn: Machine learning in Python. Journal of Machine Learning Research 12, 2825–2830 (2011).

[52] Abbott, B. et al. Analysis of first LIGO science data for stochastic gravitational waves. Phys. Rev. D 69, 122004 (2004). gr-qc/0312088.

[53] Rizzuto, A. C. et al. Zodiacal Exoplanets in Time (ZEIT). VIII. A Two-planet System in Praesepe from K2 Campaign 16. Astron. J. 156, 195 (2018). 1808.07068.

[54] Bundgaard, H. et al. Effectiveness of adding a mask recommendation to other public health measures to prevent SARS-CoV-2 infection in danish mask wearers : A randomized controlled trial. Ann. Intern. Med. 174, 335–343 (2021).

[55] Abaluck, J. et al. Impact of community masking on COVID-19: A cluster-randomized trial in bangladesh. Science 375, eabi9069 (2022).

[56] Jefferson, T. et al. Physical interventions to interrupt or reduce the spread of respiratory viruses. Cochrane Database of Systematic Reviews (2023). URL 10.1002/14651858.CD006207.pub6.

[57] Polack, F. P. et al. Safety and efficacy of the BNT162b2 mRNA covid-19 vaccine. N. Engl. J. Med. 383, 2603–2615 (2020).

[58] Baden, L. R. et al. Efficacy and safety of the mRNA-1273 SARS-CoV-2 vaccine. N. Engl. J. Med. 384, 403–416 (2021).

[59] Chemaitelly, H. et al. mRNA-1273 COVID-19 vaccine effectiveness against the B.1.1.7 and B.1.351 variants and severe COVID-19 disease in Qatar. Nat. Med. 27, 1614–1621 (2021).

[60] Shah, A. S. et al. Effect of Vaccination on Transmission of SARS-CoV-2. New England Journal of Medicine 385, 1718–1720 (2021). URL https://www.nejm.org/doi/full/10.1056/NEJMc2106757. https://www.nejm.org/doi/pdf/10.1056/NEJMc2106757.

[61] Spiliopoulos, L. On the effectiveness of COVID-19 restrictions and lockdowns: Pan metron ariston. BMC Public Health 22, 1842 (2022).

[62] Perra, N., Balcan, D., Gonçalves, B. & Vespignani, A. Towards a characterization of behavior-disease models. PLoS One 6, e23084 (2011).

[63] Agaba, G., Kyrychko, Y. & Blyuss, K. Mathematical model for the impact of awareness on the dynamics of infectious diseases. Mathematical Biosciences 286, 22–30 (2017). URL https://www.sciencedirect.com/science/article/pii/S0025556417300433.

[64] Munro, A. P. S. & House, T. Cycles of susceptibility: Immunity debt explains altered infectious disease dynamics post-pandemic. Clinical Infectious Diseases ciae493 (2024). URL 10.1093/cid/ciae493.

[65] Feng, Z., Towers, S. & Yang, Y. Modeling the effects of vaccination and treatment on pandemic influenza. AAPS J. 13, 427–437 (2011).

[66] Worby, C. J., Wallinga, J., Lipsitch, M. & Goldstein, E. Population effect of influenza vaccination under co-circulation of non-vaccine variants and the case for a bivalent a/h3n2 vaccine component. Epidemics 19, 74–82 (2017). URL https://www.sciencedirect.com/science/article/pii/S1755436517300208.

[67] Backer, J., van Boven, M., van der Hoek, W. & Wallinga, J. Vaccinating children against influenza increases variability in epidemic size. Epidemics 26, 95—103 (2019). URL 10.1016/j.epidem.2018.10.003.

[68] de Boer, P. T., Backer, J. A., van Hoek, A. J. & Wallinga, J. Vaccinating children against influenza: overall cost-effective with potential for undesirable outcomes. BMC Med. 18, 11 (2020).

[69] Dürr, H.-P. & Eichner, M. Corona-Pandemie: Zukunfts-Überlegungen aus der Sicht epidemiologischer Modellierung. Monitor Versorgungsforschung 02/22, 57 (2022).

[70] Castioni, P., Gómez, S., Granell, C. & Arenas, A. Rebound in epidemic control: how misaligned vaccination timing amplifies infection peaks. npj Complexity 1, 20 (2024).

[71] Anderson, R. M. & May, R. M. Vaccination against rubella and measles: quantitative investigations of different policies. J. Hyg. (Lond.) 90, 259–325 (1983).

[72] Cohen, T. & Lipsitch, M. Too little of a good thing: a paradox of moderate infection control. Epidemiology 19, 588–589 (2008).

[73] Heffernan, J. M. & Keeling, M. J. Implications of vaccination and waning immunity. Proc. Biol. Sci. 276, 2071–2080 (2009).

[74] Chiolero, A., Tancredi, S. & Ioannidis, J. P. A. Slow data public health. Eur. J. Epidemiol. 38, 1219–1225 (2023).

