## Supplementary materials (single file) for "Uncertainty and Inconsistency of COVID-19 Non-Pharmaceutical Intervention Effects with Multiple Competitive Statistical Models"

Bernhard Müller
*School of Physics and Astronomy, Monash University, Clayton, VIC 3800, Australia*

Inken Padberg
*Epidemiology Unit, German Rheumatism Research Centre (DRFZ), Charitéplatz 1, 10117, Berlin, Germany*

Michael Lorke
*Faculty of Physics, University of Duisburg-Essen, 47057 Duisburg, Germany*

Ralph Brinks
*Chair for Medical Biometry and Epidemiology Witten/Herdecke University, Faculty of Health/School of Medicine D-58448 Witten, Germany*

Sally Cripps
*Human Technology Institute (HTI), University of Technology Sydney, Sydney, NSW, Australia*

M. Gabriela M. Gomes
*Department of Mathematics and Statistics, University of Strathclyde, Glasgow, United Kingdom and NOVA School of Science and Technology, Centre for Mathematics and Applications (NOVA MATH), Caparica, Portugal.*

Daniel Haake
*Independent Researcher, D-14469 Potsdam, Germany*

John P. A. Ioannidis
*Departments of Medicine, of Epidemiology and Population Health, and of Biomedical Data Science, and Meta-Research Innovation Center at Stanford (METRICS), Stanford University, 3180 Porter Dr, Room A129, Stanford Research Park, Palo Alto, CA 94304, USA*

(Dated: Accepted XXX. Received YYY; in original form ZZZ)

### S1. SUPPLEMENTARY DISCUSSION OF THE BASELINE MODEL

This section provides a detailed supplementary discussion of the methodology and limitations of the baseline model. It includes additional diagnostics for the problems of autocorrelation and multicollinearity and analyses key epidemiological modelling assumptions made by the model. The implications of the epidemiological assumptions and the input data quality will be investigated in Work Packages 2 and 3 of the project.

#### A. Autocorrelation and Multicollinearity

For visually diagnosing (temporal) autocorrelation directly in the residuals instead of merely comparing the fit and the data, we show the fit residuals $\delta \ln \mathcal{R}(t)$ in Supplementary Figure 1.

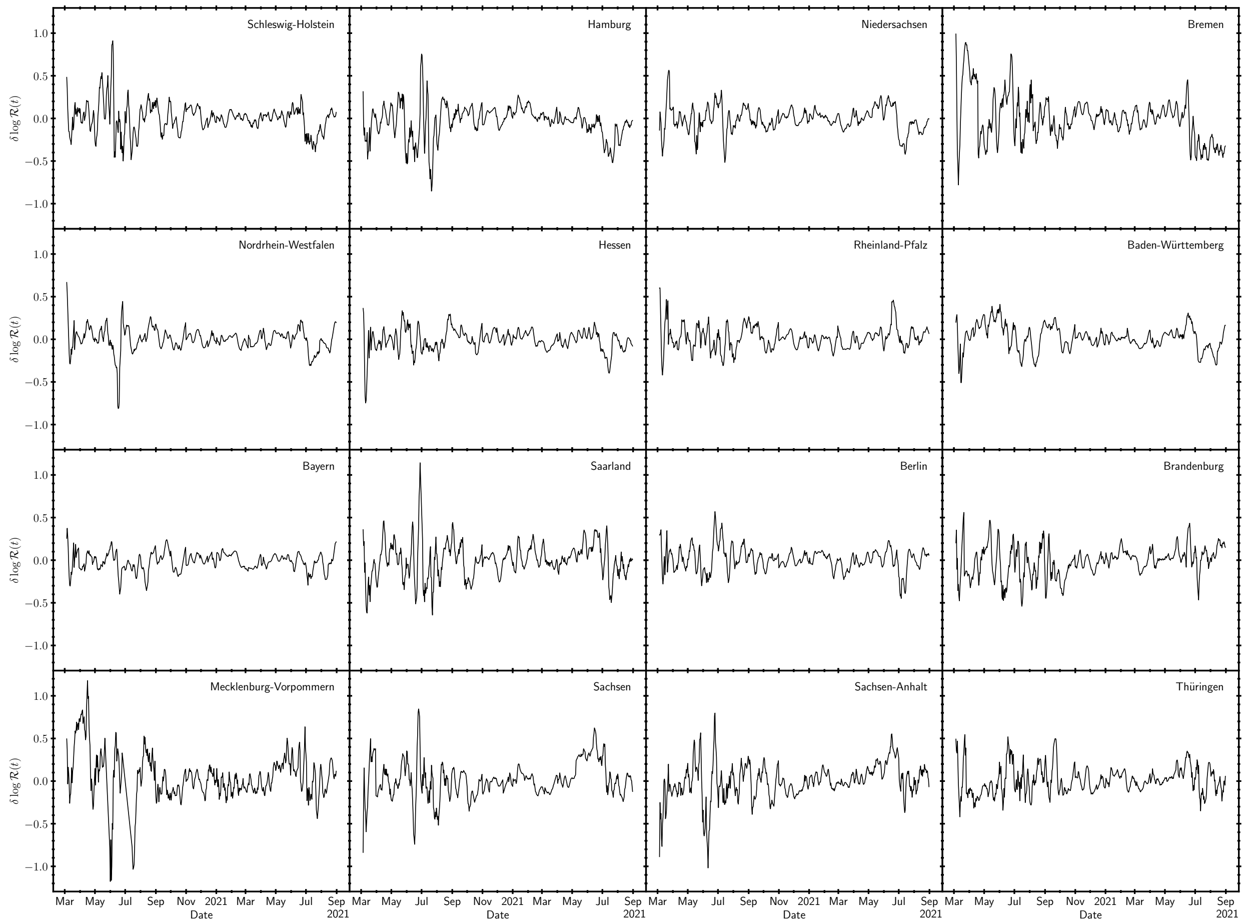


FIG. 1. Fit residuals for the baseline model for all 16 German states. Note the clear presence of autocorrelation, which invalidates the assumption of independent regression errors.

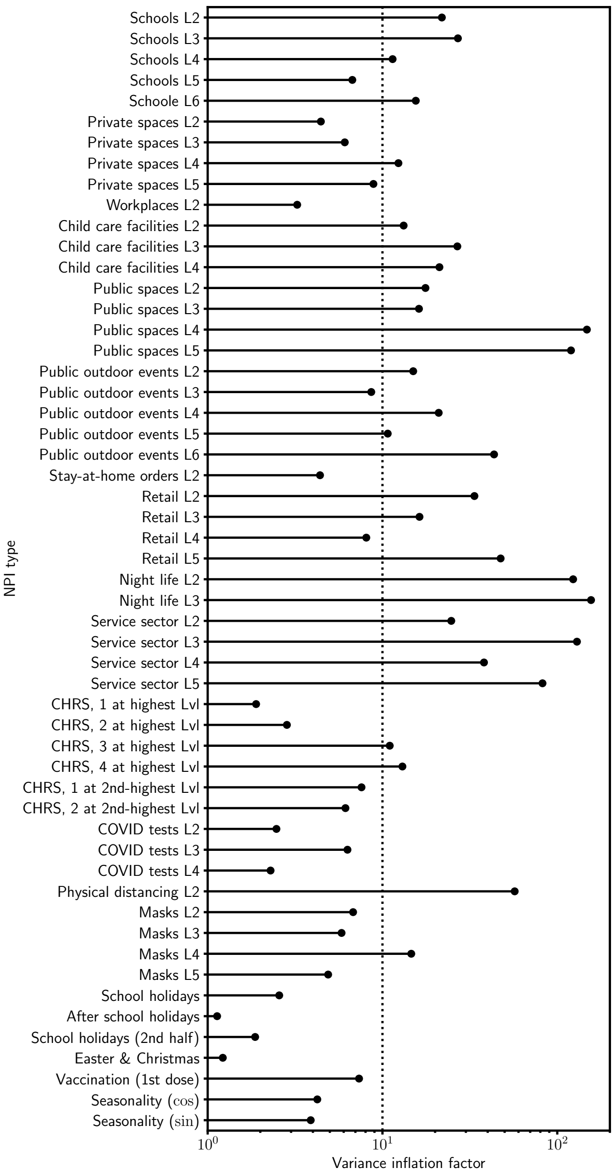


FIG. 2. Variance inflation factors for NPIs and other possible determinants of disease spread considered by *StopptCOVID*. High values indicate strong correlation with other NPIs, which may lead to unstable estimates.

Note that the residuals contain some feature that also appear to be correlated across several states (e.g., a longer phase with $\delta \ln \mathcal{R}(t) < 0$ towards the end of the time series in Schleswig-Holstein, Hamburg, Niedersachsen, Bremen, Nordrhein-Westfalen and Baden-Württemberg), but much of the high- and medium-frequency noise appears to be uncorrelated across states.

As diagnostics for multicollinearity, we show the variance inflation factors for all the explanatory variables in Supplementary Figure 2. The variance inflation factor $\mathrm{VIF}_i$ for the $i$-th explanatory variable is defined as,

$$\mathrm{VIF}_i = \frac{1}{1 - R_i^2}, \tag{1}$$

where $R_i$ is the coefficient of determination for a regression problem for the $i$-th explanatory variable in terms of the other explanatory variables,

$$X_{j,t,i} = \sum_{i' \neq i} \beta'_{i'} X_{j,t,i'}, \tag{2}$$

with regression coefficients $\beta'_{i'}$.

Note that the variance inflation factor appears in the formula for the standard errors of the regression coefficients if $\mathrm{var}\,\beta_j$ is expressed in the form $\mathrm{var}\,\beta_i = s^2 \mathrm{VIF}_i / [(n-1)\mathrm{var}\,X_i]$ in terms of the scatter $s^2$ around the regression surface and the number $n$ of data points [1]. This does *not* mean, however, that the standard formula overestimates $\mathrm{var}\,\beta_i$ and that the actual uncertainty is smaller. The variance inflation factor merely show *how much* multicollinearity *contributes* to the uncertainty.

Another possible issue with times series is non-stationarity, which can give rise to spurious correlations when applying standard regression techniques. The time series for $\mathcal{R}_t$ exhibits mild non-stationarity, especially in the form of the decrease of $\mathcal{R}_t$ at the beginning of pandemic. However, this is arguably well accounted for by the model. Stationarity of the residuals has been confirmed by an augmented Dickey-Fuller test for reach state [2]; non-stationarity of the residuals is consistently rejected at $p$-values $\ll 0.05$. The degree of non-stationarity of the covariates and the response variable $\mathcal{R}_t$ is arguably limited; The *NPI* variables are confined to the interval $[0, 1]$. $\mathcal{R}_t$ always remains of order unity after the initial phase, and can never drift to arbitrary large values for biological reasons. More complex tests for co-integration of non-stationary panel data [3, 4] were not deemed necessary at this point; a bigger priority will be to properly model factors that affect the epidemiology of early disease spread (clustering and heterogeneity) that contributes to the early non-stationarity.

#### B. Computation of the Effective Reproduction Number

Equation (3) intends to approximate $\mathcal{R}_t$ for the idealised case of a generation time of *exactly* 4 d with no individual variations in the delay between the primary and secondary cases in an infection chain,

$$\mathcal{R}_t = \frac{\mathcal{I}_t}{\mathcal{I}_{t-4}}. \tag{3}$$

However, in adopting this definition, *StopptCOVID* implicitly assumes that the incubation period is zero; in reality the mean delay between infection and symptom onset is several days [5, 6]. $\mathcal{R}_t$ as computed from Equation (3) therefore lags the true instantaneous reproduction number, i.e., it approximates the true $\mathcal{R}_t$ several days prior to time $t$. The assumption of a negative $\tau_{\mathrm{NPI}}$ between NPIs and their effect on $\mathcal{R}_t$ thus becomes even more problematic; effectively the baseline model of *StopptCOVID* assumes an effect *several* days before any NPI is switched on.

Worse yet, one-sided smoothing in Equation (3) introduces further bias. If indeed $\mathcal{I}_t = \mathcal{R}_t \mathcal{I}_{t-4}$, then Equation (3) approximates a weighted average of $\mathcal{R}_t$ for the last seven days before and including time $t$,

$$\frac{\sum_{\tau=0}^{6} \mathcal{I}_{t-\tau}}{\sum_{\tau=4}^{10} \mathcal{I}_{t-\tau}} = \frac{\sum_{\tau=0}^{6} \mathcal{R}_{t-\tau} \mathcal{I}_{t-\tau}}{\sum_{\tau=0}^{6} \mathcal{I}_{t-\tau-4}}. \tag{4}$$

For further discussion of this issue, see Section S7 B. From a rigorous perspective, the determination of $\mathcal{R}_t$ holds even more subtleties, as it is tied to the time-dependence of the infectivity function and involves the inversion of a Fredholm-type integral equation, see [7] and Section S7. The non-unicity of this inverse problem introduces additional uncertainties not related to measurements uncertainties and requires (explicit or implicit) regularisation.

Although $\mathcal{R}_t$ as used in *StopptCOVID* thus considerably lags the true $\mathcal{R}_t$, we accept their values as response variable for the purpose of this study. A more rigorous approach for calculating $\mathcal{R}_t$ will be used in Work Package 2. It is desirable to not only correct for the lag between infection and symptom onset and use an unbiased method to calculate $\mathcal{R}_t$, but to also take the time dependence of the transmission probability into account [8] as this has a non-negligible impact on epidemic dynamics [e.g., 9–11]. It is therefore natural to solve the problem of spurious lags together with a generalisation of epidemiological modelling assumptions in Work Package 2.

Regarding $\mathcal{R}(t)$, it is also important to critically examine the issue of data quality. The case data underlying the calculation of $\mathcal{R}(t)$ are not based on a representative sample, and not based on uniform, time-independent testing and diagnostic criteria. For Germany, no representative, longitudinal study like the ONS infection survey [12] is available to either directly provide better data or allow calibration of the official case numbers. If the ratio $\eta_{\mathrm{test}} = \mathcal{I}/\mathcal{I}_{\mathrm{true}}$ of detected cases $\mathcal{I}$ and true cases $\mathcal{I}_{\mathrm{true}}$ changes due to improper calibration of the official case numbers, this leads to a bias in $\ln \mathcal{R}$ of about $\delta \ln \mathcal{R} \approx \tau \, \mathrm{d} \ln \eta_{\mathrm{test}} / \mathrm{d}t$ and will impact effect estimates. Strong gradients in test numbers and the detection probability are therefore most problematic. In the context of this study, this leads to two concerns in particular. First, there was a steep ramp-up of testing during

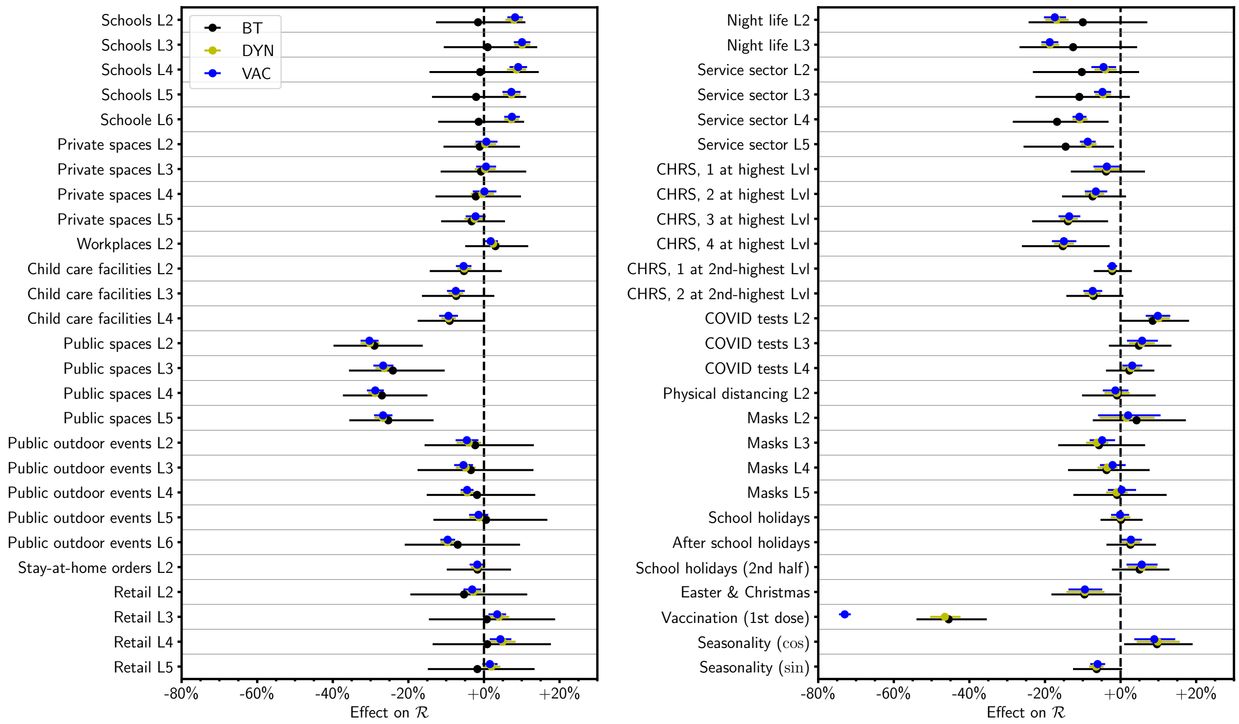


FIG. 3. Effect sizes with the corrected implementation of the vaccine effect compared to models BT and DYN with linearisation in $\ln(1 - V)$ as in the baseline model. Note that the vaccine effect shown in this figure is $1 - \eta_{\text{vac}}$, whereas Figures 2–5 show the vaccine effect on $\mathcal{R}$ for $V = 0.5$ in the linear approximation. Note also the different scale compared to the previous figures.

March 2020, which further adds to the challenge of determining the effects of early interventions. Second, compulsory rapid antigen tests (and PCR follow-up for positives) in schools were widely introduced in spring 2021 also for asymptomatic students. This potentially introduces bias around school holidays as the testing frequency of students changes abruptly. A detailed analysis of possible implications of data artefacts will be left to Work Package 3 of the project.

#### C. Implementation of Vaccine Effect

The implementation of vaccine efficacy in Equation (1) warrants scrutiny. *StopptCOVID* has chosen the particular form of this term to capture the effect of halving the fraction of unvaccinated individuals on $\mathcal{R}$. This interpretation would be perfectly reasonable if the vaccine effect on $\ln \mathcal{R}$ were linear in $\ln(1 - V)$. As pointed out in *StopptCOVID*, perfect vaccine efficacy of 100% would then imply a reduction of $\mathcal{R}$ by 50%, i.e., the regression coefficient cannot be identified with the standard vaccine efficacy, which would be desirable for its interpretation.

The logarithmic dependence of vaccine efficacy *could* be motivated in an idealised picture where new infections occur *exactly* one generation time after the previous "round" of infections. For perfect efficacy against transmission, vaccination will then decrease the reproduction number by a factor given by the fraction of unvaccinated individuals, $\mathcal{R} \to \mathcal{R}(1-V)$. This, however, would imply a reduction of $\ln\mathcal{R}$ by $\ln(1-V)$. One may then be tempted to incorporate imperfect vaccine efficacy as regression coefficient for this term. Using $\log_2(1-V)$ instead of $\ln(1-V)$ as an explanatory variable would simply amount to rescaling this variable. The vaccine efficacy $\eta_{\mathrm{vac}}$ can be obtained from $\beta_2$ in Equation (1) as $\eta_{\mathrm{vac}} = \beta_2/\ln 2$.

While this implementation of the vaccine effect may seem intuitive, it suffers from a serious problem, which becomes evident when it is derived more formally. Let $\mathcal{I}'$ and $\mathcal{I}$ be the number of new infections one generation time apart, and let $\hat{\mathcal{R}}$ be the reproduction number without vaccination, i.e., $\mathcal{I}' = \hat{\mathcal{R}}\mathcal{I}$. With vaccination, the number $\mathcal{I}'$ of new cases after a generation time becomes $\mathcal{I}' = [(1-V) + (1-\eta_{\mathrm{vac}})V]\hat{\mathcal{R}}\mathcal{I}$, since the risk of infection is reduced by a factor $1-\eta_{\mathrm{vac}}$ for vaccinated individuals. Hence

$$\ln\mathcal{R} = \ln\hat{\mathcal{R}} + \ln\left[(1-V) + (1-\eta_{\mathrm{vac}})V\right]. \tag{5}$$

By Taylor series expansion in $\ln(1-V)$, one can formally derive a vaccine effect of the form used in *StopptCOVID*. The actual vaccine effect on $\ln\mathcal{R}$ is generally *smaller* than $\eta_{\mathrm{vac}}|\ln(1-V)|$, and is also non-linear. Parameter inference based on Equation (1) therefore risks to underestimate $\eta_{\mathrm{vac}}$ (if we ignore other biases in the model). What is even more problematic, a vaccine effect of the form $\eta_{\mathrm{vac}}\ln(1-V)$ (or $\eta_{\mathrm{vac}}\log_2(1-V)$) gives the wrong limit for $V=0$, i.e., $\ln\mathcal{R}\to-\infty$ or $\mathcal{R}\to 0$ *regardless of vaccine efficacy*.

Note that the correct equation (5) *cannot* be implemented in a linear regression model; a general additive model (GAM; 13) would be required. Worse, if epidemic growth is not simplified to discrete steps over exactly one generation time, the vaccination effect involves both $V$ and $\ln\hat{\mathcal{R}}$; in a SIR-type model with vaccination and no depletion of susceptibles by infection, the vaccine effect becomes $\Delta\ln\mathcal{R} = -V\eta\left(1+\ln\hat{\mathcal{R}}\right)$. Fortunately, the correct multiplier for $\mathcal{R}$ can be conveniently included in a renewal equation as a variation of model DYN.

Since this is a highly important issue that concerns the interpretation of an estimated effect size, we here present effect estimates and confidence intervals for this modified model (model VAC), even though this entails a modification of the original epidemiological assumption. However, this modified model still does not consider multiple doses and waning of vaccine efficacy.

Model VAC is formulated in terms of the vaccine efficacy $\eta_{\mathrm{vac}}$ as

$$\begin{aligned}\bar{\mathcal{I}}_{j,t} = \mathcal{R}_{j,t}\bar{\mathcal{I}}_{j,t-4} = \Big[(1 - V_{j,t-\tau_{\mathrm{vac}}}) + (1 - \eta_{\mathrm{vac}})V_{j,t-\tau_{\mathrm{vac}}}\Big] \\ \times \exp\Bigg[\alpha_j + 0.3\nu_{\alpha,t} + 0.6\nu_{\delta,t} + \beta_0 \cos\frac{2\pi t}{365\,\mathrm{d}} + \beta_1 \sin\frac{2\pi t}{365\,\mathrm{d}} + \sum_{i=3}^{N_{\mathrm{NPI}}+2} \beta_i X_{j,i}(t)(t-\tau_{\mathrm{NPI}})\Bigg]\mathcal{I}_{j,t-4}.\end{aligned} \tag{6}$$

The implementation as a state space model proceeds analogously to model DYN. The same implementation of a stationary bootstrap is used. The changes required in the code are minor and limited to a few lines for data preparation and the setup of the transition matrix.

Results for the corrected treatment of the vaccine effect in model VAC are shown in Figure 3. It is important to revisit *all* of the effect estimates, as these may also be impacted by the different treatment of vaccination. In practice, the changes to the other effect sizes are small and remain with the BT error bars.

Figure 3 compares the vaccine effect estimated by the corrected model VAC to the estimate by models DYN and BT. The figure shows $1-\eta_{\mathrm{vac}}$, i.e., the factor by which the probability of infection and subsequent transmission is reduced for vaccinated individuals. Note that for a meaningful comparison, we show the coefficient $\eta_{\mathrm{vac}}$ in the linear approximation $\Delta \ln \mathcal{R} = \eta_{\mathrm{vac}} \ln(1 - V))$ for models DYN and BT rather than the coefficient $\beta_3 = \ln 2\eta_{\mathrm{vac}}$ in Equation (1). Figure 3 therefore shows different values for models DYN and BT than Figures 2–5 due to this rescaling.

The implementation of vaccination in *StopptCOVID* also suffers from other limitations. It only incorporates first-dose vaccination even though the second dose is known to substantially increase vaccine efficacy against infection [14, 15]. Neither does it incorporate vaccine waning and boosting, which are relevant to epidemic dynamics on time scales of months [15, 16]. Nevertheless, these limitations are accepted in this study and left to further investigation in Work Package 2.

### D. Further Discussion of Model Assumptions and Limitations

#### *Treatment of Age Structure*

*StopptCOVID* runs the regression model (1) both on the time series for $\mathcal{R}_{j,t}$ for each state as a whole and on an $\mathcal{R}_{j,t}$ computed separately for three different age groups (0-17 years, 18-59 years, 60 years or older) based on age-stratified case data.

Age structure is generally relevant to epidemic dynamics, and should therefore be accounted for in estimating NPI effects; neglecting it by assuming a homogenous population may lead to biased estimates. However, simply computing separate effective reproduction numbers $\mathcal{R}_{j,t}$ for each age group is a problematic approach. The rigorous approach is rather to promote the growth rate to a matrix; e.g., Equation (3) would become,

$$\boldsymbol{\mathcal{I}}_t = \boldsymbol{\mathcal{R}}_t \cdot \boldsymbol{\mathcal{I}}_{t-4}, \tag{7}$$

with matrices $\boldsymbol{\mathcal{I}}$ and $\boldsymbol{\mathcal{R}}$. One then has the choice of estimating *all* of the coefficients of the matrix $\boldsymbol{\mathcal{R}}$, or of specifying the components of $\boldsymbol{\mathcal{R}}$ *without* interventions and incorporating NPI effects on specific age groups (or the transmission between age groups) as perturbations of rows, columns, or individual elements of this matrix. The transmission matrix without interventions could, e.g., be constructed based on POLYMOD [17] contact data and calibrated such as to achieve the desired (scalar) value of $\mathcal{R}_0$.

The approach taken by *StopptCOVID* implicitly assumes that the contact matrix is perfectly diagonal. The pre-pandemic contact patterns are dominated by the diagonal terms; but the off-diagonal terms between the three aforementioned age groups are sizeable for household contacts. It is thus far from obvious that the non-diagonal terms can be neglected when estimating age-stratified NPI effects, especially when contact patterns and frequencies have shifted considerably during the pandemic. Moreover, the amount of transmission between age groups does not depend *only* on the transmission matrix, but also on the age-stratified fraction of infectives, so that off-diagonal terms can become relevant even when they are small compared to the diagonal terms.

Because of such complications, we decided to *only* consider the population-averaged effect of NPIs on $\mathcal{R}_t$ for all age groups in the present study. Rather than adopting the original *StopptCOVID* treatment of age dependence, we defer this issue to Work Package 2, where it will be treated with greater rigour.

*Other Heterogeneity Effects and Past Infections*

Aside from a crude treatment of age dependence, *StopptCOVID* does not take into account any other effects of heterogeneous epidemic dynamics. Individual variations, e.g., in susceptibility and infectivity, contact rates as well as the detailed structure of infection networks (clustering, degree of assortativity, detailed spatial dynamics) are not accounted for. In fact, vaccination is treated as the only effect that influences susceptibility (Section S1 C); depletion of susceptibles by prior infection is ignored, as is waning of immunity.

It has long been known even before the pandemic that heterogeneity can affect early epidemic dynamics, the size of outbreaks and the fraction of susceptibles in endemic equilibrium [e.g., 18–27]. Although heterogeneity has often been neglected in epidemic modelling during the pandemic, its importance for epidemic dynamics in the context of COVID-19 has also been discussed prominently.

In fact, *StopptCOVID does* appeal to cluster effects among the unvaccinated population for justifying that the effect of vaccination deviates from the authors' expectation, but without quantitatively modelling that purported effect. An early modelling scenario for COVID-19 by Germany's RKI also acknowledged cluster effects as an uncertainty by including an alternative scenario with a reduced fraction of effective susceptibles in the population of $2/3$ instead of 1 [28].

Importantly, in the presence of heterogeneity, it can no longer be safely assumed that past infections simply decrease $\mathcal{R}$ by a factor $1 - I - R$ (where $I$ and $R$ are the fraction of current infectives and recovered individuals) as in simple SIR models without substructure. Even ignoring heterogeneity effects *and* undetected infections, the factor $1-I-R$ would already lead to a decrease of $\mathcal{R}$ by slightly less than 5% by the end of the period of interest, comparable to many of the NPI effects estimated by *StopptCOVID*. Neglecting heterogeneity and past infections in the estimation of NPI effects is therefore problematic. These factors will be addressed in Work Package 2; we refrain from a superficial investigation here.

#### *Effect Delay of NPIs*

The original model allows negative delays, which violates causality. *StopptCOVID* justifies this negative delay based on the notion that the population anticipates NPIs. While this explanation may appear plausible, this remain an ad-hoc hypothesis within the model, and would need to be properly justified, and the semantic interpretation of effects would have to be adjusted in the sense that they represent the adherence to specific NPIs and not the effects of policies. Furthermore that adherence would need to be measured and included in the model in the first place. This may be possible for some NPIs, e.g., using properly calibrated contact data. For some NPIs, independent measurements of adherence may not be trivial, or negative delays may not possibly be justified (e.g., for testing policies that come into force exactly on the specified date). The imposition of a non-negative delay would not change results drastically under the current model assumptions (especially without improvements to the estimation of $\mathcal{R}_t$). Within *StopptCOVID*, a sensitivity analysis yielded only small changes of a few 0.01 in the regression coefficients for a delay of 0 d instead of −1 d. We also considered delays of 0 d, −1 d and −2 d for our model with ARMA($p, q$) errors. The delay of −1 d still minimises the Akaike information criterion for this model, and the maximum difference in the regression coefficients is only 0.04. For this reason, the effect delay is not investigated further in this study, but will be considered in the next work packages.

Similarly, for the effect of vaccination it would be preferable to incorporate the time dependence of vaccine efficacy after the first, second, and subsequent doses more realistically. In principle, the data for vaccine efficacy could be specified, e.g., as Bayesian priors, based on epidemiological data from studies like the ONS infection survey [12].

However, in the spirit of keeping as close to the original model as is viable in Work Package 1, these issues are not further explored in this study. We even keep these delay parameters *fixed* to the optimal values from the baseline model when we use alternative approaches. The rationale for not re-estimating the delays is that the error analysis would otherwise become substantially more complicated; in particular, some of the simple plug-in estimators in statistical analysis packages would no longer be applicable without modification.

*Further Limitations*

There are further epidemiological assumptions in *StopptCOVID* that may lead to biased effect estimates for NPIs, and ought to be investigated by extending the model in Work Package 2. The problem of disentangling the effects of NPIs from self-regulated behavioural adaptations has already been alluded to in the main discussion section, and may be investigated by recourse to contact data. The treatment of variants definitely requires improvement. Even though the imposed variant effects are in the ballpark of estimates of $\mathcal{R}_0$ for these variants [29, 30], simply specifying the effect of variants without a sensitivity analysis or direct estimation of the variant effects is not satisfactory. Other factors, such as stochastic dynamics at low case numbers or import of cases may be less problematic, as the low-case number regime has less influence on the effect estimates of *StopptCOVID*.

By including fixed effects $\alpha_j$ in Equation (1), the *StopptCOVID* model can *partly* absorb the effects of the neglected factors on disease spread. Effectively, fixed effects amount to a renormal-

TABLE I. Basic reproduction number $\mathcal{R}_0$ without NPIs from the estimated fixed effects for the 16 German states.

| State | $\mathcal{R}_0$ |
|---|---|
| Schleswig-Holstein | 2.39 |
| Hamburg | 2.35 |
| Niedersachsen | 2.24 |
| Bremen | 2.09 |
| Nordrhein-Westfalen | 2.30 |
| Hessen | 2.43 |
| Rheinland-Pfalz | 2.48 |
| Baden-Württemberg | 2.43 |
| Bayern | 2.41 |
| Saarland | 2.34 |
| Berlin | 2.27 |
| Brandenburg | 2.18 |
| Mecklenburg-Vorpommern | 2.23 |
| Sachsen | 2.28 |
| Sachsen-Anhalt | 2.31 |
| Thüringen | 2.22 |

isation of the basic reproduction number for the wild type. The seasonally-averaged values of the inferred basic reproduction number $\mathcal{R}_0 = \exp \alpha_j$ based on the estimated fixed effects are shown in Supplementary Table I. These are roughly consistent with other estimates of $\mathcal{R}_0$ for the wild type [31, 32].

This consistency does not mean that the model is already correctly specified, however. Merely lumping unmodelled effects into fixed effects for states (or, alternatively, random effects) is *not* sufficient for unbiased effect estimates. For example, entity fixed or random effects for $\mathcal{R}$ will generally not capture cluster effects, if cluster effects introduce time-dependent dynamics in $\mathcal{R}$. Even more sophisticated data analysis techniques (trend subtraction, etc.) are not guaranteed to remove systematic errors completely, and some may also unintentionally introduce a bias toward the null. Ultimately, the correction of systematic errors remains a problem of epidemiology instead of statistics, and needs to proceed by appropriately modelling the relevant epidemiological effects and estimating them along with NPI effects or constrain them by independent data.

Finally, the inferred NPI "effects" remain, strictly speaking, only statistical associations. and do not demonstrate causality. For example, feedback of epidemic dynamics on NPIs opens the possibility of reverse causation and is not included in the model, although *StopptCOVID* acknowledges the possibility of such feedback. Establishing causality and the mechanisms behind variations in $\mathcal{R}(t)$ is a complex problem, but this is a moot point – and hence not addressed in this paper – until statistical associations can be established in the first place.

TABLE II: Description of NPIs and stringency levels. Translated and adapted from Table 1 of the *StopptCOVID* study [33].

| Code | Explanation |
|---|---|
| Schools L2 | Primary schools fully open with COVID safety regulations |
| Schools L3 | Restricted opening of primary schools, e.g., shift operations, with COVID safety regulations |
| Schools L4 | Selective opening (e.g., by year or by subject) |
| Schools L5 | Primary schools closed |
| Schools L6 | Primary and secondary schools closed |
| Private spaces L2 | Recommendation to avoid contacts |
| Private spaces L3 | Upper limit of 20-100 persons for private gatherings |
| Private spaces L4 | Upper limit of 5-10 persons |
| Private spaces L5 | Upper limit of 2 households or less |
| Workplaces L2 | Recommendation to work from home + COVID safety regulations or partial closure |
| Child care facilities L2 | Open with COVID safety regulations |
| Child care facilities L3 | Restricted operation |
| Child care facilities L4 | Emergency care only or closure |
| Pubic spaces L2 | Recommendation to avoid contacts |
| Pubic spaces L3 | Upper limit of 20-100 persons for public gatherings |
| Pubic spaces L4 | Upper limit of 5-10 persons |
| Pubic spaces L5 | Upper limit of 2 households or less |
| Public outdoor events L2 | Upper limit of 1000-5000 persons |
| Public outdoor events L3 | Uper limit of 500-700 persons |
| Public outdoor events L4 | Upper limit of 100-400 persons |
| Public outdoor events L5 | Upper limit of 10-50 person |
| Public outdoor events L6 | Prohibition of events |
| Stay-at-home orders L2 | Recommendation to stay at home, stay-at-home order with or without exemptions |
| Retail L2 | Open with COVID safety regulations |

Description of explanatory variables (*continued*)

| Code | Explanation |
| --- | --- |
| Retail L3 | Restricted opening with COVID safety regulations |
| Retail L4 | Venues above 700-800 sqm closed or restricted opening hours |
| Retail L5 | Closed with possible exemption for critical supplies or other products |
| Night life L2 | COVID safety regulations (e.g. no indoor dancing) and/or restricted opening and/or partial closures (discothèques) |
| Night life L3 | Venues closed |
| Service sector L2 | Open with COVID safety regulations |
| Service sector L3 | Restricted opening with COVID safety regulations and/or closure of brothels |
| Service sector L4 | Closure in case of close contact with customers (e.g., hairdressers) |
| Service sector L5 | Complete closure |
| CHRS | Combined category depending on the activation of the highest levels of restrictions for the cultural sector, hotels, restaurants, and sports |
| Cultural sector L4 | Restriction to outside activities, sale of food & beverages, or safety restrictions for museums |
| Cultural sector L5 | Complete closure |
| Hotels L2 | COVID safety restrictions apply |
| Hotels L3 | No overnight accommodation or complete closure |
| Restaurants L4 | Outdoor dining or take-home orders only |
| Restaurants L5 | Closed or take-home orders only |
| Sports L4 | No indoor sports, outdoor facilities open with or without restrictions |
| Sports L5 | No indoor sports, outdoor facilities closed, individual outdoor sports permitted |
| COVID tests L2 | Mandatory testing in case of symptoms or suspected infection, or for essential workers |
| COVID tests L3 | Mandatory testing for public events or in school |
| COVID tests L4 | Mandatory testing upon return from high-risk countries, non-EU countries, or any foreign country |

Description of explanatory variables (*continued*)

| Code | Explanation |
|---|---|
| Physical distancing L2 | Mandatory physical distancing |
| Masks L2 | Mandatory masking on public transport or in shops |
| Masks L3 | Mandatory masking in public spaces |
| Masks L4 | Mandatory masking in secondary and/or primary schools, whether inside the classroom or generally |
| Masks L5 | Penalties for violations of mask mandates |
| School holidays | Dates differ between federal states |
| After school holidays | Period of 5 days after school holidays |
| School holidays (2nd half) | Second half of school holidays, if holidays last for at least 12 days |
| Easter/Christmas | Good Friday to Easter Monday, 24 December to 31 December |

### S2. SUMMARY OF LITERATURE REVIEW AND MODEL SELECTION

For pragmatic reasons, the literature review was based on the corpus assembled in the Royal Society's review of NPI effectiveness [34]. The literature review began with a preliminary selection of studies that use $\mathcal{R}(t)$ as an outcome, included multiple interventions, for which GRADE classification was at least "low" (i.e., not "very low") and that were directed to evaluating the effectiveness of school closures, stay-at-home-orders and mass gatherings. In addition, another 30 studies listed by [34] were selected randomly. Methods used in these studies were then briefly summarised, including whether or not a mathematical description of the methods was given.

These studies provided a first overview over the general techniques that were used for time series analysis of COVID NPI effectiveness. By extracting the common methodological features in these studies, we developed a classification scheme for a full literature review:

1. Multiple linear regression models (largest group of about 120 studies, which included, e.g., models with fixed effects in time, random effects models, autoregressive and vector autoregressive models, general additive models),

2. Machine learning models (e.g., random forest and support vector machines),

3. Models including trend/level changes for a few NPIs (e.g., segmented regression, interrupted time series),

4. Dynamical models (e.g., renewal equations and SIR and related models that can capture non-linear dynamics).

These categories were supplemented by three key dimensions of the model and data structure used in the studies:

1. Model took into account spatial coupling (Yes/No),

2. Based on Bayesian statistics (Yes/No),

3. Time-dependent response function (Yes/No), where $\mathcal{R}(t)$ depends on the past history of explanatory variables and possibly $\mathcal{R}(t)$ itself, e.g., (vector) autoregressive models or models that stratify the data by time since NPIs were implemented.

Next, we selected a subset of studies that could be classified according to this classification scheme. We initially excluded case reports, case series, contact surveys, cohort studies, and randomized controlled trials. As the preliminary round of screening (see above) also included cohort studies, all cohort studies were subsequently included, and only case reports, case series, contact surveys and randomized controlled trials were excluded (see Figure 4). Thus, another 18 additional cohort studies were included in the breakdown below. Whether or not the cohort studies are included does not affect the general picture of statistical methods for panel data in the literature. Studies that were found eligible for classification and accessible ($n = 331$) were then either categorized according to the above scheme, or discarded for classification for one of the following reasons:

1. The model structure and assumptions were too simplistic for setting with multiple NPIs, or not applicable to time series. This included studies which, e.g., considered only certain parameters (e.g., peak or cumulative infections) of the epidemic curve.

2. The description was insufficiently clear. This included some studies where no mathematical and no clear verbal description of the methods was provided.

3. The study was only simulation-based, and did not attempt to fit a model to real-world data.

4. The study provided only a descriptive or correlation/ temporal association between increase/decrease in transmission (or similar outcomes) only. This includes, e.g., studies that simply quantified correlations of outcomes such as total case numbers with NPIs.

Most (191) out of the 331 reviewed papers fit these categories. 140 of the studies were deemed not applicable because the methods were either not (well) explained, because a study was purely descriptive in nature, or because it did not perform statistical inference on time series or panel data (e.g, simulation studies or observational epidemiological studies in specific subsettings such as schools). Two of the studies could not be accessed.

These results from the review are meant to provide a qualitative summary with some additional rough quantitative information, recognising the limitations that the literature review faced: The dimensions of modelling approaches that we considered were not standard reporting items, and classification was often difficult with quite a few edge cases. The most relevant aspects for the current study, i.e., the treatment of errors and multicollinearity, was frequently opaque so that we could not systematically classify the reviewed corpus.

The different technical approaches described above naturally included a vast array of methods each with distinct advantages and disadvantages, which we explain in more detail in Sections S3–S5.

In summary, our decision to select methods for implementation in the current project was based on the following considerations:

1. In the current phase of the validation project, epidemiological model assumptions of the *StopptCOVID* study should be altered as little as possible, and methods should more or less be applicable to the dataset used in the *StopptCOVID* study as is. For this reason, including methods with trend/level changes was not attempted since these are best suited to deal with specific NPIs and can hardly be adapted to about 50 interventions irregular activation patterns as required for *StopptCOVID*. Reducing the number of NPIs examined in the study to fit the technical requirements for these methods would represent a major alterations of the epidemiological model assumptions. For a similar reason, we decided against methods including a time-dependent response function or dynamical methods which include non-linear terms. Implementation of a time-dependent response function using autoregressive terms for the response variable or non-linear terms in dynamical models would also represent major alterations of the epidemiological model assumptions.

2. Model implementation should not rely heavily on any additional data (e.g., knowledge needed for choice of informative priors for Bayesian approaches, or data for spatial coupling matrices) at this stage of the project. Such additional information could be highly uncertain, especially for COVID-19 NPIs where high-level intervention studies such as randomized controlled trails or large observational studies collecting the required data are rare. If model outcomes and final effect estimates largely depended on such additional knowledge, this would make a clean comparison of the effects across the different methods difficult. Consequently, Bayesian methods with informative priors and spatial coupling were also not included in the set of methods implemented at the current stage.

3. The selected methods should be representative of common approaches for NPI effect estimation that were found in the literature, and the multi-model comparison should include sufficiently distinct methods to test the robustness and sensitivity of effect estimates to methodological differences.

4. The model set used should effectively address the previously identified problems for error estimation and multicollinearity to provide tangible improvements over the model of the original *StopptCOVID* study.

5. The selected methods must remain computationally feasible, efficient, readily implementable for the *StopptCOVID* data set, and readily testable. For example, a Bayesian version of an ARMA model using `statsmodels.statespace` and `pymc` was considered, but ran too slowly for efficient testing and production runs. Though this is not a specific drawback of Bayesian models in general and could be fixed by optimisation, the potential gains (e.g., for regularisation) were deemed not to be commensurate with the computational costs without further optimisation.

This left the categories of multiple linear regression, machine learning methods, and dynamic models without non-linear terms. Several suitable and representative variations of these methods were selected. We decided to implement the original linear regression model of the *StopptCOVID* study, supplemented by three alternative approaches for error estimation with autocorrelated noise in linear regression, namely bootstrapping, Driscoll-Kraay and Ebisuzaki's method. We also implemented a linear two-way fixed effect regression model with time-series bootstrapping and regression with ARMA errors. Two additional linear regression-based methods (elastic net and principal component regression) were implemented to address the problem of multicollinearity, with confidence intervals obtained by a time-series bootstrap. Finally, as an examples for a machine learning method, we implemented random forest regression, and a renewal equation model was included as an example for a dynamical model. Both of these were again combined with a time series bootstrap. More details on these methods and reasons for deciding in favour or against their implementation can be found in the following sections.

We noted that details of the analysis methods could often not be gleaned readily from the abstract or from keyword searches in the reviewed papers. In many of the studies, the description of the methodology is cursory and purely verbal. Few of the papers discuss alternative or additional sources of variation that are not considered in their chosen model – such as spatial dependencies, heterogeneity or seasonality. The cursory presentation of the methodology in many studies implies that classical methods for systematic literature reviews face significant difficulties in assessing the type and quality of modelling used to estimate NPI effects.

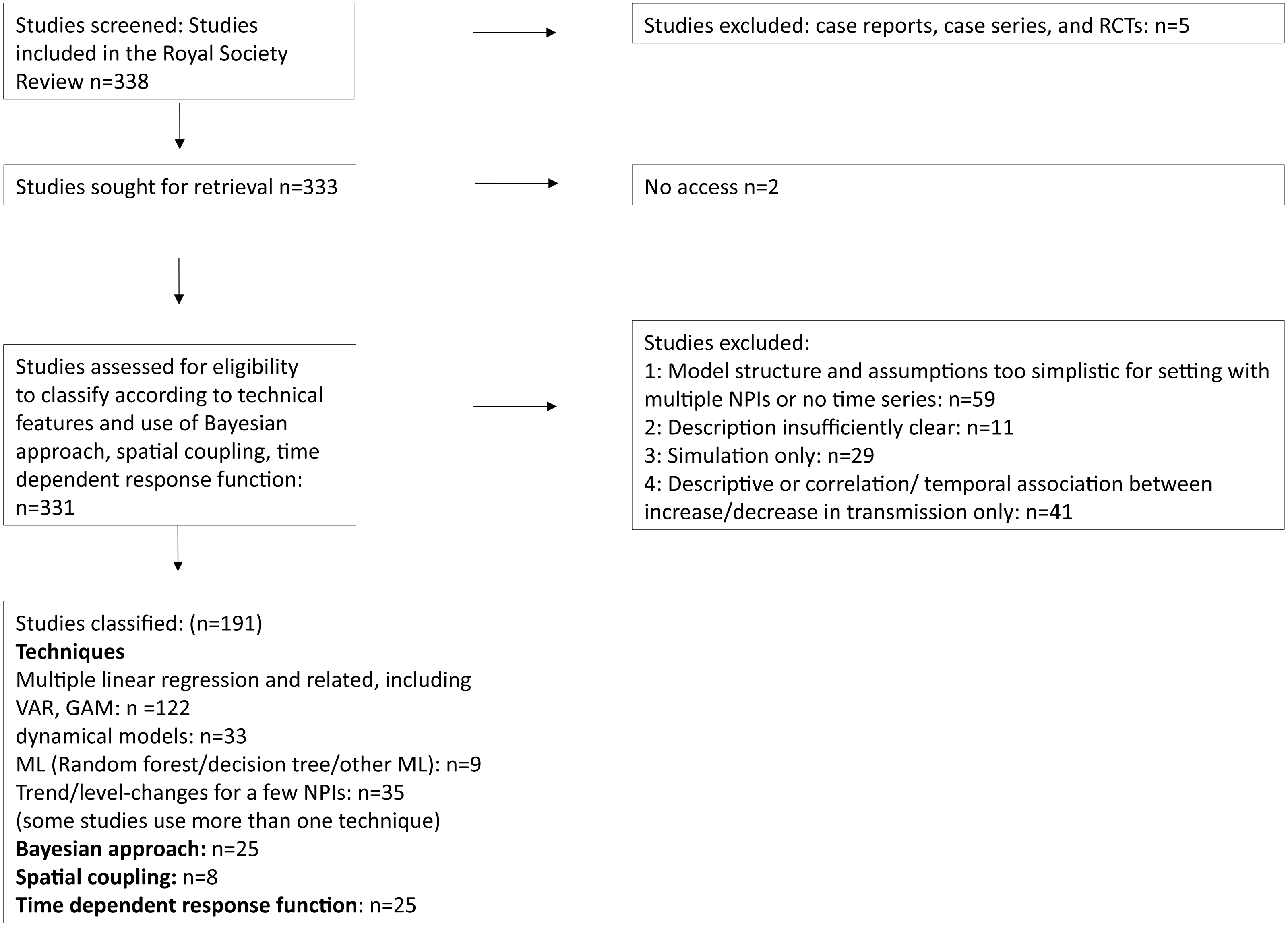


FIG. 4. Flowchart of the study selection and classification process in the literature review.

### S3. METHODS FOR NPI EFFECT ESTIMATION

Based on the analysis of NPI studies referenced in the review of [34] and a broader survey of the literature, we here present a more detailed breakdown of key characteristics and dimensions of models for estimating NPI effect sizes. We concisely describe the employed mathematical techniques, outline advantages and drawbacks, comment on the use of the various methods in NPI studies, and justify why certain methods were included in the model set or not.

#### A. Model Types

*Multiple Linear Regression and Related Models*

The first category of studies comprises multiple linear regression models or generalisations thereof, and also includes the original *StopptCOVID* model. Models within this class therefore share structural similarities with that from *StopptCOVID*, though formal variations are possible. Such formal variations do, however, imply either different epidemiological assumptions or different assumptions about error terms from observational errors or intrinsic noise. The prototype for this class is ordinary least squares for the response variable $y$ as a function of the geographical sub-entity $j$ and time index $t$,

$$y_{j,t} = \alpha_j + \sum_i \beta_i X_{j,t,i} + \epsilon_{j,t}, \tag{8}$$

where $\alpha_j$ are entity fixed effects, $\beta_i$ is the regression coefficient (effect size) for intervention $i$, $X_{j,t,i}$ is the matrix of explanatory variables (which may be lagged to account for a delay between the activation and the effect of interventions), and $\epsilon_{j,t}$ is a normally-distributed error term with variance $\sigma^2$, $\epsilon_{j,t} \sim \mathcal{N}(0, \sigma^2)$.

Variations included in this class cover, e.g., general additive models that allow for a non-linear dependence on the explanatory variables by promoting $\beta_i$ to a function,

$$y_{j,t} = \alpha_j + \sum_i \beta_i(X_{j,t,i}) + \epsilon_{j,t}, \tag{9}$$

generalised linear models that assume a non-normal distribution of the errors, as well as models for temporal autocorrelation and spillover between geographical entities, which will be discussed below.

About 120 studies in the reviewed corpus used multiple regression models or variations thereof.

*StopptCOVID* formulates its epidemiological assumptions as a (generalised) linear model, and as such multiple linear regression needs to be included in this study.

*a. Decision:* For the purpose of validation, it was decided that several linear regression-type models should be formulated to address the statistical issues in the baseline model. These models should explore appropriate models for the (autocorrelated) noise (Section S4) and techniques for dealing with the problem of multicollinearity (Section S5) in multiple regression.

*Dynamical Models*

This second category of models explicitly starts from the description of infectious disease spread as a dynamical system described by continuous or discrete time evolution equations. The prototype for such models is the classical SIR model [35, 36] with evolution equations for the time-dependent fractions $S$, $I$, and $R$ of susceptible, infected and recovered individuals and intervention effects in the growth rate,

$$\dot{S} = -\Big(\beta_0 + \sum \beta_i X_i\Big) S I, \quad \dot{I} = \Big(\beta_0 + \sum \beta_i X_i\Big) S I - \gamma I, \quad \dot{R} = \gamma I, \tag{10}$$

where $\beta_0$ is the growth rate in the absence of interventions, $\beta_i$ is again the effect size of intervention $i$, and $\gamma$ is the recovery rate; note that a possible dependence on geographical entities is not explicitly included in the notation to avoid clutter. Some metric for goodness of fit needs to be prescribed for estimating coefficients, e.g., a likelihood function for the difference $\mathcal{I} - \mathcal{I}_{\text{obs}}$ of the observed rate of infections $\mathcal{I}_{\text{obs}}$ from the predicted one.

Variations in this class include, e.g., a larger number of compartments to better model the time dependence of infectivity, asymptomatic transmission, or age dependence, or to account for vaccination status. Furthermore different choices for optimising the quality of fit (e.g., the choice of the likelihood function in maximum-likelihood or Bayesian approaches) are possible.

One should note that the distinction between linear regression models and dynamical models can become blurred on the technical level, especially when the depletion of susceptibles is ignored in dynamical models. For example, a regression model with autocorrelated error terms and certain dynamical models may in practice both be formulated as state space models [37, 38] for estimation. Sometimes the estimation problem can be explicitly transformed into a generalised linear model.

Dynamical models are used by about 30 models in the reviewed corpus. A potential advantage of dynamical models is that they can be fitted directly to case data, i.e., connect to the observational data more directly and consistently. It is particularly important to explore whether fitting the reconstructed $\mathcal{R}(t)$ instead of the case data introduces any biases or affects the confidence intervals for regression coefficients. Furthermore, a correct treatment of vaccination (Section S1 C) is readily possible with a dynamical model. On the downside, properly modelling the noise in dynamical models is not trivial due to non-stationarity, i.e., the noise amplitude depends on time as case numbers go up and down (see Section S6). Numerically, the estimation problem for dynamical models may be ill-conditioned, and convergence may be slow. For truly non-linear models, iterative solvers for fitting the model may not converge to the global optimum.

*b. Decision:* On balance, it was decided that a dynamical model should be included in the multi-model comparison, but that this model should stick as closely as possible to the epidemiological assumptions made by *StopptCOVID*. In addition, a dynamical model should be used to more accurately model the effect of vaccination.

#### *Machine Learning Methods*

This third class of models more radically relaxes assumptions about the functional dependence between the explanatory variables $X_i$ and the response variable $y$ to the general form

$$y = y(X_1, X_2, X_3 \ldots). \tag{11}$$

Similar to regression models, further embellishments can be included in this functional dependence, e.g., a time-dependent response to interventions (Section S3 B). The functional dependence may be constructed with various machine-learning techniques, e.g., in the form of averages over multiple decision trees (random forest regression), with support vector machines, or neural networks.

Less than a dozen studies in the corpus used such machine learning algorithms. These methods are attractive in that they provide for a more flexible functional dependence of the response variable on the explanatory variables, and can accommodate non-linear interactions between NPIs. Drawbacks may include the need for considerable amounts of training data for determining the large number of parameters "under the hood"[1] in these models, less straightforward estimation

[1] The term "non-parametric" can be somewhat misleading in the context of machine learning models. Any tunable algorithm for mapping input variables to output variables contains parameters, e.g., the decision boundaries in decision trees. What distinguishes such "non-parametric" models is rather that the parameters are not of interest *individually* and usually remain hidden from the user.

of confidence intervals, and the risk of overfitting. In addition, care is required to project back to linear effect estimates that can be compared with regression models, but this can be solved (Section S6).

*c. Decision:* Since machine learning methods are very distinct from linear regression methods and dynamical model, we determined that it is pertinent to compare the performance of these different classes of methods in the context of time series analyses for NPI evaluation. In order to truly cover complementary methods, an algorithm completely distinct from linear regression should be included, such as the decision-tree based methods in [39, 40].

*Trend- and Level-Change Methods*

A fourth class of models (which in practice shares some features with the regression models described earlier) focuses on trend or level changes in the response variable before and after the introduction of specific interventions. A prototype for this class is segmented regression for interrupted time series with different intercepts $\alpha_{\text{pre}}$ and $\alpha_{\text{post}}$ and different slopes $\beta_{\text{pre}}$ and $\beta_{\text{post}}$ before and after some intervention is switched on at time $t_{\text{start}}$,

$$y_t = \begin{cases} \alpha_{\text{pre}} + \beta_{\text{pre}}(t - t_{\text{start}}) + \epsilon_t, & t \leq t_{\text{start}} \\ \alpha_{\text{post}} + \beta_{\text{post}}(t - t_{\text{start}}) + \epsilon_t, & t > t_{\text{start}} \end{cases} \tag{12}$$

The changes in intercept and slope then serve to quantify the effect of the intervention. The key difference to the aforementioned linear regression methods is that different regression coefficients for the intervention and non-intervention periods are used instead of simply lumping the effect into the regression coefficient for the corresponding explanatory variable. Generalisations in this broad class of segmented regression models include, e.g., the use of higher-order polynomials to capture the trend before and after intervention and the detection of break points or jumps in the time series by statistical methods instead of specifying the intervention dates manually [see, e.g., 41–43, for a discussion of these methods]. The difference-in-differences (DiD) method [44, 45] straddles the gap between trend-change methods and the regression methods from Section S3 A. In this approach, the post-intervention level[2] is not simply compared to the pre-intervention phase of the intervention group, but the level change is compared to that in a control group. Adding a control group has the potential benefit of reducing the risk of misattributing accidental trend changes as

[2] In principle, difference-in-differences can be generalised to consider changes in slopes or higher-order derivatives as well, but these ramifications are immaterial here.

intervention effects. The DiD approach, like other approaches for introducing control groups – e.g., synthetic controls; [46, 47] – also entails implicit assumptions to justify the suitability of the control group as a basis of reference.

Thirty-five of the reviewed studies were classified as belonging to this class of methods [e.g., 48–50], and it has also been used in others [e.g., 51]. Compared to most[3] of the methods in the previous sections, a potential advantage of methods based on trend changes is that appropriate trend subtraction can partly remove errors due to unmodelled processes for more accurate effect estimates.

However, trend- and level-change methods are difficult to implement and automatise when multiple interventions are switched on and off in irregular patterns, as was the case with NPIs in Germany. In this case, it is difficult or impossible to define appropriate intervals for piecewise fits before and after the switch-on points of a given NPI, unless one only considers step changes and controls for the other NPI variables, which then just amounts to multiple regression. Similarly, it is not feasible to associate breaks in the time series unambiguously with individual NPI.

Multiple regression could still be extended and possibly improved by including DiD terms, but this would require a control group for the NPIs under consideration. Such a non-intervention group is not available for all of the NPIs investigated by *StopptCOVID*, however.

*d. Decision:* Because of these obstacles, trend- and level-change methods were deemed unsuitable for reevaluating NPI effects based on the *StopptCOVID* data, although they would be useful methods for analysing effects of NPI with one or a few well-defined intervention points. An earlier plan for the *StopptCOVID* project apparently envisaged an analysis of trend changes[4], but this plan understandably seems to have been abandoned.

*Other approaches*

More than 100 studies used methods that are not applicable to the problem of estimating the effect of several dozen interventions with general activation patterns. This includes methods that may be appropriate for other purposes, but can, e.g., only be applied to a single intervention or consider global outcomes, such as total infections over an extended period, rather than time

[3] Some methods with appropriate (e.g., autoregressive) noise models effectively also accomplish (some) trend subtraction.

[4] See `https://www.uni-bielefeld.de/fakultaeten/gesundheitswissenschaften/ag/ag2/forschung/stopptcovid.xml`.

series data [52–54]. Some studies were purely simulation-based and did not attempt to statistically estimate effect sizes. Some studies, on the other hand, offer little more than a description of the epidemic trajectory, visual comparison to simple exponential extrapolation, or are extremely deficient in the description of their methodology. A very small number of papers was not accessible to the project team.

#### B. Time Dependence of the Response Function

Models may make different assumption about the time-dependent influence of an intervention at a given time on the subsequent evolution of the growth rate. In the simplest case, the growth rate at any given instant is determined by the interventions *only* at one instant in time, e.g., at the exact time $t$ under consideration, or perhaps at some past date with a specific lag $\Delta t$. In a simple time-dependent regression model, this translates to a relationship between the explanatory variables (for interventions) and the response variable (growth rate) of the form,

$$y_t = \alpha + \sum_i \beta_i X_{t-\Delta t,i} \tag{13}$$

Instead, the growth rate at time $t$ can be taken to depend on the past history of intervention $i$ via a time-dependent response function $G_{i,\tau}$,

$$y_t = \alpha + \sum_i \sum_{\tau=1} G_{i,\tau} X_{t-\tau,i}. \tag{14}$$

Some readers may find this distinction more intuitive if expressed in terms of continuous functions, where the nature of $G$ as a Green's function becomes manifest

$$y(t) = \alpha + \sum_i \int_0^\infty G_i(\tau) X_i(t-\tau)\,\mathrm{d}\tau. \tag{15}$$

A dependence of $y(t)$ just on a single instant in time can formally be expressed by a $\delta$-function as response function,

$$y(t) = \alpha + \sum_i \beta_i X_i(t-\Delta t) = \alpha + \sum_i \int_0^\infty \beta_i \delta(\tau-\Delta t) X_i(t-\tau)\,\mathrm{d}\tau. \tag{16}$$

Only about two dozen of the reviewed studies consider a non-trivial time-dependent response function (i.e., different from a $\delta$-function). Different methods are used for estimating time-dependent response functions. Both regression techniques and machine-learning techniques can in principle estimate $G_i(\tau)$ by explicitly including the time since the activation of a specific NPI as an explanatory variable. This comes at the expense of a significant proliferation of parameters and consequently dilutes statistical power or invites overfitting. Another approach explicitly builds in a time dependence of the response variable on its own past history (autocorrelation) up to a specific time lag $q$ into regression models. This leads from Equation (9) to the class of autoregressive AR($q$) models of order $p$ with exogenous regressors,

$$y_{j,t} = \sum_{\tau=1}^{p} \phi_\tau y_{j,t-\tau} + \alpha_j + \sum_i \beta_i X_{j,t,i} + \epsilon_{j,t}, \tag{17}$$

with autoregression coefficients $\phi_\tau$. Note that the relation between the response function $G_{i,\tau}$ and the coefficients $\phi_\tau$ is generally non-trivial and non-zero for *any* $\tau > 0$ for AR($q$) processes; for AR(1), one has $G_{i,\tau} = \beta_i \phi_1^\tau$. The response function $G_{i,\tau}$ must also be distinguished from the impulse response of the underlying AR($q$) process, which does not include the regression coefficient for the exogenous variable.

These models can be further generalised to allow for a dependence of response variables in different entities $j$ on each other by promoting the autoregression coefficients to matrices (VAR and VARMAX models[5] [55, 56]). This approach is not limited to a single response variable, but can model the interaction of several endogenous variables, e.g., NPIs, human behaviour and disease spread in the context of COVID-19 [57].

It must be emphasised that the inclusion of autoregressive terms for the response variable amount to a change of epidemiological model assumptions. This can be illustrated for the simplest case of an AR(1) model,

$$y_{j,t} = \phi_1 y_{j,t-1} + \alpha_j + \sum_i \beta_i X_{j,t,i} + \epsilon_{j,t}, \tag{18}$$

by turning it into a modified differential equation using Taylor series expansion,

$$(1 - \phi_1) y + \phi_1 \dot{y} = \alpha + \sum_i \beta_i X_i(t). \tag{19}$$

Using $y = \ln \mathcal{R}$ as the response variable, this results in the model,

$$\frac{\mathrm{d} \ln \mathcal{R}}{\mathrm{d}t} = -\frac{(1 - \phi_1)}{\phi_1} \ln \mathcal{R} + \frac{1}{\phi_1} \left( \alpha + \sum_i \beta_i X_i(t) \right), \tag{20}$$

where the autoregressive term effectively drives $\ln \mathcal{R}$ to zero ($\mathcal{R}$ to 1) for $\phi_1 < 1$.

[5] Spatial coupling (Section S3 C) may also be included in this approach.

Furthermore, the interpretation of the regression coefficients changes when AR($p$) terms are present. The proper, asymptotic effect of the intervention settings $X_i$ must be estimated from the limit where they are switched on continuously; in an $AR(1)$ model, this leads to

$$\ln \mathcal{R} \to \frac{1}{1-\phi_1}\left(\alpha + \sum_i \beta_i X_i(t)\right). \tag{21}$$

Hence the asymptotic effect of an intervention with regression coefficient $\beta_i$ in an AR(1) model is the same as for a regression coefficient $\beta_j/(1-\phi_1)$ in the basic linear regression model (9).

Furthermore, autoregressive terms in the response variable need to be carefully distinguished from autoregressive *error* terms, whose inclusion leads to ARMAX-type models of order $(p, q)$,

$$y_{j,t} = \sum_{\tau=1}^{p} \phi_\tau y_{j,t-\tau} + \alpha_j + \sum_i \beta_i X_{j,t,i} + \epsilon_{j,t} + \sum_{\tau=1}^{q} \theta_\tau \epsilon_{j,t-\tau}, \tag{22}$$

with autoregression coefficients $\theta_\tau$ for for the moving-average (MA) error terms up to order $q$. Such terms represent autocorrelation in intrinsic noise responsible for deviations of the model from the observed ground truth and are further discussed in Section S4 B. Different from the AR terms, they do not affect the interpretation of the regression coefficients.

It is worth mentioning that dynamical models for case numbers $\mathcal{I}$ are also formally autoregressive, but this implies autocorrelation only for case numbers (and model residuals), but not for $\ln \mathcal{R}$. Hence such models do *not* automatically account for autocorrelated noise in $\ln \mathcal{R}$.

With regard to the determination of NPI effects, time-dependent response function represent a well-motivated epidemiological extension of the baseline model. However, their inclusion faces several obstacles. If the response functions for individual NPIs are modelled with an explicit time dependence rather than lumping that time dependence into (V)ARMAX coefficients, the number of model parameters grows considerably, and sufficient amounts of data to constrain them may no longer be available. Second, if smoothed data for $\mathcal{R}(t)$ are used, the smoothing will contaminate the true time dependence of the response function.

*a. Decision:* Because of these obstacles, and since a time-dependent response function would be an extension of the epidemiological assumptions, it was decided not to include this feature in Work Package 1. The time dependence of the response function may be revisited in the later work packages if the available data permit.

### C. Spatial Coupling

*StopptCOVID* considers disease spread in geographical sub-units to be independent of each other. In reality, infections can be transmitted[6] to other sub-units, e.g., due to human travel. Such spatial coupling is included only in eight studies considered in the literature review.

Formally, spatial coupling between response variables in different geographical entities can be implemented in regression models, dynamical models, and machine learning models relatively easily, e.g., by generalising the basic linear regression model (9) to

$$y_{j,t} = \alpha_j + \sum_i \beta_i X_{j,t,i} + \sum_k K_{jk} y_{k,t-1} + \epsilon_{j,t}, \tag{23}$$

where the matrix $K_{jk}$ describes the coupling of the response variables to its previous values *anywhere* on the previous day.[7]

While the inclusion of such spatial coupling terms is justified epidemiologically, some caveats about their practical use in NPI effect estimation must be made. First, spatial coupling along the lines of Equation (23) is sensible for incident cases $\mathcal{I}$ as response variable, but not for the growth rates or growth factors (like $\mathcal{R}$); what spreads in space are infections and not growth rates of infections. The effect of interventions on the growth rate can still be inferred from spatial models that fit $\mathcal{I}$, but considerable care is required.

Furthermore, estimation of $K_{jk}$ will usually not be feasible without specifying the form of the coupling matrix due to the large number of free parameters. If $K_{jk}$ is specified as $K_{jk} \propto K(d_{jk})$ in terms of a transmission kernel $K$ that depends on some specified distance metric $d_{jk}$ between geographical entities, the choice of the kernel function and distance metric is not trivial, e.g., because the amount of travel rather than geographical distance between sub-units may be the deciding factor, and ought to be based on solid epidemiological data [22, 58, 59]. Even when the form of the kernel function is known, similar infection dynamics across sub-units may make the estimation of non-local coupling from case number alone a highly ill-conditioned problem. Ideally, the transmission kernel would have to be reconstructed from detailed epidemiological data on infection chains, which may not be available (and is certainly not available for the evaluation of NPIs in Germany).

Furthermore a meaningful analysis of spatial disease spread will likely need to consistently work with more fine-grained (e.g., county-level) data than with aggregated state-level data. While

[6] We avoid the term common term "spillover" from econometrics because "spillover" tends to have a more restricted meaning in infectious disease epidemiology.

[7] Note that this also introduces temporal autoregression.

such data are in principle available, including it would still imply a significant redesign of data preparation for the baseline model. There are also concerns that low case numbers at the county level may limit the power of a fine-grained spatio-temporal analysis. On the other hand, case imports between states as larger entities are likely less critical than between counties, so there is justification for neglecting spatial dynamics at the state level.

*a. Decision:* Because of the above considerations, it was decided not to consider spatial dynamics in Work Package 1, but to reconsider this issue for Work Package 2 if deemed feasible.

#### D. Bayesian vs. Non-Bayesian Methods

In Bayesian models, the (posterior) probability density[8] $p(\boldsymbol{\theta}|\mathbf{y})$ for a vector $\theta$ of parameter values (which will, e.g., contain the intercepts and regression coefficient in a linear regression model) given observed data $\mathbf{y}$, is expressed in terms of the likelihood $p(\mathbf{y}|\boldsymbol{\theta})$, the probability of $p(\mathbf{y})$ of the data, and the assumed prior probability $p(\boldsymbol{\theta})$ of the parameter values,

$$p(\boldsymbol{\theta}|\mathbf{y}) = \frac{p(\mathbf{y}|\boldsymbol{\theta})}{p(\mathbf{y})} p(\boldsymbol{\theta}), \tag{24}$$

which is just Bayes' theorem for probability distributions. Here, $p(\mathbf{y})$ is an unknown a-priori-probability, and needs to be obtained from a weighted integral over the likelihood,

$$p(\boldsymbol{\theta}|\mathbf{y}) = \frac{p(\mathbf{y}|\boldsymbol{\theta})}{\int p(\mathbf{y}|\boldsymbol{\theta}) p(\boldsymbol{\theta}) \, \mathrm{d}\boldsymbol{\theta}} p(\boldsymbol{\theta}). \tag{25}$$

The Bayesian approach can be generalised to include, e.g., hyperparameters for the likelihood and multiple levels of models in a hierarchical approach.

About two dozen of the studies surveyed in the literature review use a Bayesian approach for determining NPI effects from time series data [e.g., 49, 60–63]; another more recent example is [64]. In general, Bayesian approaches have several attractive features. They can incorporate prior knowledge on (some) parameters as informative priors, which can be particular useful for breaking parameter degeneracies due to multicollinearity. Weak, non-informative priors may still be useful for addressing multicollinearity by regularisation (cp. Section S5). Bayesian methods, if used properly, are advantageous for dealing with complex likelihood functions in non-linear models that exhibit multiple local maxima that result in complex-shaped and even disjoint credible

[8] Note that we will use the term "probability" for "probability density" for the sake of brevity when there is little danger of confusion.

regions. As a downside, sampling a high-dimensional parameter pace in a Bayesian approach may be computationally expensive when combined with complex ODEs or renewal models. Furthermore, properly and efficiently accounting for autocorrelation in time series in a Bayesian approach is not trivial. Autocorrelation is not addressed rigorously in any of the reviewed studies; the incorporation of a weekly random walk component in $\mathcal{R}(t)$ in [63] is a notable attempt to incorporate it, but remains a quick fix. More rigorous methods exist, e.g., Bayesian versions of (V)ARMA models, but again these come at added computational cost.

*a. Decision:* In the context of the present study with its focus on statistical errors due to autocorrelation and multicollinearity, it was determined that a Bayesian approach would not offer substantial advantages. Given substantial uncertainties and often methodological shortcomings in NPI research, incorporation of assumed prior knowledge is best avoided. Linear regression problems, including more general forms such as autoregressive linear models, involve concave likelihood functions and will not produce complex-shaped confidence regions. After also considering the technical obstacles and computational costs, we therefore determined not to include a Bayesian approach. However, these considerations are highly specific to the design of this study. The strengths of Bayesian approaches in a broader context were acknowledged, and the use of Bayesian methods will be revisited in the other work packages, where their advantages will become relevant.

### S4. ERROR ESTIMATES FOR EFFECT SIZES

The literature review revealed that the description of the error (noise) model and statistical uncertainty quantification was often more cursory than the description of the technique for obtaining point estimates, and sometimes lacking altogether. Similarly, only a fraction of studies explicitly considered the issue of multicollinearity, and those that did often relied on *ad hoc* procedures for variable selection. For these reasons, methods for error analysis and addressing multicollinearity were not coded into explicit categories during the literature search.

In light of the sparse and scattered documentation of error analysis in the literature on NPIs, this section seeks to provide a short pedagogical review of standard methods for computing error bars for regression models based on time series data.

In discussing error bars for the estimated NPI effects, it is important to bear in mind that there are several distinct notions of error bars, reflecting different approaches to statistics (frequentist,

Bayesian, likelihoodist). Much of the discussion in this section will focus on frequentist confidence intervals, i.e., intervals constructed thus that they will contain the true values of parameters with a given coverage probability (e.g., 95%) for repeated random realisations of a "true" model; for the distinction from Bayesian credible intervals see, e.g., [65, 66]. The actual calculation of error intervals again entails statistical estimation and, very often, assumptions that only hold approximately in practice, making the *actual* coverage probability different from the desired one. Especially for more complex statistical analyses, it is therefore important to review conceptually *how* confidence intervals are constructed. This section seeks to elucidate this process both to starting practitioners and researchers from others fields who may rely on results from time series studies, e.g., for guideline development.

#### A. Linear Regression Model: Sandwich Estimators

Let us first consider error estimates for the regression coefficients $\beta_j$ in ordinary least squares (OLS) regression with a matrix of regressors $X_{ti}$ and error terms $\epsilon_t$,

$$y_t = \sum_i \beta_i X_{ti} + \epsilon_t, \tag{26}$$

or in index-free vector notation,

$$\mathbf{y} = \mathbf{X} \cdot \boldsymbol{\beta} + \boldsymbol{\epsilon}. \tag{27}$$

The solution is given by

$$\boldsymbol{\beta} = (\mathbf{X}^\top \cdot \mathbf{X})^{-1} \cdot \mathbf{X}^\top \cdot \mathbf{y}, \tag{28}$$

where $\mathbf{X}^\top$ denotes the transpose of $\mathbf{X}$.

Assuming that $\boldsymbol{\beta}$ is the true solution, one can construct confidence intervals by considering how much the estimate for $\boldsymbol{\beta}$ is perturbed by noise $\boldsymbol{\delta y}$ in the response variable. Introducing auxiliary matrices $\mathbf{D} = (\mathbf{X}^\top \cdot \mathbf{X})^{-1}$ and $\mathbf{M} = (\mathbf{X}^\top \cdot \mathbf{X})^{-1} \cdot \mathbf{X}^\top = \mathbf{D} \cdot \mathbf{X}^\top$ to simplify notation, the perturbation of the regression coefficient becomes $\boldsymbol{\beta}$

$$\boldsymbol{\beta} + \boldsymbol{\delta\beta} = \mathbf{M} \cdot (\boldsymbol{y} + \boldsymbol{\delta y}), \tag{29}$$

or, reverting to index notation,

$$\delta\beta_i = \sum_l M_{il} \delta y_l. \tag{30}$$

Thus, the covariance between the regression coefficients $\beta_i$ and $\beta_k$ is

$$\mathrm{cov}\,(\beta_i, \beta_k) = \langle \delta\beta_i\, \delta\beta_k \rangle = \sum_{l,m} \langle M_{il} M_{km} \delta y_l \delta y_m \rangle = \sum_{l,m,s,r} \langle D_{ir} X_{lr} \delta y_l D_{ks} X_{ms} \delta y_m \rangle\,, \tag{31}$$

where angled brackets denote expectation values for different error realisations. This can be written in index-free notation as

$$\langle \delta\boldsymbol{\beta}\, \delta\boldsymbol{\beta} \rangle = \mathbf{D} \cdot \langle \mathbf{X}^\top \cdot (\delta\boldsymbol{y}\, \delta\boldsymbol{y}) \cdot \mathbf{X} \rangle \cdot \mathbf{D}^\top. \tag{32}$$

where the (transformed) matrix of error covariance appears in the middle; hence this type of estimator for the (co)variances of the regression parameters is commonly know as “sandwich estimator”. The covariance matrix of the errors $\delta\boldsymbol{y}$ needs to be approximated based on the statistical properties of the residuals $\boldsymbol{\Delta y}$ for the fitted model. If the errors are normally distributed, the vector of regression parameters will also follow a (multivariate) normal distribution, and confidence regions can be constructed from the covariances from Equation (32).

Under the assumption that the errors are independent of each other and that their distribution is independent of entity and time, one can approximate the covariance matrix as a multiple of the identity matrix,

$$\langle \delta y_l\, \delta y_m \rangle = \frac{\delta_{lm}}{n-p} \sum_t \delta y_t^2 =: \delta_{lm} \sigma^2, \tag{33}$$

where $p$ is the number of fitted parameters, and $\sigma^2$ is the variance of the error distribution. This implies that the variance of individual regression coefficients is

$$\mathrm{var}\,\beta_i = \sigma^2 (\mathbf{M} \cdot \mathbf{M}^\top)_{ii} = \sigma^2 [(\mathbf{X}^\top \cdot \mathbf{X})^{-1}]_{ii}. \tag{34}$$

Equations (31,32) show, however, that correlations in the errors (e.g., autocorrelation in time) immediately affect the error bars of the regression parameters. They form the starting point for estimating errors of regression coefficients with correlated and heteroskedastic errors (i.e., errors whose distribution parameter vary across entities and time) based on certain assumption for the covariance matrix of the errors. This is achieved by using a more general form for the error covariance matrix, whose elements again need to be estimated from the regression residuals. For example, the White estimator [67] retains a diagonal error covariance matrix, but allows the diagonal elements to be non-equal to account for non-constant error variance across time (temporal heteroskedasticity). The Newey-West estimator [68] further estimates non-zero covariance between errors at different times up to a specified lag $L$ to account for autocorrelation. Driscoll &

Kraay [69] further generalised the Newey-West estimator to include both temporal autocorrelation as well as correlation across entities. The Driscoll-Kraay estimator represents one of the most general "robust" estimators for correlated errors across time and entities that is readily available in common statistical software [e.g., 70].

We note in passing that the case of weighted least squares (WLS) regression can be treated in a very much analogous manner. Whereas OLS minimises the squared sum of residuals, $|\boldsymbol{y} - \mathbf{X} \cdot \boldsymbol{\beta}|^2$, WLS minimizes $(\boldsymbol{y} - \mathbf{X} \cdot \boldsymbol{\beta})^\top \cdot \mathbf{W} \cdot (\boldsymbol{y} - \mathbf{X} \cdot \beta)]$ with some weight matrix[9] $\mathbf{W}$. A WLS regression problem can be transformed into an OLS problem for $(\hat{\mathbf{y}} - \hat{\mathbf{X}} \cdot \boldsymbol{\beta})^2$, with a transformed design matrix $\hat{\mathbf{X}} = \sqrt{\mathbf{W}} \cdot \mathbf{X}$ and regressand $\hat{\mathbf{y}} = \sqrt{\mathbf{W}} \cdot \mathbf{y}$. The computation of variances of the regression parameters proceeds analogously to the OLS case.

For models that explicitly include autocorrelated error terms, and for connecting to the Bayesian and likelihoodist viewpoint, it is useful consider the calculation regression coefficients and their errors in the framework of maximum likelihood estimation (MLE). If we assume independent, normally distributed errors with variance $\sigma^2$ for each measured data point, $\epsilon_i \sim \mathcal{N}(0, \sigma^2)$, then the likelihood $\mathcal{L}$ becomes,

$$\mathcal{L} = \prod_t \frac{1}{\sqrt{2\pi}\sigma} e^{-\frac{\epsilon_t^2}{2\sigma^2}} = \prod_t \frac{1}{\sqrt{2\pi}\sigma} e^{-\frac{(y_t - \sum_i \beta_i X_{ti})^2}{2\sigma^2}}, \tag{35}$$

i.e., it is a product of the Gaussian probability densities for the residual $y_t - \sum_i \beta_i X_{ti}$ over all data points. For practical calculations, the log-likelihood $\ln \mathcal{L}$ is more convenient,

$$\ln \mathcal{L} = -\frac{N}{2} \ln 2\pi - N \ln \sigma - \frac{1}{2\sigma^2} \sum_t \left( y_t - \sum_i \beta_i X_{ti} \right)^2. \tag{36}$$

Maximum likelihood estimates can be found by requiring that all the partial derivative of $\ln \mathcal{L}$ vanish, in particular,

$$\frac{\partial \ln \mathcal{L}}{\partial \beta_k} = \frac{1}{\sigma^2} \sum_t X_{tk} \left( y_t - \sum_i \beta_i X_{ti} \right) = 0. \tag{37}$$

This is tantamount to the normal equation $(\mathbf{X}^\top \cdot \mathbf{X}) \cdot \boldsymbol{\beta} = \mathbf{X}^\top \cdot \mathbf{y}$ for linear regression, and immediately leads to the solution from Equation (28). Furthermore, since the likelihood is quadratic in the regression coefficients $\beta_i$, the likelihood for the vector $\boldsymbol{\beta}$ (with $\sigma$ held fixed) will be that of a multivariate Gaussian centred around its MLE value $\bar{\boldsymbol{\beta}}$ with some covariance matrix $\boldsymbol{\Sigma}$,

$$\mathcal{L}(\boldsymbol{\beta}) = \mathcal{L}(\bar{\boldsymbol{\beta}}) - \frac{1}{2} (\boldsymbol{\beta} - \bar{\boldsymbol{\beta}})^\top \cdot \boldsymbol{\Sigma}^{-1} \cdot (\boldsymbol{\beta} - \bar{\boldsymbol{\beta}}). \tag{38}$$

[9] The weight matrix is usually diagonal, but from a technical point of view, it merely needs to be positive definite. WLS with a non-diagonal weight matrix effectively incorporates correlated errors with a known form of the error covariance matrix up to a scalar factor.

The coefficients of the quadratic terms in $\boldsymbol{\beta}$ can be obtained from the second-order derivative of $\ln \mathcal{L}$, i.e., from its Hessian matrix. The covariances of the regression coefficients (components of $\boldsymbol{\Sigma}$) are given by negative inverse of the Hessian,

$$\mathrm{cov}\,(\beta_i, \beta_k) = -\left(\frac{\partial^2 \ln \mathcal{L}}{\partial\beta_i \partial\beta_k}\right)^{-1}. \tag{39}$$

This result is identical to Equation (34). Equation (39) implies that for standard OLS with uncorrelated Gaussian errors, the covariances of the regression parameters can also be used to define likelihood contours. In this case (but not in general), the confidence intervals will also be identical to Bayesian credible intervals for the marginalised one-parameter posterior distributions in the case of uniform priors.

### B. ARMA Errors

Instead of estimating the temporal autocorrelation structure of unmodelled processes or observational errors based on the residuals of an OLS/WLS model, one can explicitly include autocorrelation in the noise by including lagged terms up to $q$ time steps akin to the those for the response variable in AR($p$) models; these are known as MA($q$) ("moving average") errors,

$$y_{j,t} = \alpha_j + \sum_i \beta_i X_{j,t,i} + \epsilon_{j,t} + \sum_{\tau=1}^{q} \theta_\tau \epsilon_{j,t-\tau}, \tag{40}$$

which is explicitly written for panel data with a dependence on entity $j$. Ideally, the newly added errors $\epsilon_{j,t}$ (innovations) will form a time series that is no longer autocorrelated; this generally requires selecting the appropriate number of lags and an autocorrelation structure that is well captured by MA($q$) models. In this case, error bars for the regression coefficient can then be computed by treating the innovations as independent random variables.

Note that there is a subtle distinction in regression models with MA($q$) errors between the (total) fit error or residual for the linear effect model $y_{j,t} = \alpha_j + \sum_i \beta_i X_{j,t,i}$ and the innovations. The total residual is given by a weighted sum of innovations, $\epsilon_{j,t} + \sum_{\tau=1}^{q} \theta_\tau \epsilon_{j,t-\tau}$.

Moving-average MA($q$) errors do not, however, constitute the most general form of autocorrelated regression errors within the framework of ARMA models. One can instead model the total regression residual $n_{j,t}$ as arising from an ARMA process (regression with ARMA errors) with *both* AR($p$) and MA($q$) terms,

$$y_{j,t} = \alpha_j + \sum_i \beta_i X_{j,t,i} + n_{j,t} \tag{41}$$

$$n_{j,t} - \sum_{\tau=1}^{p} \phi_\tau n_{j,t-\tau} = \epsilon_{j,t} + \sum_{\tau=1}^{q} \theta_\tau \epsilon_{j,t-\tau}. \tag{42}$$

AR($p$) error terms lead to a different autocorrelation structure of the noise; whereas MA($q$) errors are uncorrelated for lags bigger than $q$, AR($p$) are generally correlated for arbitrary lags.[10]

Conceptually the calculation of confidence intervals from the residuals or from the likelihood carries over almost directly from the case of OLS and WLS discussed in Section S4 A to the case of MA($q$) errors, or more general regression models with ARMA errors. There is, however, an important technical difference, regardless of whether estimation follows a least-square or maximum-likelihood approach.[11] This is best illustrated for the simple case of an MA(1) model of a single time series,

$$y_t = \alpha + \sum_i \beta_i X_{t,i} + \epsilon_t + \theta_1 \epsilon_{t-1}. \tag{43}$$

The innovations $\epsilon_t$ are required to construct the likelihood for the observed data given specific model parameters,

$$\ln \mathcal{L} = -\frac{N}{2} \ln 2\pi - \frac{N}{2} \ln \sigma^2 - \sum_t \frac{\epsilon_t^2}{2\sigma^2} - \ln \det \left( \frac{\partial \epsilon_{t'}}{\partial y_t} \right). \tag{44}$$

Here, the determinant term accounts for the coordinate transformation that is required to express $\mathcal{L}$ as a likelihood density on the observed data instead of the innovations.

However, different from the likelihood for OLS regression in (38), the innovations can no longer be directly replaced with the residuals, rather one needs to solve the linear system of equations (43) for the innovations $\epsilon_t$. The conditions for maximising the likelihood,

$$\frac{\partial \ln \mathcal{L}}{\partial \sigma^2} = 0, \ \frac{\partial \ln \mathcal{L}}{\partial \theta_1} = 0, \ \frac{\partial \ln \mathcal{L}}{\partial \alpha} = 0, \ \frac{\partial \ln \mathcal{L}}{\partial \beta_k} = 0, \tag{45}$$

then become a system of coupled *non-linear* equations, and need to be solved iteratively.

The covariance matrix of regression parameters can again be obtained from the inverse of the Hessian. Note that for linear models, the Hessian $\partial^2 \ln \mathcal{L} / \partial \beta_i \partial \beta_k$ does not depend on the regression

[10] For a given set of autorgression coefficients $\phi_\tau$ and $\theta_\tau$, models with ARMA errors can be transformed into equivalent ARMAX models (Equation 22) with lag operators acting on both the response variable *and* the explanatory variables, but the equivalent formulation as ARMAX model is not particularly useful for the purpose of this paper.

[11] For certain orders $(p, q)$ the ARMA coefficients can be obtained by simpler means, and there are also approximate methods as alternatives to the full MLE approximation.

coefficients $\beta_i$ because the log-likelihood is a quadratic function. In practice, outer product of gradient estimation [71] is often employed; we use it as well in `statsmodels`. By means of the information matrix equality,

$$-\left\langle \frac{\partial^2 \ln \mathcal{L}}{\partial\beta_i \partial\beta_k} \right\rangle = \left\langle \frac{\partial \ln \mathcal{L}}{\partial\beta_i} \frac{\partial \ln \mathcal{L}}{\partial\beta_k} \right\rangle, \tag{46}$$

one can obtain the expectation value of the Hessian from the expectation value[12] of the outer product of the gradient (score vector) $\partial \ln \mathcal{L}/\partial\beta_i$ of the log-likelihood with itself. This obviates the need for constructing the Hessian and ensures that the estimated inverse of the covariance matrix is symmetric and positive-definite.

#### C. Bootstrap

Another possibility to estimate the uncertainty of parameter estimates is to create different simulated data samples by appropriate random resampling either of the data themselves (case resampling) or of the residuals to create different simulated noise realisations. This *bootstrap* approach, pioneered by [72], is conceptually simple and can be applied quite generally, e.g., it provides a convenient way to determine confidence intervals for machine learning methods, where no closed-form solution for confidence intervals is available. Though bootstrapping can fail in pathological cases [e.g., 73] or sometimes in the context of shrinkage methods [74, 75], and though the optimal procedure for estimators bear some consideration [e.g., 76], non-parametric bootstrap methods generally tend to be quite versatile and robust.

Bootstrap techniques are employed in a number of studies of NPIs [e.g., 40, 77–80], in particular in conjunction with machine learning methods. The use of bootstrap methods on autocorrelated time series faces a complication, however. The resampling of the data or the residuals must not carried out completely randomly if different measurements or their errors are correlated. Thus, for time series data with autocorrelation, special variations of the bootstrap are required so that the simulated time series adequately retain the autocorrelation structure of the original data. This requirements leads to techniques that resample blocks of data (generally of varying length), such as the block bootstrap and stationary bootstrap [81, 82]; other methods exist as well [83] and may have advantages in specific context, such as the dependent wild bootstrap with its ability to handle

[12] The expectation values in Equation (46) are to be understood as weighted sums of the contribution of all data points to the Hessian and score vector.

irregularly spaced data [84]. Merely resampling geographical entities [40] will, in general, not capture the effects of autocorrelation in time. Admittedly, it is non-trivial to decide how to properly resample panel data [see, e.g., 85–87, for some discussion on the use of bootstrapping in this context]. For a bootstrap on the residuals, some possible choice include:

1. Resampling in time only with synchronisation across sub-units (i.e., the same reshuffling is applied for all sub-units). This implicitly assumes the noise to be correlated across sub-units.

2. Resampling in time only, but with independent resampling for each sub-unit.

3. Resampling both in time and across sub-units. For case resampling, this would break with the structure of the baseline *StopptCOVID* model, which explicitly includes fixed effects for *each* sub-unit, but for a bootstrap on residuals, this is a viable method.

The residuals of the baseline model show a number of temporally correlated features across several states. For this reason, we use synchronous resampling by default in this study, although independent resampling could also be justified. This choice for the bootstrap complements our use of frequency-domain methods (Section S4 D), for which we shall assume non-correlated residuals in different states. Results for the other two methods are, however, documented in Supplementary Methods S8).

The choice between resampling cases and residuals also bear some consideration. Case resampling will generally provide safer (but not necessarily better) estimates of error bars, and is generally the preferred method when there is serious danger of model misspecification or in conjunction with a method that is prone to overfitting (so that the residuals will underestimate the true noise). Thus, if the epidemiological assumptions of the baseline models are taken for granted, resampling of residuals is justified for a regression model, but in conjunction with a machine-learning approach with a flexible functional dependence of the response variable, case resampling is called for.

#### D. Frequency Domain Methods

A different approach to determine the statistical significance for serially correlated data and quantify uncertainties of derived parameter estimates is frequently used in the atmospheric sciences. To generate synthetic data (or noise) with the same autocorrelation properties as a given time series, Ebisuzaki [88] introduced a method that performs a Fourier decomposition of the time series and then generates synthetic time series for data or noise by randomising the phases of the Fourier coefficients. Ebisuzaki's method can be used to generate synthetic noise from residuals in general regression problems similar to a bootstrap on residuals, and is therefore applicable quite generically. For linear regression, however, the method can also be implemented analogously to a sandwich estimator, which obviates the need for sampling the noise distribution by expensive Monte Carlo simulations. As this may not be commonly known, we briefly explain this approach here. We directly consider the case of multiple time series in different (geographical) sub-units rather than a single time series.

The starting point is the discrete Fourier transform of the residuals $\delta y_{pl}$ (for time index $l$ and sub-unit $p$), which yields the Fourier coefficients $\delta\tilde{y}_{pl'}$ (with frequency index $l'$),

$$\delta\tilde{y}_{pl'} = \frac{1}{\sqrt{N}} \sum_{l} e^{-2\pi i l' l/N} \delta y_{pl}, \tag{47}$$

were $N$ is the length of the discrete time series. After expressing the time-domain residuals $\delta\tilde{y}_{lp}$ by the inverse transform, Equation (31) becomes,

$$\begin{aligned}\langle \delta\beta_j\, \delta\beta_k \rangle &= \sum_{p,q} \sum_{l,m} \langle M_{jpl} M_{kqm} \delta y^*_{pl} \delta y_{qm} \rangle = \sum_{p,q} \sum_{l,m} \left\langle \frac{M^*_{jpl} M_{kqm}}{N} \sum_{l'} e^{-2\pi i l' l/N} \delta\tilde{y}^*_{pl'} \sum_{m'} e^{2\pi i m' m/N} \delta\tilde{y}_{qm'} \right\rangle \\ &= \sum_{p,q} \sum_{l,m} \sum_{l',m'} \left\langle \frac{M^*_{jpl} M_{kqm}}{N} e^{-2\pi i l' l/N} \delta\tilde{y}^*_{pl'} e^{2\pi i m' m/N} \delta\tilde{y}_{qm'} \right\rangle = \sum_{p,q} \sum_{l',m'} \left\langle \tilde{M}^*_{jpl'} \tilde{M}_{kqm'} \delta\tilde{y}^*_{pl'} \delta\tilde{y}_{qm'} \right\rangle .\end{aligned} \tag{48}$$

Here the indices $l$ and $m$ denote time, $j$ and $k$ are indices for explanatory variables, $p$ and $q$ are indices for sub-units, and $l'$ and $m'$ are frequency indices; matrix elements $\tilde{M}_{kqm'}$ are Fourier transforms of $M_{kqm}$ along the time axis. The final result is nothing but an application of Parseval's theorem to Equation (31). The expectation values of products of components of $\mathbf{M}$ and $\delta\mathbf{y}$ in Equation (31) are replaced by expectation values of the corresponding products in the frequency domain.

If the Fourier amplitudes are uncorrelated between sub-units and frequency bins, i.e., $\langle \delta\tilde{y}^*_{pl'} \delta\tilde{y}_{qm'} \rangle = 0$ if $l' \neq m'$ or $p \neq q$, the estimated variance of the regression coefficient again takes on a simple form,

$$\operatorname{var}\beta_j = \sum_{p,l'} |\tilde{M}_{jpl'}|^2 P_p(l'), \tag{49}$$

where $P_p(l') \approx |\tilde{y}_{pl'}|^2$ is the estimated power in frequency bin $l'$ for sub-unit $p$. However, one can in principle generalise this frequency-domain estimator to include non-diagonal terms in the frequency-domain noise covariance matrix, e.g., for cross-sectional correlations.

In the case of WLS regression, one has the choice of estimating the power of the unweighted residuals $\boldsymbol{\delta y}$ or the weighted residuals $\boldsymbol{\delta\hat{y}} = \sqrt{\mathbf{W}} \cdot \boldsymbol{\delta y}$. In the latter case, $\mathbf{M}$ and $\boldsymbol{\delta y}$ simply need to be replaced by $\hat{\mathbf{M}} = (\mathbf{X}^\top \cdot \mathbf{W} \cdot \mathbf{X})^{-1} \cdot \mathbf{X}^\top \cdot \sqrt{\mathbf{W}}$ and $\boldsymbol{\delta\hat{y}}$ in the above equations. In the former case, the factor $\sqrt{\mathbf{W}}$ is instead absorbed into $\mathbf{M}$, i.e., $\mathbf{M}$ is replaced by $(\mathbf{X}^\top \cdot \mathbf{W} \cdot \mathbf{X})^{-1} \cdot \mathbf{X}^\top \cdot \mathbf{W}$. In the context of *StopptCOVID*, the weighting factor $\sqrt{\mathbf{W}}$ introduces substantial heteroskedasticity into the residuals. It is therefore preferable to compute confidence intervals based on the unweighted residuals.

#### E. Decision on Error Estimates

It was decided that confidence intervals for the baseline linear regression model should be recalculated using the Driscoll-Kraay sandwich estimator for autocorrelated panel data, bootstrap errors, Ebisuzaki's method as a frequency-domain method, and ARMA$(p, q)$ errors. For all other models a time series bootstrap should be used as the most flexible approach.

#### F. Injection-Recovery Test

The techniques described in Sections S4 A–S4 D all produce uncertainty estimates under the intrinsic assumptions of a model about the deterministic effects and the noise. Based on these assumptions they estimate how model parameter values vary under different noise realisations, or how the likelihood or posterior probability of parameters decreases away from their optimal values. For experiments or observations with a certain well-understood design (e.g., randomised controlled trials), the derived confidence intervals or hypothesis probabilities will relatively accurately represent the true uncertainties.

The situation is different when the deterministic and stochastic processes that shape the observations, and the observational data themselves are more complex, e.g., for models with many covariates and non-linearities, for non-trivial noise processes, or for panel or image data. In such cases, it may not be possible to compute uncertainties rigorously from first principle, as the deterministic and stochastic processes that operate in nature are not exactly known. The intrinsic estimates of uncertainties may approximate the true uncertainties, but this cannot be taken for granted.

In such cases, it is therefore important to gauge the sensitivity and specificity of model estimates by an extrinsic standard as well. To this end, one can test a model by considering its point estimates and uncertainty estimates when confronted with synthetic observables containing a known "injected" signal and some pre-specified level of noise (or possibly no noise at all). The point estimates and error bars "recovered" by a statistical inference model then serve to realistically gauge the sensitivity and specificity of the model and the actual coverage of the error regions estimated within the model. Different types of signals may be injected to study misclassification.

Ideally, one injects real signals from previous, accurate measurements (e.g., electrocardiograms for certain heart conditions), or theoretical predictions from accurate analytic calculations or numerical simulations (e.g., gravitational waveform for inspiralling black holes). Similarly, the noise for the synthetic observations is ideally taken from measurements or a well-tested noise model.

In some contexts, e.g., for the effect of NPIs on the effective reproduction number, the "true" signal and noise model is unknown. In this case, an injection-recovery test is still useful for testing the sensitivity of an inference method to an *assumed* model for the signal (i.e., in our context, the effects of NPIs on $\mathcal{R}(t)$). For example, model $Y$ may determine that a large effect seen by another model $X$ is null with very small error bars, based on the intrinsic signal and noise model of $Y$. But this may simply be due to limitations of model $Y$ that cause it to systematically underestimate this effect. If model $Y$ disagrees with model $X$ on some point estimate or error region for an effect, this has additional force if model $Y$ is in principle capable of recovering this effect. An injection-recovery test can reveal whether this is the case, or whether model $Y$ yields biased estimates if injected with an assumed signal. Note that "bias" in this context only implies bias relative to an assumed model of effects (which may not actually represent nature). Furthermore, an inference model may be biased with respect to its own signal model if it includes shrinkage (see Section S5) to trade smaller variance of the effect estimates for bias.

The concept of injection-recovery test is complementary to other methods for model comparison based on metrics for goodness-of-fit and parsimony like the Akaike information criterion [AIC; 89]. Importantly, it can be applied very generally even when a metric like the AIC cannot be computed or meaningfully compared between different models. This is the case when the underlying error models are incompatible or a likelihood function is not even computed within the model, or when number of (effective) parameters is not readily quantifiable in complex machine learning algorithms. In this case, an injection-recovery test can still provide a means for model comparison.

### S5. APPROACHES FOR HANDLING MULTICOLLINEARITY

This section reviews remedies for multicollinearity in regression models, and specifically in the context of *StopptCOVID*, similar in spirit to our discussion of error analysis in Section S4.

Appropriate methods to address problems with strong multicollinearity are problem-dependent, as is the interpretation of their results. For example, if collinearity between the explanatory variables results from a *known* causal relationship (e.g., between body mass index and blood pressure), one or more of the highly correlated variables may be dropped based upon a heuristic approach that takes these known interdependencies into account. If multicollinearity results from a *possible, but yet unknown* causal relationship between the explanatory variables (e.g., between biomarkers influenced by a yet unknown disease), dimensionality reduction may be applied to identify the underlying causal factors, project the explanatory variables onto them and discard dimensions in the space of explanatory variables that may be regarded as "noise" as they do not explain substantial variance in the data.

Finally, in the context of NPI evaluations, one faces multicollinearity between accidentally correlated explanatory variables (practically) without any intrinsic causal relationship, e.g., mask mandates and school closures do not intrinsically influence each other and can in principle be implemented at will. Here, multicollinearity effectively results from a bad experimental design that does not generate enough data points to allow, as much as possible, for a *ceteris paribus* evaluation of individual NPIs (or carefully designed bundles of NPIs).[13] Under these circumstances, methods for dimensionality reduction, feature selection and/or regularisation can at best hope to identify groups of NPIs with similar activation patterns for assigning them some joint effect, pragmatically identify candidates for "important" NPIs based on more or less heuristic criteria, and attempt to avoid overfitting. In this case, it is ultimately impossible to rectify multicollinearity without introducing bias as a trade-off (bias-variance trade-off).

Before reviewing possible remedies for multicollinearity in the *StopptCOVID* data set and in NPI studies more broadly, it is useful to present the problem from a slightly different angle. Note that the following discussion only deals with multicollinearity in linear regression.

[13] Even greater care with the experimental setup is required to detect possible (non-linear) interactions between NPI effects on $\mathcal{R}(t)$, which, by construction, cannot even be captured in a linear model that treats the effects as independent from each other.

### A. Relation to Singular Values of the Design Matrix

Least-squares linear regression is tantamount to finding an approximate solution to an overdetermined system of equation, $\mathbf{y} = \mathbf{X} \cdot \boldsymbol{\beta}$ with more observations $N_{\text{obs}}$ than regression parameters $N_{\text{reg}}$. For the purpose of this section, it is useful to reformulate this problem and its solution using the singular value decomposition (SVD) of the matrix $\mathbf{X}$,

$$\mathbf{X} = \mathbf{U} \cdot \mathbf{S} \cdot \mathbf{V}^\top, \tag{50}$$

where $\mathbf{U}$ and $\mathbf{V}$ are orthogonal[14] $N_{\text{obs}} \times N_{\text{obs}}$ and $N_{\text{reg}} \times N_{\text{reg}}$ matrices. $\mathbf{S}$ is a rectangular diagonal $N_{\text{obs}} \times N_{\text{reg}}$ matrix,

$$\mathbf{S} = \begin{pmatrix} \lambda_1 & 0 & 0 & 0 \\ 0 & \lambda_2 & 0 & 0 \\ 0 & 0 & \ddots & \vdots \\ 0 & 0 & \dots & \lambda_{N_{\text{reg}}} \\ 0 & 0 & 0 & 0 \\ \vdots & \vdots & \vdots & \vdots \end{pmatrix}, \tag{51}$$

where $\lambda_1, \dots, \lambda_{N_{\text{reg}}}$ are the singular values of the design matrix, conventionally sorted in descending order. Using the SVD, the overdetermined problem $\mathbf{y} = \mathbf{U} \cdot \mathbf{S} \cdot \mathbf{V}^\top \cdot \boldsymbol{\beta}$ can be written as $\mathbf{U}^\top \mathbf{y} = \mathbf{S} \cdot (\mathbf{V}^\top \cdot \boldsymbol{\beta})$, which amounts to decoupled equations of the transformed variables $\hat{\boldsymbol{\beta}} = \mathbf{V}^\top \cdot \boldsymbol{\beta}$ and $\hat{\mathbf{y}} = \mathbf{U}^\top \cdot \mathbf{y}$,

$$\lambda_i \hat{\beta}_i = \hat{y}_i. \tag{52}$$

This has the obvious solution[15]

$$\hat{\beta}_i = \lambda_i^{-1} \hat{y}_i, \tag{53}$$

or in terms of components of $\boldsymbol{\beta}$ and $\mathbf{y}$,

$$\beta_i = \sum_{j=1}^{N_{\text{reg}}} \sum_{k=1}^{N_{\text{obs}}} V_{ij} \lambda_j^{-1} U_{kj} y_k. \tag{54}$$

Equation (53,54) nicely serve to illustrate the repercussions of multicollinearity. Strong multicollinearity implies that some of the singular values $\lambda_i$ are significantly smaller than $\lambda_1$. The $\hat{y}_i$ with small $\lambda_i$ (and the patterns in the observations corresponding to them) have high influence on

[14] This implies that $\mathbf{U}^\top$ and $\mathbf{V}^\top$ are the inverses of $\mathbf{U}$ and $\mathbf{V}$, respectively.

[15] Note that this solution is, of course, mathematically identical to Equation (28).

the regression coefficients $\beta_i$. Small amounts of noise $\delta\hat{y}_i$ in the transformed observations $\hat{y}_i$ can tilt the regression coefficients considerably away from their true values because the error $\delta\hat{\beta}_i$ gets inflated by $\lambda_i^{-1}$,

$$\delta\hat{\beta}_i = \lambda_i^{-1}\delta\hat{y}_i. \tag{55}$$

The patterns in the data corresponding to these problematic small singular values are often those most strongly affected by intrinsic or observational noise, i.e., the $\hat{y}_i$ with low singular values are often determined by high-frequency noise or strongly sensitive to a few influential data points.

#### B. Truncated SVD Regression and Related Methods

One possibility to avoid the large errors associated with small singular values is to simply discard the corresponding components of the solution or of the regressors (dimensionality reduction); this is known as as truncated SVD regression [90, 91] or non-centred principal component[16] regression. The process of setting certain $\lambda_i$ to zero can be viewed in different, but mathematically equivalent ways. Truncated SVD/principal component regression is commonly interpreted as performing regression using the transformed explanatory variables (principal components) $\hat{X}_i = \sum_{i'} V_{ii'} X_{i'}$ and discarding some of these. For certain problems, these transformed explanatory variables may be interpreted as latent variables that capture real phenomena hidden the explanatory variables. The rationalisation for such an interpretation is that the transformed explanatory variables provide a series of optimal least-square approximations to the original explanatory variables. In the case of NPIs, these principal components could be viewed as characteristic modes of activating or not activating certain NPIs.

"Reification" of the principal components is problematic on scientific grounds in some cases, however. Moreover the transition to transformed explanatory variables in truncated SVD regression is not required. One can just as well view the process as the application of a filter to the observed data whereby some of the transformed observations $\hat{y}_i$ are set to zero by a projection operator $\mathbf{P}$ before transforming back,

$$\mathbf{y} \to \mathbf{U} \cdot \mathbf{P} \cdot \mathbf{U}^\top \cdot \mathbf{y}. \tag{56}$$

[16] Centring is inappropriate in the case of NPIs because it affects the interpretability of the regression coefficients. If an NPI variable $X_i$ is changed in a counterfactual scenario, this will also affect its average and hence the centring. The difference in $\mathcal{R}$ between two cases with $X_i = 0$ and $X_i = 1$ is therefore not solely given by the regression coefficient in centred PCR regression, but also depends on the different centring in both cases.

Finally, one can simply view truncated SVD regression as shrinking (components of) the effect estimates by modifying $\lambda_i$ in Equation (55).

If truncated SVD/principal component regression is viewed as a transformation to a reduced set of new explanatory variables, this transformation may be suboptimal in the sense that it does not take into account any correlations of the initial set of explanatory variables with the response variable. This is remedied in partial least squares regression [92], which sometimes achieves better regression results for a given number of retained components.

#### C. Regularisation using Penalty Terms

Very unstable estimates of certain coefficients $\hat{\beta}_i$ in the case of multicollinearity can also be understood from the shape of the likelihood function for linear regression,

$$\ln \mathcal{L} = \frac{1}{2}(\mathbf{X} \cdot \boldsymbol{\beta} - \mathbf{y}) \cdot (\mathbf{X} \cdot \boldsymbol{\beta} - \mathbf{y}). \tag{57}$$

Unstable estimates $\hat{\beta}_i$ for small singular values $\lambda_i$ result from the shallowness of the paraboloid function $\ln \mathcal{L}$ in these directions. One approach to achieve more stable estimates is therefore to add penalty terms of the likelihood. The prototype for such a *regularisation* with penalty terms is Tikhonov regularisation, also known as ridge regression [93, 94], which adds a quadratic penalty term in $\boldsymbol{\beta}$,

$$\ln \mathcal{L} = \frac{1}{2}(\mathbf{X} \cdot \boldsymbol{\beta} - \mathbf{y}) \cdot (\mathbf{X} \cdot \boldsymbol{\beta} - \mathbf{y}) + \frac{\Gamma}{2}\boldsymbol{\beta} \cdot \boldsymbol{\beta}, \tag{58}$$

where $\Gamma$ is a tunable parameter. The penalty term modifies the normal equations,

$$\mathbf{X}^\top \cdot \mathbf{X} \cdot \boldsymbol{\beta} - \mathbf{X}^\top \cdot \mathbf{y} + \Gamma\boldsymbol{\beta} = 0, \tag{59}$$

or upon inserting the singular value decomposition $\mathbf{X} = \mathbf{U} \cdot \mathbf{S} \cdot \mathbf{V}^\top$ and transforming to $\hat{\boldsymbol{\beta}}$ and $\hat{\mathbf{y}}$,

$$(\mathbf{S}^2 + \Gamma\mathbf{I}) \cdot \hat{\boldsymbol{\beta}} - \mathbf{S} \cdot \hat{\mathbf{y}} = 0. \tag{60}$$

One can hence cast the solution for the regression coefficients into the form,

$$\hat{\beta}_i = \frac{\lambda_i}{\lambda_i^2 + \Gamma}\hat{y}_i, \tag{61}$$

which can be compared to the non-regularised solution (55). For $\Gamma \ll \lambda_i^2$, one recovers the standard solution, whereas (components of the) regression coefficients with small singular values and $\Gamma \gg \lambda_i^2$ are suppressed. Equation (61) illustrates the relation of Tikhonov regularisation to truncated

SVD regression; instead of an abrupt cutoff of components with small singular values, Tikhonov regularisation works with a smoothly varying suppression factor.

Different regularisation methods for regression are in use; e.g., LASSO (Least Absolute Shrinkage and Selection Operator) regression [95] instead adds a penalty term proportional to the absolute value $|\boldsymbol{\beta}|$. Elastic net regression [96] combines the Tikohonov and LASSO penalty terms,

$$\ln \mathcal{L} = \frac{1}{2}(\mathbf{X}\cdot\boldsymbol{\beta} - \mathbf{y})\cdot(\mathbf{X}\cdot\boldsymbol{\beta} - \mathbf{y}) + \frac{\Gamma}{2}\boldsymbol{\beta}\cdot\boldsymbol{\beta} + \Theta|\boldsymbol{\beta}|, \tag{62}$$

where $\Theta$ is another tunable parameter. The choice between different forms of the penalty terms can to some extent be addressed by heuristics for model selection (see below), but is generally problem-dependent. Some characteristic features of the different methods can be identified, however. Tikohonv regularisation tends to attribute effects more evenly to heavily correlated explanatory variables, whereas LASSO tends to set some regression coefficients to zero (implicit feature selection) and attributes bigger effects to the remaining ones.

#### D. Other Methods to Address Multicollinearity

Other types of methods for mitigating the problem of multicollinearity (e.g., by feature selection) are also in use in the literature. We do not attempt to survey these exhaustively, but note that some of these methods come with significant caveats. Feature selection by *stepwise regression* has a long history [97]. This approach relies on adding or deleting variables based on whether this significantly (as quantified statistical tests) improves or degrades the fit. Such techniques are employed in some NPI studies [98]; but stepwise regression techniques are broadly considered as problematic because of a substantial risk of bias away from the null (to the point of postulating significant effects from pure noise), too narrow confidence intervals, and the problematic use of multiple significance tests [99–102].

#### E. Determination of Tunable Parameters

All methods for handling multicollinearity involve a trade-off between sacrifing some *in-sample* accuracy of the fit and introducing an explicit bias towards the null in exchange for smaller confidence intervals for the regression parameters and, hopefully, more accurate parameter estimates *in practice*. Deciding whether the shrinkage of the regression parameters merely filters out unwanted errors or also filters out true effects is therefore delicate. The ultimate test is model validation on new and truly independent data, which are typically unavailable.

In lieu of such true out-of-sample data, one can resort either to various information criteria such as the Akaike information criterion [AIC; 89] or to mock out-of-sample data using hold-out validation or cross validation. Likelihood-based information criteria like the AIC face a subtle issue with time series data, however, as the likelihood cannot be computed assuming independent errors at each data point and must take the error covariance matrix into account.[17] Packages for this purpose may not be readily available.

Alternatively, one can split the data into a training set and a validation set (hold-out validation) or perform multiple fits leaving out different parts of the data (cross-validation), and then assess the goodness of fit on these mock out-of-sample validation data. As with bootstrapping, these techniques need to be adjusted when serial correlation is present. A possible solution for cross-validation consists in a time series split that partitions the data, e.g., into $N$ blocks and $N$ different partitions into training and validation data, or variations thereof [103]. The $n$-th training set consists of the first $n$ blocks, and the remaining block constitute the corresponding validation data.

*a. Decision:* It was decided to implement both truncated SVD (principal component) regression and elastic net regression as regularised alternatives to the baseline regression model. The implementation of the selected methods is outlined in Section S6.

### S6. TECHNICAL DESCRIPTION OF MODEL SET

#### Standard Linear Regression Models with Different Error Estimators

**Model DK** uses Driscoll-Kraay errors [69], which are directly available in `statsmodels`. We follow [104] in choosing the maximum lag for the estimation of error covariances as $[4(N_t/100)^{2/9}]$, which is also used in Stata [70]. Noting that these lags are often too small [70], we also tested a maximum lag of 90 d to capture low-frequency variations in the residuals, but did not find substantial differences.

**Model Ebisuzaki** implements a frequency-domain method based on [88]. The method is formulated as a plug-in estimator for confidence intervals based on the power spectra of the un-

[17] This problem does not arise when autocorelated errors are explicitly included and the likelihood is correctly computed based on the pointwise innovations.

weighted residuals and the matrix $\mathbf{M} \cdot \mathbf{W}$ (where $\mathbf{M}$ is the pseudo-inverse of the design matrix $\mathbf{X}$, and $\mathbf{W}$ is the weight matrix). A detailed derivation is provided in Supplementary Methods S4. The power spectra are estimated using the Fast Fourier Transform (FFT). To avoid contamination by edge effects (which may in particular add power at low frequencies), we use reflection padding, i.e., we construct truly periodic data by attaching a mirrored version of the respective time series. The Fourier transform then has twice the original frequency resolution and is downsampled by merging frequency bins in pairs. Windowing is another means of dealing with edge effects in power spectrum estimation [e.g., 105, 106], but can be problematic in cases where features close to the beginning and end of the time series contribute significant power [107], as is the case for the *StopptCOVID* data (cp. Figure 1).

**Model BT** computes bootstrap errors using the `arch` package [108]. We use a stationary bootstrap with exponentially distributed block sizes [82]. The average block size is determined based on the autocorrelation structure of the residuals [109, 110]. The same sequence of bootstrapped dates is used for all states, and we resample only in time and not among states. Alternative choices for reasmpling do not significantly affect the error estimates (Supplementary Methods S8). Error bars at 95% confidence level are computed from the standard deviation of the parameters in 500 bootstrap samples. Time series bootstrap errors are also used for models 2WFE, DYN, RF, Elastic Net, and PCR. For the more expensive models DYN and RF, we use a smaller number of 100 samples, however.

#### Linear regression with two-way fixed effects

**Model 2WFE** implements regression with two-way fixed effects as another means for incorporating unmodelled time-dependent processes, see, e.g., Section 13.3.3 in [1] and [111]. This approach can be viewed as an extension of the difference-in-differences method. In its basic form, regression with two-way fixed effects adds time-dependent fixed-effects $\gamma_t$ to Equation (8),

$$y_{j,t} = \alpha_j + \gamma_t + \sum_{\substack{i \\ \text{(non-seasonal)}}} \beta_i X_{j,t,i} + \epsilon_{j,t}, \tag{63}$$

where the sum now runs only over those (non-seasonal) regressors that depend on time *and* entity. Two-way fixed effects enjoy considerable popularity in econometrics [112] and are also represented in the literature on NPIs [e.g., 113]. Like all other methods, regression with two-way fixed effects is subject to limitations and, in certain cases, prone to biases [112, 114, 115]. For example, in the case of NPI evaluation two-way fixed effects would by construction fail to attribute any effects to NPIs in the extreme case of complete mixing between geographical entities. Two-way fixed effects in their most simple form can no longer produce effect estimates for seasonality and events with fixed dates (Easter and Christmas). This, however, can be remedied by hierarchical inference. To obtain effect estimates for the cosine and sine components of seasonality and the Easter and Christmas season, we simply regress the fixed effects in terms of these three explanatory variables $X_{t,i}$,

$$\gamma_t = \alpha + \sum_{\substack{i \\ \text{(seasonal)}}} \beta_i X_{t,i} + \epsilon_t, \tag{64}$$

where the sum runs over the (seasonal) regressors that depend *only* on time. Regression with two-way fixed effects is combined with a time-series bootstrap as explained above. Since two-way fixed effects regression subtracts dynamics common across entities, one expects the residuals to be more weakly correlated across the federal states. Hence, asynchronous resampling appears to be the most appropriate strategy for the bootstrap, and is used by default. For the three seasonal regressors, we perform a case bootstrap on the fixed effects $\gamma_t$. Note that computing the errors in $\gamma_t$ from the first step of the two regression steps are not propagated to the computation of errors for the seasonal term this way. The downstream impact of errors in $\gamma_t$ can be checked by directly sampling the distribution of the seasonal effect estimates under the bootstrap for the first regression step. These errors were found to be much smaller than those estimated from the second step under a case bootstrap.

#### Linear Regression with ARMA Errors

**Model ARMA($p, q$)** uses regression with ARMA errors, i.e., the uncorrelated error terms $\epsilon_{j,t}$ in Equation (1) are replaced by autocorrelated noise $n_{j,t}$,

$$\ln \mathcal{R}_{j,t} = \alpha_j + \sum_i \beta_i X_{j,t,i} + n_{j,t}. \tag{65}$$

Here $n_{j,t}$ is determined by an ARMA process of order $(p, q)$,

$$n_{j,t} - \sum_{\tau=1}^{p} \phi_\tau n_{j,t-\tau} = \epsilon_{j,t} + \sum_{\tau=1}^{q} \theta_\tau \epsilon_{j,t-\tau}, \tag{66}$$

with autoregression coefficient $\phi_\tau$ and $\theta_\tau$, and $\epsilon_{j,t}$ are uncorrelated innovations. Regression with ARMA errors is implemented in Python using `statsmodels.statespace` [116]. Harvey's representation of ARMA($p, q$) processes in state-space form is used [117], i.e., Equation (66) for the noise $n_{j,t+}$ is written as the state vector equation in the state-space representation with the help of auxiliary variables $\zeta^{(2)}_{j,t+1}, \ldots, \zeta^{(r)}_{j,t+1}$, where $r = \max(p, q+1)$,

$$\begin{pmatrix} n_{j,t} \\ \zeta^{(2)}_{j,t+1} \\ \zeta^{(3)}_{j,t+1} \\ \vdots \\ \zeta^{(r)}_{j,t+1} \end{pmatrix} = \begin{pmatrix} \phi_1 & 1 & 0 & \ldots & 0 \\ \phi_2 & 0 & 1 & \ldots & 0 \\ \phi_3 & 0 & 0 & \ddots & 0 \\ \vdots & \vdots & \vdots & 0 & 1 \\ \phi_r & 0 & 0 & 0 & 0 \end{pmatrix} \cdot \begin{pmatrix} n_{j,t} \\ \zeta^{(2)}_{j,t} \\ \zeta^{(3)}_{j,t} \\ \vdots \\ \zeta^{(r)}_{j,t} \end{pmatrix} + \epsilon_t \begin{pmatrix} 1 \\ \theta_1 \\ \theta_2 \\ \vdots \\ \theta_r \end{pmatrix}, \tag{67}$$

where the innovations $\epsilon_{j,t}$ are normally distributed, $\epsilon_{j,t} \sim \mathcal{N}(0, \sigma^2)$. The coefficients $\phi_l$ and $\theta_l$ are set to zero for $l > p$ and $l > q$, respectively. Equation (65) for the response variable is written as the observation equation of the state-space model using a design matrix $(1, 0, \ldots, 0)^\top$, and the regression terms and fixed effects are added as intercept of the observations,

$$\ln \mathcal{R}_{j,t} = \begin{pmatrix} 1 \\ 0 \\ 0 \\ \vdots \\ 0 \end{pmatrix} \cdot \begin{pmatrix} n_{j,t} \\ \zeta^{(2)}_{j,t} \\ \zeta^{(3)}_{j,t} \\ \vdots \\ \zeta^{(r)}_{j,t} \end{pmatrix} + \alpha_j + \sum_i \beta_i X_{j,t,i}. \tag{68}$$

For reasons of computational efficiency, the time series for individual states are concatenated. At the end of each time series, the transition matrix is set to zero so that the error $n_{j,0}$ and the auxiliary variables $\zeta^{(l)}_{j,0}$ are zero at the beginning of the next time series. Similarly, $n_{j,0}$ and the auxiliary variables $\zeta^{(l)}_{j,0}$ are set to zero in the first state by specifying the initial state of the model. The intercept for the observations for time index $t = 0$ is set to the observed value $y_{j,0}$ in each state, so that this data point does not influence the parameter estimates. This approach reduces the dimension of the state vector to $\max(p, q+1)$ instead of $N_s \max(p, q+1)$ for a panel model, and hence considerably speeds up execution in `statsmodels.statespace` (due to the non-sparse representation of `numpy` matrices).

We also make one further modification to the naive ARMA noise model in order to avoid problems with noise estimation at very low frequencies by introducing a frequency cut-off. We reset the auxiliary variables $\zeta^{(l)}_{j,0}$ to zero every 109 time steps (one fifth of the time series) by zeroing the transition matrix. This effectively cuts the noise time series into five independent chunks for each state. This modification ensures that the model does not interpret the low-frequency component of $\mathcal{R}(t)$ (which reflects the non-stationarity of $\mathcal{R}(t)$ during the initial phase of the pandemic) as

ARMA noise. The idea of spectral estimation based on locally stationary segments has been used more broadly in the literature, and can be generalised to more sophisticated techniques to estimate (time-varying) spectral properties of non-stationary time series using adaptive segmentation [118–121].

Confidence intervals for the regression coefficients are computed using the outer product of gradients approximation [71] (see also Section S4 B). The order $(p, q)$ of the ARMA errors is determined based on the Bayesian Information Criterion (BIC, [122]); for applications of information criteria to time series analysis, see, e.g., [106]. For a grid of models with $p \leq 7$ and $q \leq 13$, we obtain a minimum BIC for $(p, q) = (1, 11)$. However, the point estimates and confidence intervals for most regression coefficients exhibit little variation for orders higher than $(p + q) \gtrsim 6$.

#### Dynamical Model – Discrete Renewal Equation

**Model DYN** is a dynamical model based on a renewal equation. In formulating this model, we remain as close as possible to the regression model used in *StopptCOVID* to ensure that the inferred effect sizes have exactly the same interpretation as in the baseline model, and that the same input data can be used. Combining the definition of $\mathcal{R}_{j,t}$ (Equation 3) and the linear regression model for $\ln \mathcal{R}_{j,t}$ from Equation (1) immediately leads to a renewal equation,

$$\bar{\mathcal{I}}_{j,t} = \mathcal{R}_{j,t}\bar{\mathcal{I}}_{j,t-4} = \exp\Bigg\{\alpha_j + 0.3\nu_{\alpha,t} + 0.6\nu_{\delta,t} + \beta_0 \cos\frac{2\pi t}{365\,\mathrm{d}} + \beta_1 \sin\frac{2\pi t}{365\,\mathrm{d}} - \beta_2 \log_2[1 - V(t - \tau_{\mathrm{vac}})] + \sum_{i=3}^{N_{\mathrm{NPI}}+2} \beta_i X_{j,i}(t)(t - \tau_{\mathrm{NPI}})\Bigg\}\bar{\mathcal{I}}_{j,t-4}, \tag{69}$$

for the *smoothed* case data $\bar{\mathcal{I}}_{j,t}$,

$$\bar{\mathcal{I}}_{j,t} = \frac{1}{7}\sum_{\tau=0}^{6} \bar{\mathcal{I}}_{j,t-\tau}. \tag{70}$$

Such a renewal equation for incident cases $\mathcal{I}$ can be viewed as a discrete version of an integro-differential age-of-infection model along the lines of the original Kermack-McKendrick theory [35]. In certain limiting cases (e.g., for certain infectivity functions or when $\mathcal{R}$ varies slowly), such an integro-differential model can be converted into an equivalent differential equation model of the SIR family [see, e.g., 123]. Equation (69) corresponds most closely, but not exactly to an SIR model under the assumption that the depletion of susceptibles can be neglected (i.e., $S \approx 1$). This approximate correspondence is briefly outlined in Section S7. Note that formulating a renewal equation for the smoothed case data $\bar{\mathcal{I}}$ instead of the daily case data $\mathcal{I}$ modifies the character of the renewal equation and the relation between $\mathcal{R}_{j,t}$ and the time-dependent transmission probability $T(t, \tau)$, but a rigorous analysis of this issue is deferred to Work Package 2. Again, the issue is briefly outlined in Appendix S7.

We again use `statsmodels.statespace` to estimate this model in the form

$$\bar{\mathcal{I}}_{j,t} = \mathcal{R}_{j,t}\bar{\mathcal{I}}_{j,t-4} + \epsilon_{j,t-1}, \tag{71}$$

with errors $\epsilon_{j,t-1}$. These error are to be understood not as observational errors, but as daily fluctuations of the *actual* infections, and will therefore influence case numbers after time $t$ as well. For the purpose of model estimation, the errors are assumed to be independent, but it is critical to model them as heteroskedastic, i.e., as having time-dependent variance. The variance of the fluctuations will be larger when case numbers are high. If daily infections were determined by a Poisson process, one would expect a variance corresponding to daily case numbers; in reality complex infection dynamics such as clustering of infections will generally lead to a larger variance. However, one still expects the variance to *scale* with $\mathcal{I}$, and we therefore use normally distributed errors with $\epsilon_{j,t} \sim \mathcal{N}(0, \sigma^2\bar{\mathcal{I}}_{j,t})$. The choice of normally distributed errors is to make the estimation of the parameters in the state-space formulation computationally efficient based on standard Python packages; MLE with non-normal errors would require substantial algorithmic changes. The normality assumption is no longer accurate for small case numbers, but periods with low case numbers do not critically influence MLE results anyway.

The dynamical model is built completely into the state equation of the state-space model,

$$\begin{pmatrix} \bar{\mathcal{I}}_{j,t+1} \\ \bar{\mathcal{I}}_{j,t} \\ \bar{\mathcal{I}}_{j,t-1} \\ \bar{\mathcal{I}}_{j,t-2} \end{pmatrix} = \begin{pmatrix} 0 & 0 & 0 & \mathcal{R}_{j,t+1} \\ 1 & 0 & 0 & 0 \\ 0 & 1 & 0 & 0 \\ 0 & 0 & 1 & 0 \end{pmatrix} \cdot \begin{pmatrix} \bar{\mathcal{I}}_{j,t} \\ \bar{\mathcal{I}}_{j,t-1} \\ \bar{\mathcal{I}}_{j,t-2} \\ \bar{\mathcal{I}}_{j,t-3} \end{pmatrix} + \begin{pmatrix} \epsilon_{j,t} \\ 0 \\ 0 \\ 0 \end{pmatrix}, \tag{72}$$

where $\mathcal{R}_{j,t}$ is computed according to Equation (1). The observation equation is trivial; the design matrix is simply $(1, 0, 0, 0)^\top$. As for regression with ARMA errors, the time series for the different states are patched together; the transition matrix is zeroed at the end of each time series, and the correct number of cases for the first day of the time series is enforced by appropriately specifying the initial conditions for the state vector or the state intercept.

When computing confidence intervals for the NPI effect sizes, accounting for autocorrelated errors of a dynamical model requires considerable care. We choose a time series bootstrap, which is still relatively easy to implement (albeit computationally costly), and allows sanity checks on the simulated observational data generated by the resampling process. First, the bootstrap needs to take into account that the errors are heteroskedastic, which can be dealt with by rescaling (standardising) the residuals before reshuffling them [124, 125]. For example, one could resample the rescaled residuals $\delta\hat{\bar{\mathcal{I}}}_{j,t}$,

$$\delta\hat{\bar{\mathcal{I}}}_{j,t} = \frac{1}{\bar{\mathcal{I}}_{j,t}^{1/2}}\left(\mathcal{R}_{j,t}\bar{\mathcal{I}}_{j,t-4} - \bar{\mathcal{I}}_{j,t}\right), \tag{73}$$

and then construct the simulated data $\bar{\mathcal{I}}^{\mathrm{sim}}_{j,t}$ as

$$\bar{\mathcal{I}}^{\mathrm{sim}}_{j,t} = \bar{\mathcal{I}}_{j,t} + \bar{\mathcal{I}}^{1/2}_{j,t}\delta\hat{\bar{\mathcal{I}}}_{j,\mathcal{S}(t)}, \tag{74}$$

where $\mathcal{S}(t)$ denotes the time indices of the reshuffled residuals. This can, however, violate a second requirement on the simulated data, viz., that case numbers must be positive.

We therefore resample the logarithmic difference between the predicted cases $\bar{\mathcal{I}}^{\mathrm{pred}}_{j,t} = \mathcal{R}_{j,t}\bar{\mathcal{I}}_{j,t-4}$ and the observed cases. When the residuals are small, the logarithmic difference $\delta\ln\bar{\mathcal{I}}_{j,t} = \ln(\bar{\mathcal{I}}^{\mathrm{pred}}_{j,t}/\bar{\mathcal{I}}_{j,t})$ corresponds to the relative (one-step) prediction error, and its variance scales roughly with $\bar{\mathcal{I}}^{-1}_{j,t}$. After scaling, rearranging and unscaling $\delta\ln\bar{\mathcal{I}}_{j,t}$, the simulated data are obtained as

$$\bar{\mathcal{I}}^{\mathrm{sim}}_{j,t} = \bar{\mathcal{I}}_{j,t}\, e^{\bar{\mathcal{I}}^{-1/2}_{j,t}\left(\bar{\mathcal{I}}^{1/2}\,\delta\ln\bar{\mathcal{I}}\right)_{j,\mathcal{S}(t)}}. \tag{75}$$

This guarantees that the simulated data remain positive. The use of $\delta\ln\bar{\mathcal{I}}$ as opposed to $\delta\bar{\mathcal{I}}$ will lead to a small upward bias in the simulated case data, but his effect is minute unless case numbers are very small, and the case numbers are not the quantity of interest anyway. Compared to the alternative procedure of simply limiting the bootstrapped residuals in Equation (74) to ensure positivity of the simulated data, we found no substantial differences.

#### Random Forest Regression

**Model RF** employs random forest regression [126] and has been implemented in Python using `sklearn` [127]. Random forest regression was chosen because this has been implemented previously by several studies of NPI effects [39, 40]. Another advantage consists in relatively low training costs, often with little or no loss of accuracy compared to more expensive methods [128], which also expedites uncertainty quantification by bootstrap methods and hyperparameter optimisation. The potentially higher interpretability of the decision trees in random forest regression compared to, e.g., neural networks is only a secondary consideration, as we do not exploit this feature in practice.

We run random forest regression with squared error minimisation in the tree-splitting steps. Different from the linear regression models, we do *not* include dummy variables for states for the reasons discussed above. There is a concern that mixing state dummy variables and NPI indicator variables in decision trees is not epidemiologically meaningful. Furthermore, given the large number of explanatory variables, we believe that it is advisable not to waste any levels of the decision trees on dummy variables for states. Tests indicated that dropping dummy variables for states has no major impact on effect estimates, and still permits (nominally) smaller fit errors than linear regression with fixed effects for states.

The number of trees (between 12 and 200), their maximum depth (between 2 and 19), and the number of features considered in each split (between 1 and 18) are optimised using cross validation on a coarse parameter grid. A time series split with 5 splits is used for cross validation; it is critical that no random reshuffling of individual data points is applied during cross validation when working on autocorrelated time series data. Note that the estimates of effect sizes are relatively robust except for extreme choices for the hyperparameters.

For the best-fit model (100 trees, maximum depth 13, maximum of 4 features per split), we estimate error bars using a stationary bootstrap on the *case data*. Different from the case of linear regression, a case bootstrap is required because the large number of parameters implicit in random forest regression leads to a risk of overfitting and can reduce the residuals to very small values for a sufficiently large number of trees and depth of trees. Bootstrapping of residuals would severely underestimate the actual parameter errors in this case.

The extraction of linear effect sizes from random forest regression requires care and is subject to ambiguities that cannot be fully resolved. A key requirement is that the effect size extracted from an arbitrary dependence of $\ln\mathcal{R}$ on the NPI variables,

$$\ln\mathcal{R} = \mathcal{R}(X_1, X_2, X_3 \ldots) \tag{76}$$

must reduce to the (linear) regression coefficients if the functional dependence *is* actually linear. This can be achieved by noting that for a linear model, the regression coefficient can be obtained as the difference in $\Delta\ln\mathcal{R}_i$ when NPI $i$ switched on $X_i = 1$ compared to when it is switched off

$X_i = 0$, if all other NPIs and other explanatory variables are the same in both cases,

$$\beta_i = \Delta \ln \mathcal{R}_i = \mathcal{R}(X_0, \ldots, X_{i-1}, 1, X_{i+1}, \ldots, X_{N_{\mathrm{NPI+2}}}) \\ - \mathcal{R}(X_0, \ldots, X_{i-1}, 0, X_{i+1}, \ldots, X_{N_{\mathrm{NPI+2}}}). \tag{77}$$

Equation (77) holds at any time and in any state for a linear model. For a non-linear model, $\Delta \ln \mathcal{R}$ will depend on the state of the other NPIs. The natural choice for defining an interpretable linear effect size for a non-linear model is thus a weighted average over $\ln \mathcal{R}_{j,t,i}$ (now explicitly expressed as depending on time $t$ and geographical entity $j$),

$$\beta_i = \frac{\sum_{j,t} w_{j,t} \Delta \ln \mathcal{R}_{j,t,i}}{\sum_{j,t} w_{j,t}}, \tag{78}$$

where $w_{j,t}$ is a weight function. We use Equation (78) to define effect sizes for individual NPIs in the case of random forest regression, and apply the same weights as for the baseline model. This set of effect sizes is labelled "RF1".

Alternatively, one can, e.g., consider the effect of switching on NPI $i$ while all other NPIs are switched off in the model, but the seasonal features remain switched on,

$$\Delta \ln \mathcal{R}_{j,t,i} = \mathcal{R}(\cos \frac{2\pi t}{365\,\mathrm{d}}, \sin \frac{2\pi t}{365\,\mathrm{d}}, 0, \ldots, 0, 1, 0, \ldots, 0) \\ - \mathcal{R}(\cos \frac{2\pi t}{365\,\mathrm{d}}, \sin \frac{2\pi t}{365\,\mathrm{d}}, 0, \ldots, 0, 0, 0, \ldots, 0). \tag{79}$$

The set of putative effects sizes in the absence of all other interventions is labelled "RF0".

#### Linear Regression with Shrinkage

We implement two regression methods with shrinkage to address the effects of multicollinearity in the NPIs.

**Model PCR** implements principal component regression (PCR) using principal component analysis (PCA) of the non-standardised data without demeaning (which might be more precisely called truncated SVD regression). PCA is performed using `statsmodels` on the NPI variables only, and not on the fixed effects for different states as the interpretation of transformed explanatory variables and regression coefficients that mix fixed effects and treatment effects would be somewhat problematic. The optimal choice of principal components is determined by cross validation using a time series split with five blocks. The optimal number of components is found to be 13.

**Model Elastic Net** uses the routine for elastic net regression [96] in `scikit-learn` [127]. As a minor modification of the baseline model, we use a universal intercept $\alpha$ instead of fixed effects for different states,

$$y_{j,t} = \alpha + \sum_i \beta_i X_{j,t,i} + \epsilon_{j,t}. \tag{80}$$

The rationale for dropping fixed effects for states is to prevent the LASSO penalty term to zero *some* of the fixed effects for states but not others, which would potentially skew NPI effect estimates considerably. Since the fixed effects for states are relatively similar (Supplementary Table I), replacing the fixed effects with a single intercept does not substantially degrade goodness-of-fit. As for principal component regression, we perform cross validation with a time series split to optimise the choice for the Tikhonov and LASSO penalty terms. The model grid for cross validation covers 21 values from $10^{-4}$ to $10^{-5}$ for the sum $\Gamma + \Theta$ of the LASSO and Tikhonov term, and values for $\Theta/(\Theta + \Gamma)$ of 0.001, 0.002, 0.01, 0.05, 0.1, 0.2, 0.3, 0.4, 0.5, 0.6, 0.7, 0.8, 0.9, 0.95, and 0.99 to vary the ratio between the two terms.

### S7. FORMULATION OF THE *STOPPTCOVID* MODEL AS A RENEWAL EQUATION

In this section, we provide some technical discussion on the formulation of the *StopptCOVID* model as a discrete dynamical model. We discuss the relation between the renewal equation to integro-differential and differential equation models for disease spread, and comment on the implication of the use of rolling averages of case numbers instead of daily case numbers.

#### A. Relation to SIR Model

The renewal equation,

$$\mathcal{I}_t = \mathcal{R}_t \mathcal{I}_{t-4}, \tag{81}$$

with a time-dependent reproduction number can be formulated as an integro-differential equation of Kermack-McKendrick type by introducing the age distribution $\mathcal{I}(t, \tau)$ of cases by time $t$ and age of infection $\tau$. The Kermack-McKendrick equations [35] for $\mathcal{I}(t, \tau)$ and the susceptible fraction $S$ become

$$\frac{\partial \mathcal{I}(t, \tau)}{\partial t} + \frac{\partial \mathcal{I}(t, \tau)}{\partial \tau} = \delta(\tau)\, \lambda(t)\, S(t), \tag{82}$$

$$\frac{\mathrm{d}S(t)}{\mathrm{d}t} = -\lambda(t)\, S(t), \tag{83}$$

where $\lambda(t)$ is the force of infection at time $t$, and $\delta$ denotes the Dirac delta function. After infections at time $t$ have occurred at time $t$ and age of infection $\tau$, one simply has $\mathcal{I}(t,\tau) = \mathcal{I}(t-\tau,0) = \mathcal{I}_t$ for the correspondence between the continuous function $\mathcal{I}(t,\tau)$ and the discrete case data $\mathcal{I}_t$. The force of infection is given by

$$\lambda(t) = \int_0^\infty T(t,\tau)\mathcal{I}(t,\tau)\,\mathrm{d}\tau, \tag{84}$$

in terms of a time-dependent infectivity function $T(t,\tau)$. Equations (69) and (81) for the discrete renewal equation are recovered in the limit $S = 1$ (i.e., negligible depletion of susceptibles) and for an infectivity function with a $\delta$-function peak, $T(t,\tau) = \mathcal{R}_t\delta(\tau - 4\,\mathrm{d})$.

Equation (82) can be converted into the differential equations for the SIR model as follows.[18] Let $\tau_{\text{inf}}$ be the maximum infectious period, and define the fractions $I$ and $R$ of infectives and recovered individuals as

$$I = \int_{0^-}^{\tau_{\text{inf}}^+} \mathcal{I}(t,\tau)\,\mathrm{d}\tau, \quad R = \int_{\tau_{\text{inf}}^-}^{\infty} \mathcal{I}(t,\tau)\,\mathrm{d}\tau, \tag{85}$$

i.e., as the fractions of individuals with an age of infection smaller or larger than $\tau_{\text{inf}}$. Here the superscipted symbols + and − are used to indicate whether or not $\delta$-function peaks at the boundaries of the domain of integration are included or not. Integrating Equation (82) over the corresponding intervals yields

$$\frac{\partial}{\partial t}\int_{0^-}^{\tau_{\text{inf}}^+} \mathcal{I}(t,\tau)\,\mathrm{d}\tau + \int_{0^-}^{\tau_{\text{inf}}^+} \frac{\partial \mathcal{I}(t,\tau)}{\partial \tau}\,\mathrm{d}\tau = S(t)\int_{0^-}^{\tau_{\text{inf}}^+} \delta(\tau)\lambda(t)\,\mathrm{d}\tau, \tag{86}$$

or,

$$\frac{\partial I}{\partial t} + \mathcal{I}(t,\tau_{\text{inf}}) = S(t)\int_{0^-}^{\tau_{\text{inf}}^+} T(t,\tau)\mathcal{I}(t,\tau)\,\mathrm{d}\tau. \tag{87}$$

If $\mathcal{I}$ varies *slowly* (i.e., $\tau_{\text{inf}}\,\partial\mathcal{I}/\partial t \ll \mathcal{I}$), we have $\mathcal{I}(t,\tau) \approx I/\tau_{\text{inf}}$, and hence

$$\frac{\partial I}{\partial t} = \frac{\int_{0^-}^{\tau_{\text{inf}}^+} T(t,\tau)\,\mathrm{d}\tau}{\tau_{\text{inf}}} S(t)I(t) - \frac{I(t)}{\tau_{\text{inf}}}, \tag{88}$$

which has the form of the differential equation for $I$ in the standard SIR model. Alternatively, if the reproduction number $\mathcal{R}$ varies slowly, we can approximate $\mathcal{I}(t,\tau) = \mathcal{I}(t-\tau,0) \approx \mathcal{I}(t,0)\exp(-\omega\tau)$ with a growth rate $\omega = \tau_{\text{gen}}^{-1}\ln\mathcal{R}$. In this case, one can solve the integral for $I(t)$ from Equation (85)

[18] The reformulation as differential equations can also proceed slightly differently, and without the approximations used here, if an exponential distribution of the infectious period is assumed.

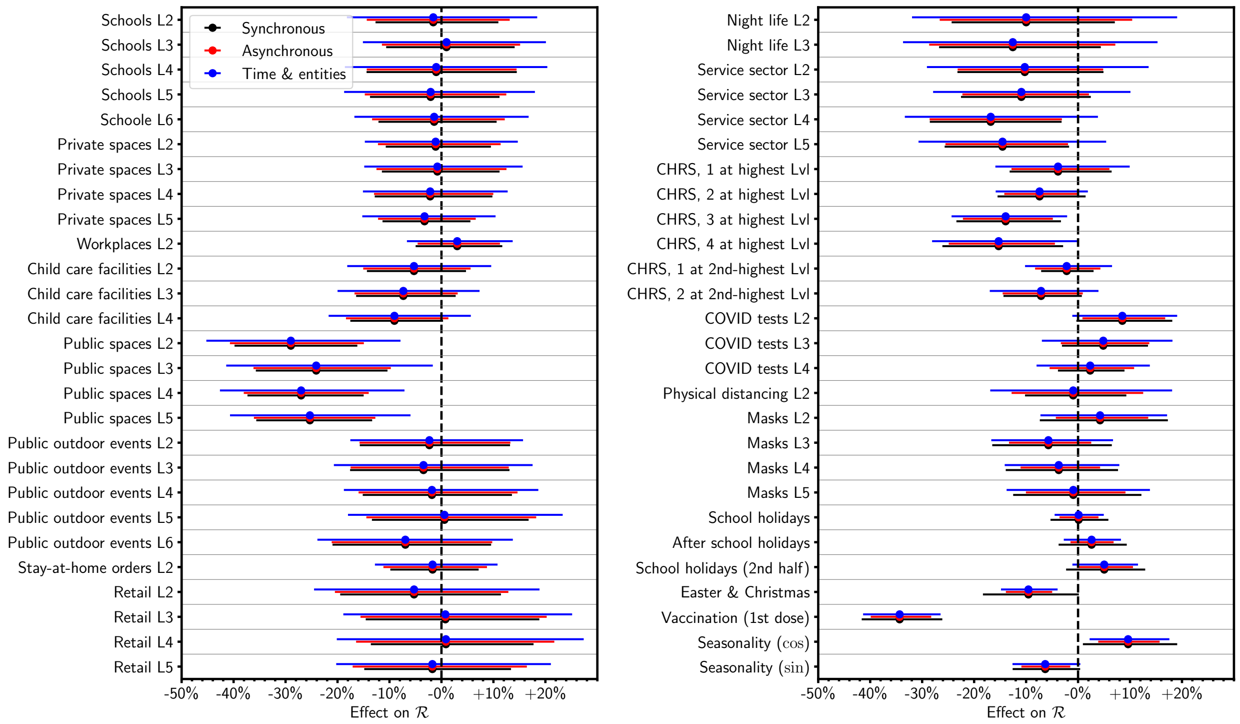


FIG. 5. Sensitivity of bootstrap confidence intervals to the resampling method (black: synchronous resampling across all states, red: asynchronous resampling for each state, blue: asynchronous resampling of residuals and resampling of the bootstrapped residuals across entities (federal states).

analytically, and then obtain $\mathcal{I}(t,\tau)$ in terms of $I(t)$. Equation (87) then becomes

$$\frac{\partial I}{\partial t} = \frac{\omega \int_{0^-}^{\tau_{\text{inf}}^+} T(t,\tau) e^{-\omega\tau}\, \mathrm{d}\tau}{1 - e^{-\omega\tau_{\text{inf}}}} S(t)\, I(t) - \frac{\omega}{e^{\omega\tau_{\text{inf}}} - 1} I(t), \tag{89}$$

which is again just the differential equation for $I$ in the standard SIR model. Note that the rate $\omega$ and $\mathcal{R}$ are related to the infectivity function $T$; the exact relation is not considered important for our purpose here.

#### B. Impact of Rolling Averaging in *StopptCOVID* Case Data

*StopptCOVID* computes the reproduction number from rolling averages of the daily case numbers. As a result, their model and its formulation as a renewal equation $\bar{\mathcal{I}}_t = \mathcal{R}_t \bar{\mathcal{I}}_{t-4}$ are in fact equivalent to

$$\sum_{\tau=0}^{6} \mathcal{I}_{t-\tau} = \mathcal{R}_t \sum_{\tau=4}^{10} \mathcal{I}_{t-\tau}. \tag{90}$$

This can formally be cast as a renewal equation,

$$\mathcal{I}_t = \mathcal{R}_t \sum_{\tau=4}^{10} \mathcal{I}_{t-\tau} - \sum_{\tau=1}^{6} \mathcal{I}_{t-\tau}, \tag{91}$$

but as the second sum on the right-hand-side is negative, this no longer has a direct meaningful epidemiological interpretation. However, the deviation from the desired form of the renewal equation $\mathcal{I}_t = \mathcal{R}_t \mathcal{I}_{t-4}$ can be made more explicit using the concept of slow variation of $\mathcal{R}_t$ as in Section S7 A. Equation (91) can be written as

$$\mathcal{I}_t = \mathcal{R}(t)\mathcal{I}_{t-4} + \sum_{\tau=1}^{6} (\mathcal{R}_t - \mathcal{R}_{t-\tau})\mathcal{I}_{t-\tau-4}. \tag{92}$$

If the temporal variations in the *actual* $\mathcal{R}_t$ are small over the time scale of a week, using rolling averages of case numbers instead of daily case numbers only introduces a minor perturbation of the desired renewal equation. The actual amount of bias introduced by the rolling average will be analysed in the next work packages.

### S8. RESAMPLING STRATEGY FOR BOOTSTRAP ERRORS

Figure 5 addresses the sensitivity of the estimated confidence intervals to the resampling method used for the time series bootstrap. We consider the three methods already outlined in Section S4 C:

- For *synchronous resampling*, *all* time indices are replaced by elements of the *same* time series bootstrap sequence $\tau(t)$, i.e., the bootstrapped residuals are $\delta y_{j,\tau(t)}$.
- For *asynchronous* resampling, we use a different bootstrap sequence $\tau_j(t)$ for each entity $j$, i.e., the bootstrapped residuals are $\delta y_{j,\tau_j(t)}$.
- For resampling both in time and across entities, the residual from state $j$ is replaced with that from state $\sigma(j)$ *and* independent resampling in time is applied, i.e., the resampled residuals are $\delta y_{\sigma(j),\tau_j(t)}$.

Figure 5 shows that most confidence intervals do not depend heavily on the choice of resampling method. Asynchronous resampling actually *narrows* some confidence intervals, in particular for seasonality, school holidays and vaccination. This gives some additional credence to ranking the associations with these explanatory variables as significant. We stress that none of these resampling methods is a priori better than the others. The key point for this study is that they are consistent at a level that does not qualitatively affect our findings.

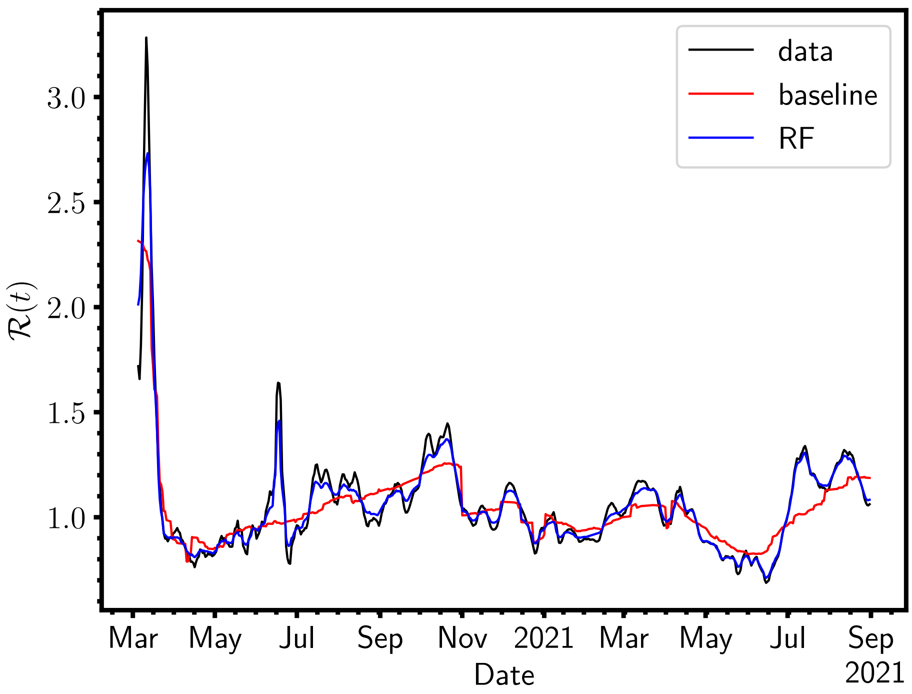


FIG. 6. Fit of $\mathcal{R}(t)$ (national average) from random forest regression (blue) compared to the national case data (black) and the fit for the baseline model (red). Note that model RF reproduces the temporal dynamics quite closely and significantly better than the linear regression model, despite nominally small linear effect sizes for the explanatory variables.

### S9. SUPPLEMENTARY DISCUSSION OF RANDOM FOREST REGRESSION

In this section, we illustrate some peculiarities of random forest regression that complicate the interpretation of the inferred NPI effect sizes, most of which tend to be quite small. To illustrate that the model does not underfit, we compare the nationally averaged $\mathcal{R}(t)$-curve to the random forest fit in Supplementary Figure 6. Random forest regression clearly fits the observed data better than the linear regression model.

To illustrate why this is not reflected in the linear effect estimates, we consider how the model prediction changes as selected NPIs or groups of NPIs are switched on, taking the state of Bavaria as an example in Figure 7. We first consider the case when only seasonal features (school holidays, after school holidays, school holidays (2nd half), Easter & Christmas, seasonality) and vaccination are switched on in the model. In this case, $\ln \mathcal{R}(t)$ already drops to about 0.4 after the first outbreak in Spring 2020, and exhibits a non-trivial time dependence that bears little resem-

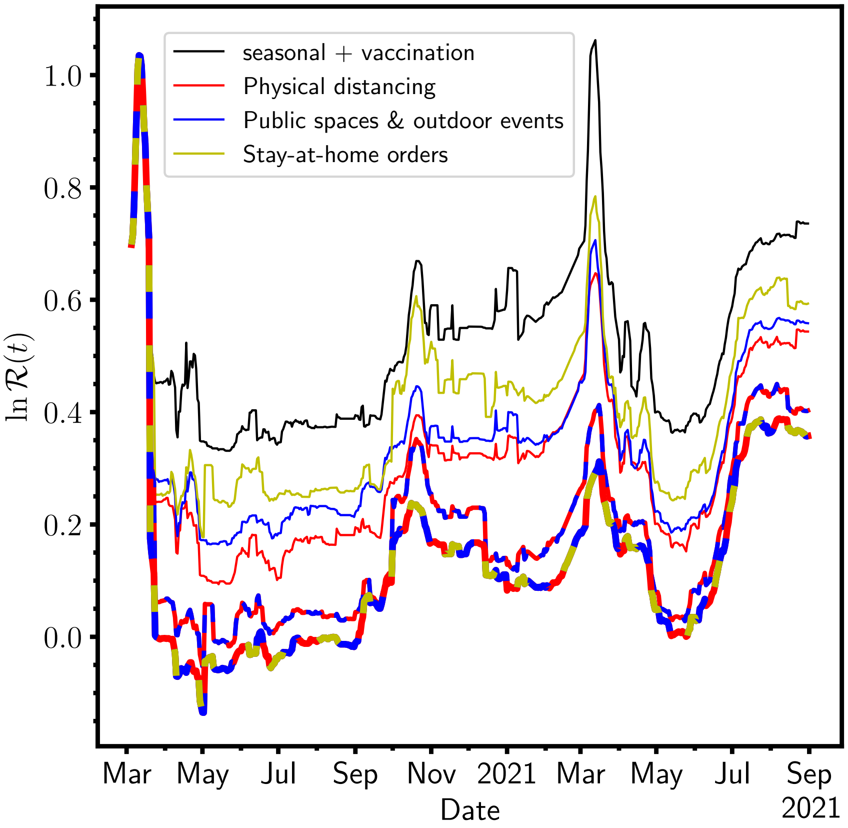


FIG. 7. Predicted $\ln \mathcal{R}(t)$ using random forest regression for various counterfactual scenarios in the state of Bavaria. The black curve only includes seasonal features and vaccination while all NPIs are set as inactive. Red solid and dashed curves show the effect of including physical distancing in the prediction whenever it was mandated in Bavaria. Similarly, blue colour indicates that active NPIs for *public spaces* and public outdoor events are included in the prediction, and yellow indicates that stay-at-home orders are included. Thicker lines are used for the counterfactual scenarios that combine these NPIs.

blance to the seasonal explanatory variables. While unintuitive, this behaviour is easily explained. As soon as a sine and cosine term for seasonality are included as explanatory variables, decision-tree based algorithms can effectively construct arbitrary functions of the phase angle $\varphi$ by using two-dimensional step functions in $\cos\varphi$ and $\sin\varphi$. Thus, random forest regression implicitly allows for almost arbitrary NPI-independent temporal background dynamics similar to regression with two-way fixed effects. The only restriction is that the background dynamics must still have annual periodicity.

We then separately switch on NPIs for i) physical distancing, ii) the whole group of NPIs for public spaces and public outdoor events. Any of these (groups of), and iii) stay-at-home orders

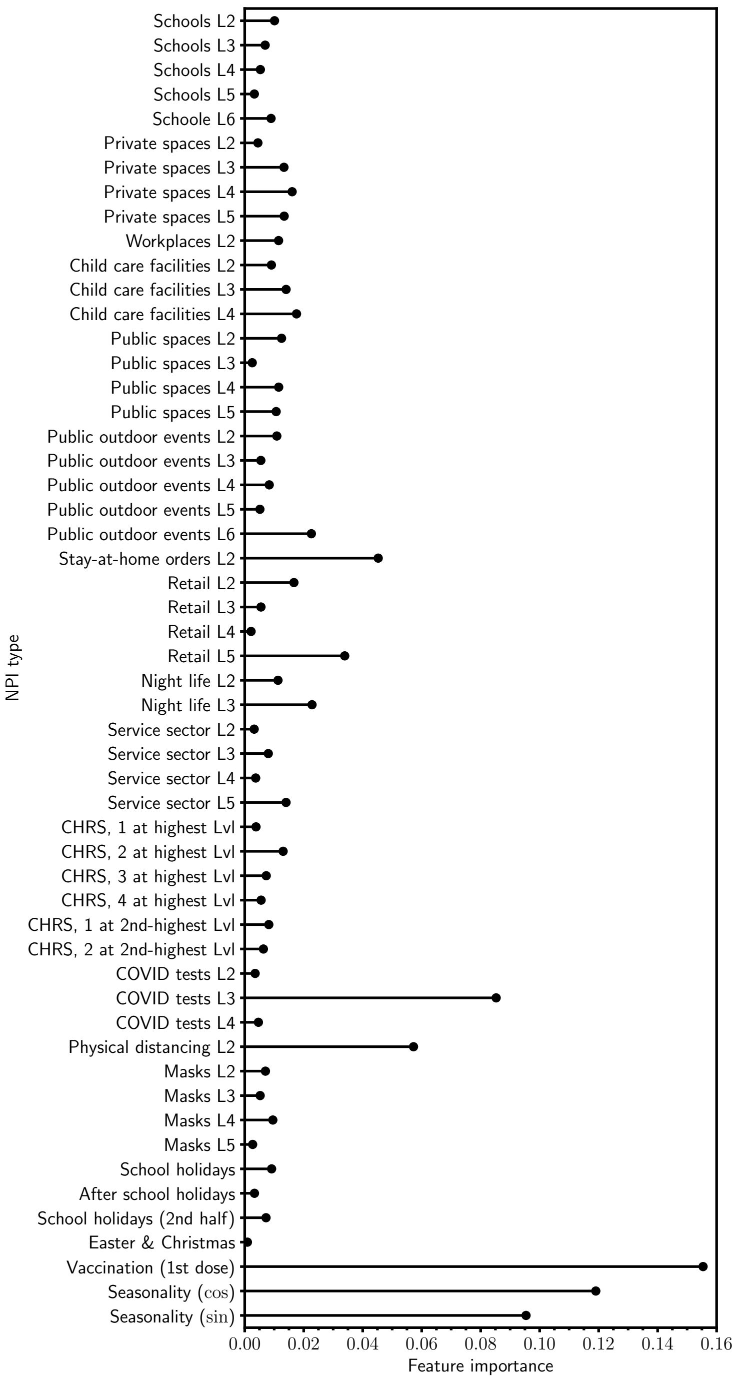


FIG. 8. Feature importance in the random forest regression model for NPIs, seasonal terms, and vaccination.

and NPIs reduce $\ln\mathcal{R}(t)$ by about another 0.25 when no other NPIs are switched on. Stay-at-home orders have the weakest effect, but still lead to a substantial reduction in the model.

When NPIs for physical distancing, public spaces and public outdoor events are combined, the effect on $\ln\mathcal{R}(t)$ is visibly less than the sum of the two individual effects during some periods.

When stay-at-home orders are added, this generally has much less of an effect in the model than when they are switched on without the other NPIs. In other words, the model "sees" a saturation effect when additional NPIs are added, which is absent by construction in linear regression. Whether this saturation effect is real or merely a convenient description of the data would have to be determined by other means, however.

Faced with these non-trivial interactions of interventions implicit in random forest regression, feature importance (Gini importance) is a useful quantity to consider. Feature importance quantifies how much certain explanatory variables contribute on average to the total reduction of the summed squared error (or any other metric used for minimisation). Feature importance by construction sum up to unity and are always positive. They must not be confused with effect size estimates, but nonetheless give an indication of the relevance of explanatory variables within the model.

Feature importances are shown in Figure 8. Vaccination and seasonality are ranked as most important by this criterion. This is a further indication that vaccination (in the short term) and NPI-independent temporal dynamics substantially influence the epidemic trajectory.

Methods for partitioning *effect sizes* in the presence of interactions exists, e.g., based on Shapley values [129, 130]. However, such standard methods also encounter difficulties. In our concrete example the Shapley values (evaluated using the SHAP package [131]) counterintuitively assign a small *increase* of $\mathcal{R}(t)$ to vaccination before vaccination even starts. This just means that the model sees a vaccination fraction of zero as an indicator of an earlier time period with higher $\mathcal{R}(t)$, but evidently introduces subtleties into the interpretation of the Shapley values. More refined approaches to extract interpretable effect sizes from complex machine learning models for NPI evaluation and in other, similar contexts will be a fruitful topic for further research, but are beyond the scope of this paper.

TABLE III. Estimated effect sizes and confidence intervals

| Regressor | DK | BT | Ebisuzaki | 2WFE | ARMA(6,11) | DYN | RF1 | RF0 | elastic net | PCR |
|---|---|---|---|---|---|---|---|---|---|---|
| Schools L2 | -0.02 | -0.02 | -0.02 | -0.01 | **-0.06** | 0.08 | 0.01 | -0.08 | 0.00 | **-0.03** |
| | (-0.11,0.07) | (-0.14,0.10) | (-0.17,0.14) | (-0.11,0.08) | **(-0.09,-0.03)** | (0.06,0.10) | (-0.00,0.02) | (-0.18,0.03) | (-0.05,0.05) | **(-0.06,-0.00)** |
| Schools L3 | 0.01 | 0.01 | 0.01 | 0.00 | **-0.06** | 0.10 | -0.01 | -0.02 | -0.01 | **-0.03** |
| | (-0.08,0.10) | (-0.11,0.13) | (-0.14,0.16) | (-0.09,0.10) | **(-0.09,-0.03)** | (0.08,0.12) | (-0.01,0.00) | (-0.08,0.03) | (-0.05,0.02) | **(-0.06,-0.00)** |
| Schools L4 | -0.01 | -0.01 | -0.01 | -0.01 | **-0.06** | 0.08 | **-0.01** | -0.07 | -0.04 | **-0.05** |
| | (-0.11,0.09) | (-0.16,0.14) | (-0.18,0.16) | (-0.10,0.09) | **(-0.10,-0.03)** | (0.06,0.11) | **(-0.02,-0.00)** | (-0.17,0.02) | (-0.08,0.01) | **(-0.08,-0.03)** |
| Schools L5 | -0.02 | -0.02 | -0.02 | -0.00 | **-0.06** | 0.07 | -0.00 | -0.01 | -0.04 | **-0.02** |
| | (-0.11,0.07) | (-0.15,0.11) | (-0.18,0.14) | (-0.10,0.09) | **(-0.10,-0.03)** | (0.05,0.09) | (-0.01,0.01) | (-0.04,0.03) | (-0.08,0.01) | **(-0.03,-0.01)** |
| Schoole L6 | -0.01 | -0.01 | -0.01 | -0.01 | **-0.05** | 0.07 | -0.00 | -0.01 | **-0.03** | **-0.02** |
| | (-0.10,0.07) | (-0.13,0.10) | (-0.16,0.13) | (-0.10,0.08) | **(-0.08,-0.03)** | (0.05,0.09) | (-0.02,0.01) | (-0.08,0.05) | **(-0.06,-0.00)** | **(-0.04,-0.00)** |
| Private spaces | -0.01 | -0.01 | -0.01 | -0.01 | -0.01 | 0.00 | -0.00 | -0.01 | -0.02 | **-0.04** |
| L2 | (-0.06,0.04) | (-0.11,0.09) | (-0.14,0.12) | (-0.08,0.06) | (-0.03,0.01) | (-0.03,0.03) | (-0.01,0.01) | (-0.06,0.03) | (-0.06,0.02) | **(-0.05,-0.02)** |
| Private spaces | -0.01 | -0.01 | -0.01 | -0.03 | **-0.02** | 0.00 | 0.01 | 0.01 | -0.02 | **-0.03** |
| L3 | (-0.07,0.05) | (-0.12,0.11) | (-0.14,0.13) | (-0.11,0.04) | **(-0.04,-0.00)** | (-0.02,0.03) | (-0.01,0.03) | (-0.04,0.06) | (-0.07,0.02) | **(-0.06,-0.00)** |
| Private spaces | -0.02 | -0.02 | -0.02 | -0.03 | **-0.03** | -0.00 | -0.01 | **-0.12** | **-0.05** | **-0.07** |
| L4 | (-0.09,0.04) | (-0.14,0.09) | (-0.15,0.11) | (-0.10,0.04) | **(-0.04,-0.01)** | (-0.04,0.03) | (-0.03,0.00) | **(-0.24,-0.00)** | **(-0.10,-0.01)** | **(-0.10,-0.04)** |
| Private spaces | -0.03 | -0.03 | -0.03 | -0.03 | **-0.03** | -0.03 | -0.01 | -0.12 | **-0.04** | -0.02 |
| L5 | (-0.08,0.02) | (-0.12,0.05) | (-0.14,0.08) | (-0.09,0.03) | **(-0.05,-0.01)** | (-0.05,0.00) | (-0.02,0.00) | (-0.27,0.02) | **(-0.08,-0.00)** | (-0.04,0.00) |
| Workplaces L2 | 0.03 | 0.03 | 0.03 | 0.00 | -0.01 | 0.02 | -0.00 | -0.05 | -0.02 | **-0.03** |
| | (-0.01,0.07) | (-0.05,0.11) | (-0.05,0.11) | (-0.04,0.05) | (-0.03,0.01) | (0.01,0.04) | (-0.02,0.01) | (-0.11,0.01) | (-0.05,0.01) | **(-0.06,-0.00)** |
| Child care | **-0.05** | -0.05 | -0.05 | 0.01 | **-0.07** | **-0.05** | 0.01 | -0.04 | -0.03 | **-0.05** |
| facilities L2 | **(-0.10,-0.01)** | (-0.15,0.05) | (-0.18,0.07) | (-0.06,0.08) | **(-0.10,-0.04)** | **(-0.07,-0.03)** | (-0.01,0.02) | (-0.09,0.02) | (-0.09,0.02) | **(-0.07,-0.03)** |
| Child care | **-0.08** | -0.08 | -0.08 | -0.01 | **-0.07** | **-0.08** | -0.01 | -0.12 | **-0.06** | **-0.04** |
| facilities L3 | **(-0.13,-0.02)** | (-0.18,0.03) | (-0.20,0.05) | (-0.07,0.06) | **(-0.10,-0.04)** | **(-0.10,-0.06)** | (-0.02,0.00) | (-0.27,0.02) | **(-0.10,-0.01)** | **(-0.07,-0.00)** |
| Child care | **-0.09** | -0.09 | -0.09 | -0.01 | **-0.05** | **-0.10** | **-0.01** | 0.00 | -0.05 | **-0.06** |
| facilities L4 | **(-0.15,-0.04)** | (-0.19,0.00) | (-0.22,0.03) | (-0.07,0.06) | **(-0.08,-0.02)** | **(-0.12,-0.08)** | **(-0.03,-0.00)** | (-0.07,0.07) | (-0.11,0.01) | **(-0.10,-0.03)** |
| Public spaces L2 | **-0.34** | **-0.34** | **-0.34** | 0.02 | **-0.09** | **-0.36** | **-0.03** | -0.13 | **-0.11** | **-0.02** |
| | **(-0.48,-0.21)** | **(-0.51,-0.18)** | **(-0.53,-0.16)** | (-0.11,0.14) | **(-0.12,-0.06)** | **(-0.40,-0.32)** | **(-0.06,-0.01)** | (-0.26,0.01) | **(-0.18,-0.04)** | **(-0.03,-0.01)** |
| Public spaces L3 | **-0.28** | **-0.28** | **-0.28** | 0.02 | **-0.06** | **-0.31** | 0.00 | 0.00 | -0.01 | **-0.03** |
| | **(-0.42,-0.13)** | **(-0.44,-0.11)** | **(-0.46,-0.09)** | (-0.11,0.14) | **(-0.09,-0.04)** | **(-0.34,-0.27)** | (-0.01,0.01) | (-0.03,0.03) | (-0.08,0.05) | **(-0.04,-0.02)** |

TABLE III. Estimated effect sizes and confidence intervals (ctd.)

| Regressor | DK | BT | Ebisuzaki | 2WFE | ARMA(6,11) | DYN | RF1 | RF0 | elastic net | PCR |
|---|---|---|---|---|---|---|---|---|---|---|
| Public spaces L4 | **-0.31** | **-0.31** | **-0.31** | 0.02 | **-0.08** | **-0.34** | -0.00 | -0.09 | -0.05 | **-0.04** |
| | **(-0.46,-0.17)** | **(-0.47,-0.16)** | **(-0.49,-0.14)** | (-0.09,0.14) | **(-0.10,-0.05)** | **(-0.37,-0.31)** | (-0.03,0.02) | (-0.21,0.04) | (-0.13,0.03) | **(-0.07,-0.01)** |
| Public spaces L5 | **-0.29** | **-0.29** | **-0.29** | 0.04 | **-0.06** | **-0.31** | -0.01 | -0.08 | -0.06 | **-0.07** |
| | **(-0.43,-0.15)** | **(-0.44,-0.14)** | **(-0.46,-0.13)** | (-0.07,0.14) | **(-0.08,-0.04)** | **(-0.34,-0.28)** | (-0.03,0.01) | (-0.17,0.00) | (-0.14,0.01) | **(-0.10,-0.03)** |
| Public outdoor | -0.02 | -0.02 | -0.02 | 0.00 | **-0.02** | **-0.04** | 0.01 | 0.10 | 0.00 | **-0.05** |
| events L2 | (-0.10,0.05) | (-0.17,0.12) | (-0.20,0.15) | (-0.10,0.11) | **(-0.04,-0.01)** | **(-0.08,-0.01)** | (-0.00,0.03) | (-0.01,0.21) | (-0.06,0.06) | **(-0.08,-0.02)** |
| Public outdoor | -0.04 | -0.04 | -0.04 | -0.00 | -0.00 | **-0.05** | 0.01 | -0.02 | -0.03 | -0.01 |
| events L3 | (-0.11,0.04) | (-0.19,0.12) | (-0.21,0.14) | (-0.12,0.11) | (-0.03,0.02) | **(-0.08,-0.02)** | (-0.00,0.03) | (-0.06,0.02) | (-0.08,0.01) | (-0.02,0.01) |
| Public outdoor | -0.02 | -0.02 | -0.02 | -0.00 | -0.01 | **-0.05** | 0.01 | -0.01 | -0.01 | **-0.02** |
| events L4 | (-0.10,0.06) | (-0.16,0.13) | (-0.19,0.15) | (-0.11,0.11) | (-0.03,0.01) | **(-0.06,-0.03)** | (-0.01,0.02) | (-0.07,0.06) | (-0.05,0.02) | **(-0.04,-0.00)** |
| Public outdoor | 0.01 | 0.01 | 0.01 | 0.03 | **-0.08** | -0.01 | 0.00 | -0.04 | -0.01 | **-0.04** |
| events L5 | (-0.07,0.08) | (-0.14,0.16) | (-0.17,0.18) | (-0.08,0.14) | **(-0.10,-0.06)** | (-0.04,0.01) | (-0.01,0.02) | (-0.09,0.02) | (-0.04,0.03) | **(-0.05,-0.02)** |
| Public outdoor | -0.07 | -0.07 | -0.07 | -0.01 | **-0.05** | **-0.10** | **-0.02** | -0.06 | **-0.04** | **-0.05** |
| events L6 | (-0.16,0.02) | (-0.24,0.09) | (-0.25,0.11) | (-0.12,0.10) | **(-0.07,-0.03)** | **(-0.12,-0.08)** | **(-0.04,-0.00)** | (-0.13,0.01) | **(-0.07,-0.01)** | **(-0.08,-0.02)** |
| Stay-at-home | -0.02 | -0.02 | -0.02 | -0.02 | **-0.11** | **-0.02** | **-0.03** | -0.20 | -0.02 | **-0.09** |
| orders L2 | (-0.04,0.01) | (-0.10,0.07) | (-0.13,0.09) | (-0.08,0.03) | **(-0.12,-0.09)** | **(-0.04,-0.00)** | **(-0.05,-0.00)** | (-0.41,0.01) | (-0.05,0.02) | **(-0.13,-0.05)** |
| Retail L2 | -0.05 | -0.05 | -0.05 | 0.03 | **-0.03** | **-0.03** | 0.01 | -0.10 | -0.05 | 0.00 |
| | (-0.20,0.09) | (-0.22,0.11) | (-0.25,0.14) | (-0.07,0.13) | **(-0.06,-0.00)** | **(-0.05,-0.00)** | (-0.01,0.02) | (-0.22,0.03) | (-0.15,0.04) | (-0.04,0.04) |
| Retail L3 | 0.01 | 0.01 | 0.01 | 0.04 | **-0.03** | 0.04 | 0.01 | -0.02 | -0.03 | **-0.04** |
| | (-0.14,0.15) | (-0.16,0.17) | (-0.18,0.19) | (-0.05,0.14) | **(-0.06,-0.00)** | (0.01,0.06) | (-0.01,0.02) | (-0.06,0.01) | (-0.09,0.03) | **(-0.07,-0.01)** |
| Retail L4 | 0.01 | 0.01 | 0.01 | 0.06 | -0.01 | 0.05 | 0.00 | -0.01 | -0.04 | **-0.03** |
| | (-0.14,0.16) | (-0.15,0.16) | (-0.19,0.21) | (-0.03,0.16) | (-0.04,0.02) | (0.02,0.08) | (-0.01,0.01) | (-0.04,0.01) | (-0.09,0.01) | **(-0.04,-0.01)** |
| Retail L5 | -0.02 | -0.02 | -0.02 | 0.04 | **-0.05** | 0.02 | **-0.03** | **-0.19** | **-0.07** | **-0.09** |
| | (-0.16,0.13) | (-0.16,0.13) | (-0.19,0.16) | (-0.05,0.13) | **(-0.07,-0.02)** | (-0.00,0.04) | **(-0.05,-0.01)** | **(-0.35,-0.03)** | **(-0.11,-0.02)** | **(-0.12,-0.07)** |
| Night life L2 | -0.11 | -0.11 | -0.11 | **-0.15** | **-0.05** | **-0.19** | -0.00 | -0.04 | -0.08 | **-0.06** |
| | (-0.33,0.12) | (-0.28,0.07) | (-0.33,0.12) | **(-0.29,-0.01)** | **(-0.08,-0.01)** | **(-0.22,-0.15)** | (-0.02,0.01) | (-0.11,0.02) | (-0.15,0.00) | **(-0.09,-0.03)** |
| Night life L3 | -0.13 | -0.13 | -0.13 | **-0.16** | -0.03 | **-0.21** | -0.01 | **-0.12** | **-0.10** | **-0.10** |
| | (-0.35,0.09) | (-0.31,0.04) | (-0.36,0.09) | **(-0.30,-0.02)** | (-0.06,0.00) | **(-0.24,-0.18)** | (-0.03,0.00) | **(-0.24,-0.00)** | **(-0.17,-0.03)** | **(-0.14,-0.07)** |

TABLE III. Estimated effect sizes and confidence intervals (ctd.)

| Regressor | DK | BT | Ebisuzaki | 2WFE | ARMA(6,11) | DYN | RF1 | RF0 | elastic net | PCR |
|---|---|---|---|---|---|---|---|---|---|---|
| Service sector | -0.11 | -0.11 | -0.11 | -0.06 | -0.02 | **-0.04** | 0.00 | -0.02 | -0.02 | **-0.02** |
| L2 | (-0.28,0.07) | (-0.26,0.05) | (-0.29,0.07) | (-0.16,0.05) | (-0.06,0.01) | **(-0.07,-0.01)** | (-0.00,0.01) | (-0.06,0.01) | (-0.09,0.04) | **(-0.03,-0.01)** |
| Service sector | -0.12 | -0.12 | -0.12 | -0.03 | -0.01 | **-0.05** | 0.01 | -0.06 | -0.05 | -0.03 |
| L3 | (-0.30,0.07) | (-0.26,0.02) | (-0.27,0.04) | (-0.13,0.07) | (-0.03,0.02) | **(-0.07,-0.03)** | (-0.01,0.02) | (-0.14,0.02) | (-0.11,0.01) | (-0.06,0.00) |
| Service sector | **-0.18** | **-0.18** | **-0.18** | -0.07 | -0.02 | **-0.11** | -0.01 | -0.04 | **-0.10** | **-0.06** |
| L4 | **(-0.37,-0.00)** | **(-0.34,-0.03)** | **(-0.35,-0.01)** | (-0.17,0.04) | (-0.05,0.01) | **(-0.14,-0.09)** | (-0.02,0.01) | (-0.08,0.01) | **(-0.19,-0.01)** | **(-0.07,-0.04)** |
| Service sector | -0.16 | **-0.16** | **-0.16** | -0.05 | -0.01 | **-0.09** | -0.01 | -0.17 | **-0.09** | **-0.05** |
| L5 | (-0.34,0.02) | **(-0.30,-0.02)** | **(-0.31,-0.00)** | (-0.15,0.05) | (-0.03,0.02) | **(-0.11,-0.07)** | (-0.03,0.01) | (-0.36,0.01) | **(-0.16,-0.02)** | **(-0.08,-0.03)** |
| CHRS, 1 at | -0.04 | -0.04 | -0.04 | 0.02 | 0.02 | **-0.04** | -0.00 | 0.03 | -0.04 | **-0.02** |
| highest Lvl | (-0.10,0.02) | (-0.14,0.06) | (-0.15,0.07) | (-0.04,0.08) | (0.00,0.03) | **(-0.07,-0.00)** | (-0.01,0.01) | (-0.04,0.10) | (-0.09,0.02) | **(-0.03,-0.01)** |
| CHRS, 2 at | **-0.08** | -0.08 | -0.08 | 0.02 | 0.01 | **-0.07** | 0.01 | -0.02 | -0.05 | **-0.06** |
| highest Lvl | **(-0.13,-0.03)** | (-0.17,0.01) | (-0.16,0.00) | (-0.03,0.07) | (-0.01,0.03) | **(-0.10,-0.04)** | (-0.02,0.03) | (-0.11,0.08) | (-0.12,0.02) | **(-0.07,-0.04)** |
| CHRS, 3 at | **-0.15** | **-0.15** | **-0.15** | -0.00 | 0.00 | **-0.15** | -0.01 | -0.06 | **-0.11** | **-0.02** |
| highest Lvl | **(-0.21,-0.09)** | **(-0.27,-0.03)** | **(-0.26,-0.04)** | (-0.08,0.07) | (-0.02,0.03) | **(-0.18,-0.12)** | (-0.02,0.01) | (-0.12,0.00) | **(-0.19,-0.03)** | **(-0.04,-0.00)** |
| CHRS, 4 at | **-0.17** | **-0.17** | **-0.17** | 0.02 | 0.00 | **-0.16** | -0.00 | -0.04 | **-0.13** | **-0.04** |
| highest Lvl | **(-0.24,-0.09)** | **(-0.30,-0.03)** | **(-0.29,-0.04)** | (-0.07,0.10) | (-0.03,0.04) | **(-0.19,-0.13)** | (-0.02,0.02) | (-0.15,0.07) | **(-0.22,-0.03)** | **(-0.07,-0.02)** |
| CHRS, 1 at 2nd- | **-0.02** | -0.02 | -0.02 | 0.01 | 0.02 | **-0.02** | 0.00 | 0.00 | -0.02 | -0.02 |
| highest Lvl | **(-0.04,-0.00)** | (-0.07,0.03) | (-0.09,0.05) | (-0.03,0.05) | (0.01,0.04) | **(-0.03,-0.01)** | (-0.01,0.01) | (-0.04,0.04) | (-0.07,0.02) | (-0.05,0.01) |
| CHRS, 2 at 2nd- | **-0.07** | -0.07 | -0.07 | -0.01 | -0.01 | **-0.08** | -0.01 | -0.04 | -0.05 | **-0.06** |
| highest Lvl | **(-0.10,-0.04)** | (-0.15,0.01) | (-0.16,0.02) | (-0.06,0.05) | (-0.04,0.01) | **(-0.10,-0.05)** | (-0.02,0.00) | (-0.09,0.01) | (-0.11,0.01) | **(-0.09,-0.03)** |
| COVID tests L2 | 0.08 | 0.08 | 0.08 | 0.02 | -0.01 | 0.10 | -0.00 | -0.02 | 0.04 | 0.02 |
| | (0.05,0.11) | (-0.00,0.17) | (-0.00,0.16) | (-0.02,0.07) | (-0.03,0.02) | (0.07,0.12) | (-0.01,0.01) | (-0.05,0.00) | (-0.03,0.11) | (0.01,0.04) |
| COVID tests L3 | 0.05 | 0.05 | 0.05 | 0.01 | -0.01 | 0.05 | **-0.08** | **-0.21** | -0.01 | **-0.11** |
| | (0.01,0.09) | (-0.03,0.13) | (-0.04,0.13) | (-0.04,0.06) | (-0.04,0.02) | (0.02,0.09) | **(-0.14,-0.02)** | **(-0.34,-0.08)** | (-0.08,0.06) | **(-0.13,-0.08)** |
| COVID tests L4 | 0.02 | 0.02 | 0.02 | 0.01 | 0.02 | 0.03 | -0.00 | -0.04 | 0.00 | 0.01 |
| | (-0.01,0.06) | (-0.04,0.09) | (-0.06,0.10) | (-0.04,0.06) | (0.00,0.04) | (0.00,0.05) | (-0.01,0.01) | (-0.09,0.01) | (-0.04,0.05) | (-0.01,0.04) |
| Physical | -0.01 | -0.01 | -0.01 | 0.01 | -0.01 | -0.01 | -0.10 | **-0.24** | **-0.09** | **-0.17** |
| distancing L2 | (-0.05,0.03) | (-0.11,0.09) | (-0.14,0.13) | (-0.05,0.08) | (-0.03,0.01) | (-0.04,0.02) | (-0.21,0.01) | **(-0.47,-0.02)** | **(-0.16,-0.02)** | **(-0.22,-0.12)** |

TABLE III. Estimated effect sizes and confidence intervals (ctd.)

| Regressor | DK | BT | Ebisuzaki | 2WFE | ARMA(6,11) | DYN | RF1 | RF0 | elastic net | PCR |
|---|---|---|---|---|---|---|---|---|---|---|
| Masks L2 | 0.04 | 0.04 | 0.04 | -0.00 | **-0.11** | 0.01 | -0.00 | -0.04 | 0.00 | -0.02 |
| | (-0.02,0.11) | (-0.08,0.16) | (-0.05,0.14) | (-0.08,0.08) | **(-0.13,-0.09)** | (-0.06,0.09) | (-0.01,0.01) | (-0.10,0.02) | (-0.05,0.06) | (-0.05,0.02) |
| Masks L3 | -0.06 | -0.06 | -0.06 | -0.01 | **-0.16** | **-0.06** | 0.00 | -0.01 | -0.05 | **-0.03** |
| | (-0.13,0.01) | (-0.18,0.06) | (-0.16,0.04) | (-0.10,0.08) | **(-0.18,-0.13)** | **(-0.10,-0.03)** | (-0.01,0.01) | (-0.06,0.04) | (-0.11,0.01) | **(-0.05,-0.01)** |
| Masks L4 | -0.04 | -0.04 | -0.04 | -0.02 | **-0.15** | **-0.04** | -0.00 | -0.07 | -0.05 | **-0.07** |
| | (-0.10,0.03) | (-0.15,0.07) | (-0.12,0.05) | (-0.11,0.07) | **(-0.17,-0.13)** | **(-0.06,-0.01)** | (-0.02,0.01) | (-0.15,0.00) | (-0.10,0.01) | **(-0.10,-0.04)** |
| Masks L5 | -0.01 | -0.01 | -0.01 | -0.02 | **-0.12** | -0.01 | -0.00 | -0.01 | -0.02 | **-0.02** |
| | (-0.08,0.07) | (-0.13,0.12) | (-0.11,0.09) | (-0.12,0.08) | **(-0.15,-0.09)** | (-0.04,0.02) | (-0.01,0.01) | (-0.04,0.01) | (-0.08,0.03) | **(-0.04,-0.01)** |
| School holidays | 0.00 | 0.00 | 0.00 | 0.01 | -0.00 | -0.00 | -0.00 | 0.03 | -0.00 | -0.02 |
| | (-0.03,0.04) | (-0.05,0.06) | (-0.04,0.04) | (-0.02,0.05) | (-0.01,0.00) | (-0.03,0.03) | (-0.02,0.01) | (-0.02,0.09) | (-0.06,0.05) | (-0.07,0.03) |
| After school | 0.03 | 0.03 | 0.03 | 0.04 | 0.00 | 0.03 | 0.00 | -0.02 | 0.02 | **-0.02** |
| holidays | (-0.01,0.06) | (-0.04,0.09) | (-0.02,0.07) | (0.01,0.07) | (-0.00,0.01) | (0.00,0.05) | (-0.00,0.01) | (-0.06,0.02) | (-0.05,0.08) | **(-0.02,-0.01)** |
| School holidays | 0.05 | 0.05 | 0.05 | 0.02 | 0.00 | 0.06 | 0.01 | 0.04 | 0.04 | -0.00 |
| (2nd half) | (0.02,0.08) | (-0.02,0.12) | (-0.00,0.10) | (-0.01,0.06) | (-0.01,0.01) | (0.02,0.09) | (-0.01,0.02) | (-0.01,0.09) | (-0.03,0.11) | (-0.03,0.03) |
| Easter & | **-0.10** | -0.10 | **-0.10** | **-0.13** | -0.01 | **-0.10** | -0.01 | -0.04 | -0.09 | 0.00 |
| Christmas | **(-0.14,-0.06)** | (-0.20,0.00) | **(-0.15,-0.05)** | **(-0.23,-0.04)** | (-0.03,0.01) | **(-0.15,-0.04)** | (-0.02,0.00) | (-0.09,0.01) | (-0.19,0.01) | (-0.01,0.01) |
| Vaccination (1st | **-0.42** | **-0.42** | **-0.42** | -0.12 | **-0.36** | **-0.43** | **-0.15** | **-0.27** | **-0.29** | **-0.12** |
| dose) | **(-0.48,-0.36)** | **(-0.54,-0.30)** | **(-0.52,-0.32)** | (-0.36,0.12) | **(-0.40,-0.31)** | **(-0.49,-0.38)** | **(-0.21,-0.09)** | **(-0.41,-0.14)** | **(-0.44,-0.15)** | **(-0.15,-0.09)** |
| Seasonality | **0.09** | **0.09** | **0.09** | **0.15** | **0.08** | **0.09** | 0.02 | 0.13 | **0.12** | **0.11** |
| (cos) | **(0.05,0.13)** | **(0.01,0.17)** | **(0.04,0.15)** | **(0.06,0.23)** | **(0.06,0.09)** | **(0.04,0.15)** | (-0.03,0.06) | (-0.07,0.33) | **(0.05,0.19)** | **(0.07,0.16)** |
| Seasonality | **-0.07** | -0.07 | **-0.07** | **-0.10** | **-0.18** | **-0.07** | -0.00 | 0.05 | -0.03 | **-0.03** |
| (sin) | **(-0.10,-0.03)** | (-0.13,0.00) | **(-0.12,-0.01)** | **(-0.17,-0.02)** | **(-0.19,-0.16)** | **(-0.09,-0.05)** | (-0.04,0.04) | (-0.10,0.19) | (-0.10,0.05) | **(-0.07,-0.00)** |

The table shows point estimates and confidence intervals for all regressors and for all models except the baseline model (whose confidence intervals are unrealistically narrow). If an intervention is associated with lower $\mathcal{R}(t)$ according to the point estimate, entries are shown in blue, otherwise in red. The point estimates and confidence intervals for the seasonality components are not colour-coded. If the entire confidence interval spans negative values, i.e., if an association with higher $\mathcal{R}(t)$ can be excluded at a 95% confidence level, entries are marked in boldface. For the seasonality components, entries are marked in boldface if the confidence interval does not overlap with the null. Dashes indicate that (interpretable) point estimates and confidence intervals cannot be obtained with a certain method.

[1] Greene, W. H. *Econometric Analysis* (Pearson Education, 2003), fifth edn. URL http://pages.stern.nyu.edu/~wgreene/Text/econometricanalysis.htm.

[2] Dickey, D. A. & Fuller, W. A. Likelihood ratio statistics for autoregressive time series with a unit root. *Econometrica* **49**, 1057–1072 (1981). URL http://www.jstor.org/stable/1912517.

[3] Pedroni, P. Panel cointegration: Asymptotic and finite sample properties of pooled time series tests with an application to the ppp hypothesis. *Econometric Theory* **20**, 597–625 (2004).

[4] Westerlund, J. Testing for error correction in panel data. WorkingPaper 056, METEOR, Maastricht University School of Business and Economics (2006).

[5] Lauer, S. A. *et al.* The incubation period of coronavirus disease 2019 (COVID-19) from publicly reported confirmed cases: Estimation and application. *Ann. Intern. Med.* **172**, 577–582 (2020).

[6] McAloon, C. *et al.* Incubation period of COVID-19: a rapid systematic review and meta-analysis of observational research. *BMJ Open* **10** (2020). URL https://bmjopen.bmj.com/content/10/8/e039652. https://bmjopen.bmj.com/content/10/8/e039652.full.pdf.

[7] Pijpers, F. P. A non-parametric method for determining epidemiological reproduction numbers. *Journal of Mathematical Biology* **82**, 37 (2021). URL https://doi.org/10.1007/s00285-021-01590-6.

[8] Cori, A., Ferguson, N. M., Fraser, C. & Cauchemez, S. A New Framework and Software to Estimate Time-Varying Reproduction Numbers During Epidemics. *American Journal of Epidemiology* **178**, 1505–1512 (2013). URL https://doi.org/10.1093/aje/kwt133.

[9] Donnelly, C. A. *et al.* Epidemiological determinants of spread of causal agent of severe acute respiratory syndrome in hong kong. *Lancet* **361**, 1761–1766 (2003).

[10] Riley, S. *et al.* Transmission dynamics of the etiological agent of SARS in hong kong: impact of public health interventions. *Science* **300**, 1961–1966 (2003).

[11] Kreck, M. & Scholz, E. Back to the roots: A discrete Kermack-McKendrick model adapted to covid-19. *Bull. Math. Biol.* **84**, 44 (2022).

[12] Pouwels, K. B. *et al.* Community prevalence of SARS-CoV-2 in England from April to November, 2020: results from the ONS Coronavirus Infection Survey. *Lancet Public Health* **6**, e30–e38 (2021).

[13] Hastie, T. & Tibshirani, R. Generalized Additive Models. *Statistical Science* **1**, 297 – 310 (1986). URL https://doi.org/10.1214/ss/1177013604.

[14] Polack, F. P. *et al.* Safety and efficacy of the BNT162b2 mRNA covid-19 vaccine. *N. Engl. J. Med.* **383**, 2603–2615 (2020).

[15] Chemaitelly, H. *et al.* mRNA-1273 COVID-19 vaccine effectiveness against the B.1.1.7 and B.1.351 variants and severe COVID-19 disease in Qatar. *Nat. Med.* **27**, 1614–1621 (2021).

[16] Baden, L. R. *et al.* Efficacy and safety of the mRNA-1273 SARS-CoV-2 vaccine. *N. Engl. J. Med.* **384**, 403–416 (2021).

[17] Mossong, J. *et al.* Social contacts and mixing patterns relevant to the spread of infectious diseases. *PLoS Medicine* **5**, e74 (2008).

[18] Grassberger, P. On the critical behavior of the general epidemic process and dynamical percolation. *Mathematical Biosciences* **63**, 157–172 (1983). URL `https://www.sciencedirect.com/science/article/pii/0025556482900360`.

[19] Colgate, S. A., Stanley, E. A., Hyman, J. M., Layne, S. P. & Qualls, C. Risk behavior-based model of the cubic growth of acquired immunodeficiency syndrome in the united states. *Proceedings of the National Academy of Sciences* **86**, 4793–4797 (1989). URL `https://www.pnas.org/content/86/12/4793`.

[20] Pastor-Satorras, R., Castellano, C., Van Mieghem, P. & Vespignani, A. Epidemic processes in complex networks. *Rev. Mod. Phys.* **87**, 925–979 (2015). URL `https://link.aps.org/doi/10.1103/RevModPhys.87.925`.

[21] Kiss, I. Z., Green, D. M. & Kao, R. R. The effect of contact heterogeneity and multiple routes of transmission on final epidemic size. *Mathematical Biosciences* **203**, 124–136 (2006). URL `https://www.sciencedirect.com/science/article/pii/S0025556406000356`.

[22] Keeling, M. J. & Rohani, P. *Modeling Infectious Diseases in Humans and Animals* (Princeton University Press, 2008). URL `http://www.jstor.org/stable/j.ctvcm4gk0`.

[23] Chowell, G., Sattenspiel, L., Bansal, S. & Viboud, C. Mathematical models to characterize early epidemic growth: A review. *Physics of Life Reviews* **18**, 66–97 (2016). URL `https://www.sciencedirect.com/science/article/pii/S1571064516300641`.

[24] Viboud, C., Simonsen, L. & Chowell, G. A generalized-growth model to characterize the early ascending phase of infectious disease outbreaks. *Epidemics* **15**, 27–37 (2016). URL `https://www.sciencedirect.com/science/article/pii/S1755436516000037`.

[25] Neipel, J., Bauermann, J., Bo, S., Harmon, T. & Jülicher, F. Power-law population heterogeneity governs epidemic waves. *PLoS ONE* **15**, e0239678 (2020). 2008.00471.

[26] Britton, T., Ball, F. & Trapman, P. A mathematical model reveals the influence of population heterogeneity on herd immunity to SARS-CoV-2. *Science* **369**, 846–849 (2020). URL https://www.science.org/doi/abs/10.1126/science.abc6810.

[27] Gomes, M. G. M. *et al.* Individual variation in susceptibility or exposure to SARS-CoV-2 lowers the herd immunity threshold. *Journal of Theoretical Biology* **540**, 111063 (2022). URL https://www.sciencedirect.com/science/article/pii/S0022519322000613.

[28] an der Heiden, M. & Buchholz, U. Modellierung von Beispielszenarien der SARS-CoV-2-Epidemie 2020 in Deutschland (2020).

[29] Davies, N. G. *et al.* Estimated transmissibility and impact of SARS-CoV-2 lineage b.1.1.7 in england. *Science* **372**, eabg3055 (2021).

[30] Liu, Y. & Rocklöv, J. The reproductive number of the Delta variant of SARS-CoV-2 is far higher compared to the ancestral SARS-CoV-2 virus. *Journal of Travel Medicine* **28**, taab124 (2021). URL https://doi.org/10.1093/jtm/taab124. https://academic.oup.com/jtm/article-pdf/28/7/taab124/41825935/taab124.pdf.

[31] Yu, P., Zhu, J., Zhang, Z. & Han, Y. A familial cluster of infection associated with the 2019 novel coronavirus indicating possible person-to-person transmission during the incubation period. *J. Infect. Dis.* **221**, 1757–1761 (2020).

[32] Liu, Y., Gayle, A. A., Wilder-Smith, A. & Rocklöv, J. The reproductive number of COVID-19 is higher compared to SARS coronavirus. *Journal of Travel Medicine* **27**, taaa021 (2020). URL https://doi.org/10.1093/jtm/taaa021.

[33] an der Heiden, M., Hicketier, A. & Bremer, V. Wirksamkeit und Wirkung von anti-epidemischen Maßnahmen auf die COVID-19-Pandemie in Deutschland (StopptCOVID-Studie) (2023). URL https://www.rki.de/DE/Content/InfAZ/N/Neuartiges_Coronavirus/Projekte_RKI/StopptCOVID_studie.html.

[34] Murphy, C. *et al.* Effectiveness of social distancing measures and lockdowns for reducing transmission of COVID-19 in non-healthcare, community-based settings. *Philos. Trans. A Math. Phys. Eng. Sci.* **381**, 20230132 (2023).

[35] Kermack, W. & McKendrick, A. A contribution to the mathematical theory of epidemics. *Proc. R. Soc. Lond. Ser. Math. Phys. Eng. Sci.* **115**, 700–721 (1927).

[36] Anderson, R. M. & May, R. M. Population biology of infectious diseases: Part I. *Nature* **280**, 361–367 (1979).

[37] Kalman, R. E. A new approach to linear filtering and prediction problems. *Journal of Basic Engineering* **82**, 35–45 (1960). URL https://doi.org/10.1115/1.3662552.

[38] Harvey, A. *State space models*, 269–275 (Palgrave Macmillan UK, London, 2010). URL https://doi.org/10.1057/9780230280830_30.

[39] Tao, S., Bragazzi, N. L., Wu, J., Mellado, B. & Kong, J. D. Harnessing Artificial Intelligence to assess the impact of nonpharmaceutical interventions on the second wave of the Coronavirus Disease 2019 pandemic across the world. *Sci. Rep.* **12**, 944 (2022).

[40] Nader, I. W., Zeilinger, E. L., Jomar, D. & Zauchner, C. Onset of effects of non-pharmaceutical interventions on COVID-19 infection rates in 176 countries. *BMC Public Health* **21**, 1472 (2021).

[41] Wagner, A. K., Soumerai, S. B., Zhang, F. & Ross-Degnan, D. Segmented regression analysis of interrupted time series studies in medication use research. *Journal of Clinical Pharmacy and Therapeutics* **27**, 299–309 (2002). URL https://onlinelibrary.wiley.com/doi/abs/10.1046/j.1365-2710.2002.00430.x.

[42] Muggeo, V. M. R. Estimating regression models with unknown break-points. *Statistics in Medicine* **22**, 3055–3071 (2003). URL https://onlinelibrary.wiley.com/doi/abs/10.1002/sim.1545.

[43] Celestin Hategeka, Hinda Ruton, Mohammad Karamouzian, Larry D Lynd & Michael R Law. Use of interrupted time series methods in the evaluation of health system quality improvement interventions: a methodological systematic review. *BMJ Global Health* **5**, e003567 (2020).

[44] Card, D. & Krueger, A. B. Minimum Wages and Employment: A Case Study of the Fast-Food Industry in New Jersey and Pennsylvania. *American Economic Review* **84**, 772–793 (1994). URL https://ideas.repec.org/a/aea/aecrev/v84y1994i4p772-93.html.

[45] Lechner, M. The Estimation of Causal Effects by Difference-in-Difference Methods. University of St. Gallen Department of Economics working paper series 2010 2010-28, Department of Economics, University of St. Gallen (2010). URL https://ideas.repec.org/p/usg/dp2010/2010-28.html.

[46] Kreif, N. *et al.* Examination of the synthetic control method for evaluating health policies with multiple treated units. *Health Econ.* **25**, 1514–1528 (2016).

[47] Bouttell, J., Craig, P., Lewsey, J., Robinson, M. & Popham, F. Synthetic control methodology as a tool for evaluating population-level health interventions. *J. Epidemiol. Community Health* **72**, 673–678 (2018).

[48] Wieland, T. A phenomenological approach to assessing the effectiveness of COVID-19 related non-

pharmaceutical interventions in germany. *Saf. Sci.* **131**, 104924 (2020).

[49] Casini, L. & Roccetti, M. Reopening italy's schools in september 2020: a bayesian estimation of the change in the growth rate of new SARS-CoV-2 cases. *BMJ Open* **11**, e051458 (2021).

[50] Guo, C. *et al.* Physical distancing implementation, ambient temperature and Covid-19 containment: An observational study in the United States. *Sci. Total Environ.* **789**, 147876 (2021).

[51] Küchenhoff, H., Günther, F., Höhle, M. & Bender, A. Analysis of the early COVID-19 epidemic curve in Germany by regression models with change points. *Epidemiol. Infect.* **149**, e68 (2021).

[52] Staguhn, E. D., Weston-Farber, E. & Castillo, R. C. The impact of statewide school closures on COVID-19 infection rates. *Am. J. Infect. Control* **49**, 503–505 (2021).

[53] Yehya, N., Venkataramani, A. & Harhay, M. O. Statewide interventions and coronavirus disease 2019 mortality in the united states: An observational study. *Clin. Infect. Dis.* **73**, e1863–e1869 (2021).

[54] Boutzoukas, A. E. *et al.* Secondary transmission of COVID-19 in K-12 schools: Findings from 2 states. *Pediatrics* **149** (2022).

[55] Sims, C. A. Macroeconomics and reality. *Econometrica* **48**, 1–48 (1980). URL `http://www.jstor.org/stable/1912017`.

[56] Stock, J. H. & Watson, M. W. Vector Autoregressions. *Journal of Economic Perspectives* **15**, 101–115 (2001). URL `https://ideas.repec.org/a/aea/jecper/v15y2001i4p101-115.html`.

[57] Pleninger, R., Streicher, S. & Sturm, J.-E. Do COVID-19 containment measures work? evidence from switzerland. *Schweiz. Z. Volkswirtsch. Stat.* **158**, 5 (2022).

[58] Keeling, M. J., Woolhouse, M. E. J., May, R. M., Davies, G. & Grenfell, B. T. Modelling vaccination strategies against foot-and-mouth disease. *Nature* **421**, 136–142 (2003).

[59] Riley, S., Eames, K., Isham, V., Mollison, D. & Trapman, P. Five challenges for spatial epidemic models. *Epidemics* **10**, 68–71 (2015). URL `https://www.sciencedirect.com/science/article/pii/S1755436514000310`. Challenges in Modelling Infectious DIsease Dynamics.

[60] Flaxman, S. *et al.* Estimating the effects of non-pharmaceutical interventions on COVID-19 in europe. *Nature* **584**, 257–261 (2020).

[61] Brauner, J. M. *et al.* Inferring the effectiveness of government interventions against COVID-19. *Science* **371**, eabd9338 (2021).

[62] Hunter, P. R., Colón-González, F. J., Brainard, J. & Rushton, S. Impact of non-pharmaceutical interventions against COVID-19 in europe in 2020: a quasi-experimental non-equivalent group and time series design study. *Euro Surveill.* **26** (2021).

[63] Sharma, M. *et al.* Understanding the effectiveness of government interventions against the resurgence of COVID-19 in europe. *Nat. Commun.* **12**, 5820 (2021).

[64] Khazaei, Y., Küchenhoff, H., Hoffmann, S., Syliqi, D. & Rehms, R. Using a Bayesian hierarchical approach to study the association between non-pharmaceutical interventions and the spread of Covid-19 in Germany. *Sci. Rep.* **13**, 18900 (2023).

[65] Greenland, S. *et al.* Statistical tests, P values, confidence intervals, and power: a guide to misinterpretations. *Eur. J. Epidemiol.* **31**, 337–350 (2016).

[66] Hespanhol, L., Vallio, C. S., Costa, L. M. & Saragiotto, B. T. Understanding and interpreting confidence and credible intervals around effect estimates. *Braz. J. Phys. Ther.* **23**, 290–301 (2019).

[67] White, H. A heteroskedasticity-consistent covariance matrix estimator and a direct test for heteroskedasticity. *Econometrica* **48**, 817–838 (1980). URL `http://www.jstor.org/stable/1912934`.

[68] Newey, W. K. & West, K. D. A Simple, Positive Semi-definite, Heteroskedasticity and Autocorrelation Consistent Covariance Matrix. *Econometrica* **55**, 703–708 (1987). URL `https://ideas.repec.org/a/ecm/emetrp/v55y1987i3p703-08.html`.

[69] Driscoll, J. C. & Kraay, A. C. Consistent Covariance Matrix Estimation With Spatially Dependent Panel Data. *The Review of Economics and Statistics* **80**, 549–560 (1998). URL `https://ideas.repec.org/a/tpr/restat/v80y1998i4p549-560.html`.

[70] Hoechle, D. Robust standard errors for panel regressions with cross-sectional dependence. *Stata Journal* **7**, 281–312 (2007). URL `https://ideas.repec.org/a/tsj/stataj/v7y2007i3p281-312.html`.

[71] Berndt, E. R., Hall, B., Hall, R. & Hausman, J. Estimation and Inference in Nonlinear Structural Models. In *Annals of Economic and Social Measurement, Volume 3, number 4*, 653–665 (National Bureau of Economic Research, Inc, 1974). URL `https://EconPapers.repec.org/RePEc:nbr:nberch:10206`.

[72] Efron, B. Bootstrap Methods: Another Look at the Jackknife. *The Annals of Statistics* **7**, 1 – 26 (1979). URL `https://doi.org/10.1214/aos/1176344552`.

[73] Athreya, K. B. Bootstrap of the mean in the infinite variance case. *The Annals of Statistics* **15**, 724–731 (1987). URL `http://www.jstor.org/stable/2241336`.

[74] CHATTERJEE, A. & LAHIRI, S. N. Asymptotic properties of the residual bootstrap for lasso estimators. *Proceedings of the American Mathematical Society* **138**, 4497–4509 (2010). URL

http://www.jstor.org/stable/41059185.

[75] Chatterjee, A. & Lahiri, S. N. Bootstrapping lasso estimators. *Journal of the American Statistical Association* **106**, 608–625 (2011). URL https://doi.org/10.1198/jasa.2011.tm10159. https://doi.org/10.1198/jasa.2011.tm10159.

[76] Hall, P. Theoretical comparison of bootstrap confidence intervals. *The Annals of Statistics* **16**, 927–953 (1988). URL http://www.jstor.org/stable/2241604.

[77] Mavragani, A. & Gkillas, K. Exploring the role of non-pharmaceutical interventions (NPIs) in flattening the greek COVID-19 epidemic curve. *Sci. Rep.* **11**, 11741 (2021).

[78] Liu, Y. *et al.* The impact of non-pharmaceutical interventions on SARS-CoV-2 transmission across 130 countries and territories. *BMC Med.* **19**, 40 (2021).

[79] Barros, V. *et al.* A causal inference approach for estimating effects of non-pharmaceutical interventions during covid-19 pandemic. *PLoS One* **17**, e0265289 (2022).

[80] Barbeito, I. *et al.* Effectiveness of non-pharmaceutical interventions in nine fields of activity to decrease SARS-CoV-2 transmission (spain, september 2020-may 2021). *Front. Public Health* **11**, 1061331 (2023).

[81] Kunsch, H. R. The Jackknife and the Bootstrap for General Stationary Observations. *The Annals of Statistics* **17**, 1217 – 1241 (1989). URL https://doi.org/10.1214/aos/1176347265.

[82] Politis, D. N. & Romano, J. P. The stationary bootstrap. *Journal of the American Statistical Association* **89**, 1303–1313 (1994). URL https://doi.org/10.1080/01621459.1994.10476870.

[83] Kreiss, J.-P. & Lahiri, S. N. Bootstrap Methods for Time Series. In Subba Rao, T., Subba Rao, S. & Rao, C. (eds.) *Time Series Analysis: Methods and Applications*, vol. 30 of *Handbook of Statistics*, chap. 1, 3–26 (Elsevier, 2012). URL https://www.sciencedirect.com/science/article/pii/B9780444538581000016.

[84] Shao, X. The dependent wild bootstrap. *Journal of the American Statistical Association* **105**, 218–235 (2010). URL https://doi.org/10.1198/jasa.2009.tm08744. https://doi.org/10.1198/jasa.2009.tm08744.

[85] Kapetanios, G. A bootstrap procedure for panel data sets with many cross-sectional units. *The Econometrics Journal* **11**, 377–395 (2008). URL http://www.jstor.org/stable/23116081.

[86] Gonçalves, S. The moving blocks bootstrap for panel linear regression models with individual fixed effects. *Econometric Theory* **27**, 1048–1082 (2011).

[87] Jiti, G., Bin, P. & Yayi, Y. A Simple Bootstrap Method for Panel Data Inferences (2022). URL

https://bridges.monash.edu/articles/journal_contribution/A_Simple_Bootstrap_Method_for_Panel_Data_Inferences/21531603.

[88] Ebisuzaki, W. A Method to Estimate the Statistical Significance of a Correlation When the Data Are Serially Correlated. *Journal of Climate* **10**, 2147–2153 (1997).

[89] Akaike, H. *Information Theory and an Extension of the Maximum Likelihood Principle*, 199–213 (Springer New York, New York, NY, 1998). URL https://doi.org/10.1007/978-1-4612-1694-0_15.

[90] Hansen, P. C. Truncated Singular Value Decomposition Solutions to Discrete Ill-Posed Problems with Ill-Determined Numerical Rank. *SIAM Journal on Scientific and Statistical Computing* **11**, 503–518 (1990). URL https://doi.org/10.1137/0911028. https://doi.org/10.1137/0911028.

[91] Xu, P. Truncated SVD methods for discrete linear ill-posed problems. *Geophysical Journal International* **135**, 505–514 (1998). URL https://doi.org/10.1046/j.1365-246X.1998.00652.x.

[92] Wold, S., Sjöström, M. & Eriksson, L. Pls-regression: a basic tool of chemometrics. *Chemometrics and Intelligent Laboratory Systems* **58**, 109–130 (2001). URL https://www.sciencedirect.com/science/article/pii/S0169743901001551. PLS Methods.

[93] Tikhonov, A. N. Solution of Incorrectly Formulated Problems and the Regularization Method. *Soviet Mathematics Doklady* **14**, 1035–1038 (1963).

[94] Hoerl, A. E. & Kennard, R. W. Ridge regression: Biased estimation for nonorthogonal problems. *Technometrics* **12**, 55–67 (1970). URL http://www.jstor.org/stable/1267351.

[95] Tibshirani, R. Regression shrinkage and selection via the lasso. *Journal of the Royal Statistical Society: Series B (Methodological)* **58**, 267–288 (1996). URL https://rss.onlinelibrary.wiley.com/doi/abs/10.1111/j.2517-6161.1996.tb02080.x.

[96] Zou, H. & Hastie, T. Regularization and variable selection via the elastic net. *Journal of the Royal Statistical Society Series B: Statistical Methodology* **67**, 301–320 (2005). URL https://doi.org/10.1111/j.1467-9868.2005.00503.x.

[97] Efroymson, M. A. Multiple regression analysis. In *Mathematical Methods for Digital Computers* (John Wiley, New York, 1960).

[98] Huntley, K. S. *et al.* Associations of Stay-at-Home Order Enforcement With COVID-19 Population Outcomes: An Interstate Statistical Analysis. *American journal of epidemiology* **191**, 561—569 (2022). URL https://europepmc.org/articles/PMC8780467.

[99] Miller, A. J. Selection of subsets of regression variables. *Journal of the Royal Statistical Society:*

*Series A (General)* **147**, 389–410 (1984). URL https://rss.onlinelibrary.wiley.com/doi/abs/10.2307/2981576.

[100] Altman, D. G. & Andersen, P. K. Bootstrap investigation of the stability of a Cox regression model. *Statistics in Medicine* **8**, 771–783 (1989). URL https://onlinelibrary.wiley.com/doi/abs/10.1002/sim.4780080702.

[101] Hurvich, C. M. & Tsai, C.-L. The impact of model selection on inference in linear regression. *The American Statistician* **44**, 214–217 (1990). URL http://www.jstor.org/stable/2685338.

[102] Smith, G. Step away from stepwise. *Journal of Big Data* **5**, 32 (2018). URL https://doi.org/10.1186/s40537-018-0143-6.

[103] Bergmeir, C. & Benítez, J. M. On the use of cross-validation for time series predictor evaluation. *Information Sciences* **191**, 192–213 (2012). URL https://www.sciencedirect.com/science/article/pii/S0020025511006773. Data Mining for Software Trustworthiness.

[104] Newey, W. K. & West, K. D. Automatic Lag Selection in Covariance Matrix Estimation. *The Review of Economic Studies* **61**, 631–653 (1994). URL https://doi.org/10.2307/2297912.

[105] Welch, P. The use of fast Fourier transform for the estimation of power spectra: A method based on time averaging over short, modified periodograms. *IEEE Transactions on Audio and Electroacoustics* **15**, 70–73 (1967).

[106] von Storch, H. & Zwiers, F. W. *Statistical Analysis in Climate Research* (Cambridge University Press, 1999).

[107] Müller, B., Janka, H.-T. & Marek, A. A New Multi-dimensional General Relativistic Neutrino Hydrodynamics Code of Core-collapse Supernovae. III. Gravitational Wave Signals from Supernova Explosion Models. *Astrophys. J.* **766**, 43 (2013). 1210.6984.

[108] Sheppard, K. *et al.* bashtage/arch: Release 7.2 (2024). URL https://doi.org/10.5281/zenodo.593254.

[109] Politis, D. N. & White, H. Automatic block-length selection for the dependent bootstrap. *Econometric Reviews* **23**, 53–70 (2004). URL https://doi.org/10.1081/ETC-120028836.

[110] Andrew Patton, D. N. P. & White, H. Correction to "Automatic Block-Length Selection for the Dependent Bootstrap" by D. Politis and H. White. *Econometric Reviews* **28**, 372–375 (2009). URL https://doi.org/10.1080/07474930802459016.

[111] Wooldridge, J. M. Two-way fixed effects, the two-way mundlak regression, and difference-in-differences estimators (2021). URL https://ssrn.com/abstract=3906345.

[112] de Chaisemartin, C. & D'Haultfœuille, X. Two-way fixed effects and differences-in-differences estimators with several treatments. *Journal of Econometrics* **236**, 105480 (2023). URL https://www.sciencedirect.com/science/article/pii/S0304407623001963.

[113] Fowler, J. H., Hill, S. J., Levin, R. & Obradovich, N. Stay-at-home orders associate with subsequent decreases in COVID-19 cases and fatalities in the United States. *PLOS ONE* **16**, 1–15 (2021). URL https://doi.org/10.1371/journal.pone.0248849.

[114] Goodman-Bacon, A. Difference-in-differences with variation in treatment timing. *Journal of Econometrics* **225**, 254–277 (2021). URL https://www.sciencedirect.com/science/article/pii/S0304407621001445. Themed Issue: Treatment Effect 1.

[115] Imai, K. & Kim, I. S. On the Use of Two-Way Fixed Effects Regression Models for Causal Inference with Panel Data. *Political Analysis* **29**, 405–415 (2021). URL https://ideas.repec.org/a/cup/polals/v29y2021i3p405-415_8.html.

[116] Fulton, C. Estimating time series models by state space methods in Python: Statsmodels (2017). URL https://www.chadfulton.com/files/fulton_statsmodels_2017_v1.pdf.

[117] Harvey, A. C. *Time series models* (Harvester Wheatsheaf, London, 1993), 2nd ed. edn.

[118] Adak, S. Time-Dependent Spectral Analysis of Nonstationary Time Series. *Journal of the American Statistical Association* **93**, 1488–1501 (1998). URL https://www.tandfonline.com/doi/abs/10.1080/01621459.1998.10473808.

[119] Davis, R. A., Lee, T. C. M. L. & Rodriguez-Yam, G. A. Structural Break Estimation for Nonstationary Time Series Models. *Journal of the American Statistical Association* **101**, 223–239 (2006). URL https://doi.org/10.1198/016214505000000745.

[120] Rosen, O., Wood, S. & Stoffer, D. S. Adaptspec: Adaptive spectral estimation for nonstationary time series. *Journal of the American Statistical Association* **107**, 1575–1589 (2012). URL https://doi.org/10.1080/01621459.2012.716340.

[121] Bertolacci, M., Rosen, O., Cripps, E. & Cripps, S. Adaptspec-x: Covariate-dependent spectral modeling of multiple nonstationary time series. *Journal of Computational and Graphical Statistics* **31**, 436–454 (2022). URL https://doi.org/10.1080/10618600.2021.2000870.

[122] Schwarz, G. Estimating the Dimension of a Model. *The Annals of Statistics* **6**, 461–464 (1978). URL http://www.jstor.org/stable/2958889.

[123] Brauer, F. The Kermack–McKendrick epidemic model revisited. *Mathematical Biosciences* **198**, 119–131 (2005). URL https://www.sciencedirect.com/science/article/pii/S0025556

405001331.

[124] MacKinnon, J. G. Bootstrap Methods In Econometrics. Working Paper 1028, Economics Department, Queen's University (2006). URL https://ideas.repec.org/p/qed/wpaper/1028.html.

[125] Fernández-Casal, R., Castillo-Páez, S. & Flores, M. A nonparametric bootstrap method for heteroscedastic functional data. *Journal of Agricultural, Biological and Environmental Statistics* **29**, 169–184 (2024). URL https://doi.org/10.1007/s13253-023-00561-2.

[126] Breiman, L. Random forests. *Machine Learning* **45**, 5–32 (2001).

[127] Pedregosa, F. *et al.* Scikit-learn: Machine learning in Python. *Journal of Machine Learning Research* **12**, 2825–2830 (2011).

[128] Borisov, V. *et al.* Deep neural networks and tabular data: A survey. *IEEE Transactions on Neural Networks and Learning Systems* **35**, 7499–7519 (2024).

[129] Shapley, L. S. *17. A Value for n-Person Games*, 307–318 (Princeton University Press, Princeton, 1953). URL https://doi.org/10.1515/9781400881970-018.

[130] Lundberg, S. M. *et al.* From local explanations to global understanding with explainable AI for trees. *Nature Machine Intelligence* **2**, 56–67 (2020).

[131] URL https://shap.readthedocs.io/.
